# Genome-wide association study of placental weight in 179,025 children and parents reveals distinct and shared genetic influences between placental and fetal growth

**DOI:** 10.1101/2022.11.25.22282723

**Authors:** Robin N. Beaumont, Christopher Flatley, Marc Vaudel, Xiaoping Wu, Jing Chen, Gunn-Helen Moen, Line Skotte, Øyvind Helgeland, Pol Sole-Navais, Karina Banasik, Clara Albiñana, Justiina Ronkainen, João Fadista, Sara Elizabeth Stinson, Katerina Trajanoska, Carol A Wang, David Westergaard, Sundararajan Srinivasan, Carlos Sánchez-Soriano, Jose Ramon Bilbao, Catherine Allard, Marika Groleau, Teemu Kuulasmaa, Daniel J. Leirer, Frédérique White, Pierre-Étienne Jacques, Haoxiang Cheng, Ke Hao, Ole A. Andreassen, Bjørn Olav Åsvold, Mustafa Atalay, Laxmi Bhatta, Luigi Bouchard, Ben Michael Brumpton, Søren Brunak, Jonas Bybjerg-Grauholm, Cathrine Ebbing, Paul Elliott, Line Engelbrechtsen, Christian Erikstrup, Marisa Estarlich, Steve Franks, Romy Gaillard, Frank Geller, Jakob Grove, David M Hougaard, Eero Kajantie, Camilla S. Morgen, Ellen A Nohr, Mette Nyegaard, Colin NA Palmer, Ole Birger Pedersen, The Early Growth Genetics (EGG) Consortium, Fernando Rivadeneira, Sylvain Sebert, Beverley M. Shields, Camilla Stoltenberg, Ida Surakka, Lise Wegner Thørner, Henrik Ullum, Marja Vaarasmaki, Bjarni J Vilhjalmsson, Cristen J. Willer, Timo A. Lakka, Dorte Jensen Gybel-Brask, Mariona Bustamante, Torben Hansen, Ewan R Pearson, Rebecca Reynolds, Sisse R. Ostrowski, Craig E Pennell, Vincent W. V. Jaddoe, Janine F Felix, Andrew T. Hattersley, Mads Melbye, Deborah A. Lawlor, Kristian Hveem, Thomas Werge, Henriette Svarre Nielsen, Per Magnus, David M Evans, Bo Jacobsson, Marjo-Riitta Järvelin, Ge Zhang, Marie-France Hivert, Stefan Johansson, Rachel M. Freathy, Bjarke Feenstra, Pål R. Njølstad

**Affiliations:** Department of Clinical and Biomedical Sciences, Faculty of Health and Life Sciences, University of Exeter, Exeter, UK; Department of Obstetrics and Gynecology, Institute of Clinical Sciences, Sahlgrenska Academy, University of Gothenburg, Gothenburg, Sweden; Department of Genetics and Bioinformatics, Health Data and Digitalization, Norwegian Institute of Public Health, Oslo, Norway; Center for Diabetes Research, Department of Clinical Science, University of Bergen, Bergen, Norway; Department of Epidemiology Research, Statens Serum Institut, Copenhagen, Denmark; Division of Biomedical Informatics, Cincinnati Children’s Hospital Medical Center, Department of Pediatrics, University of Cincinnati College of Medicine, Cincinnati, Ohio, United States of America; Institute of Clinical Medicine, Faculty of Medicine, University of Oslo, Norway; University of Queensland Diamantina Institute, The University of Queensland, Brisbane, Australia; K.G. Jebsen Center for Genetic Epidemiology, Department of Public Health and Nursing, NTNU, Norwegian University of Science and Technology, Norway; Population Health Science, Bristol Medical School, University of Bristol, UK; Institute for Molecular Bioscience, The University of Queensland, Brisbane, Australia; Translational Disease Systems Biology, Novo Nordisk Foundation Center for Protein Research, University of Copenhagen, Copenhagen, Denmark; National Centre for Register-Based Research, Aarhus University, Aarhus, Denmark; Research Unit of Population Health, University of Oulu, Oulu, Finland; Department of Clinical Sciences, Lund University Diabetes Centre, Malmö, Sweden; Institute for Molecular Medicine Finland (FIMM), University of Helsinki, Helsinki, Finland; Novo Nordisk Foundation Center for Basic Metabolic Research, Faculty of Health and Medical Sciences, University of Copenhagen, Copenhagen, Denmark; Department of Internal Medicine, Erasmus MC, University Medical Center Rotterdam, Rotterdam, The Netherlands; Department of Human Genetics, McGill University, Montréal, Québec, Canada; School of Medicine and Public Health, College of Medicine, Public Health and Wellbeing, The University of Newcastle, Newcastle, New South Wales, Australia, 2308; Hunter Medical Research Institute, New Lambton Heights, New South Wales, Australia; Dept. of Obstetrics and Gynecology, Copenhagen University Hospital, Hvidovre, Denmark; Methods and Analysis, Statistics Denmark. Copenhagen, Denmark; Division of Population Health & Genomics, School of Medicine, University of Dundee, Dundee; Centre for Cardiovascular Science, The University of Edinburgh, Edinburgh, United Kingdom; Dpt. of Genetics, Physical Anthropology and Animal Physiology, University of the Basque Country (UPV/EHU) Leioa, Spain; Biocruces-Bizkaia Health Research Institute, Barakaldo, Spain; Spanish Biomedical Research Center in Diabetes and Associated Metabolic Disorders (CIBERDEM), Spain; Centre de recherche du Centre Hospitalier de l’Universite de Sherbrooke, Sherbrooke, QC; Département de Biologie, Faculté des Sciences, Université de Sherbrooke, Sherbrooke, Québec, Canada; Institute of Biomedicine, School of Medicine, University of Eastern Finland, Kuopio Campus, Finland; Icahn School of Medicine at Mount Sinai; NORMENT Centre, Institute of Clinical Medicine, University of Oslo, Oslo, Norway; Division of Mental Health and Addiction, Oslo University Hospital, NO-0315 Oslo, Norway; K.G. Jebsen Center for Genetic Epidemiology, Department of Public Health and Nursing, NTNU, Norwegian University of Science and Technology, Trondheim, Norway; HUNT Research Centre, Department of Public Health and Nursing, NTNU, Norwegian University of Science and Technology, Levanger, Norway; Department of Endocrinology, Clinic of Medicine, St. Olavs Hospital, Trondheim University Hospital, Trondheim, Norway; Department of Biochemistry and Functional Genomics, Faculty of Medicine and Health Sciences, Université de Sherbrooke, Sherbrooke, Québec, Canada; Clinical Department of Laboratory Medicine, Centre intégré universitaire de santé et de services sociaux (CIUSSS) du Saguenay–Lac-St-Jean – Hôpital Universitaire de Chicoutimi, Saguenay, Québec, Canada; Clinic of Medicine, St. Olavs Hospital, Trondheim University Hospital, Trondheim, Norway; Department for Congenital Disorders, Statens Serum Institut, Copenhagen, Denmark; iPSYCH, The Lundbeck Foundation Initiative for Integrative Psychiatric Research, Aarhus, Denmark; Department of Clinical Science, University of Bergen, Bergen, Norway; Department of Gynecology and Obstetrics, Haukeland University Hospital, Bergen, Norway; Department of Epidemiology and Biostatistics, School of Public Health, Imperial College London, London, United Kingdom; Department of Obstetrics and Gynecology, Hillerød Hospital, Denmark; Dept. Clinical Immunology, Aarhus University Hospital, Aarhus, Denmark; Dept. Clinical Medicine, Aarhus University, Aarhus, Denmark; Faculty of nursing and chiropody, Universitat de València, C/Menendez Pelayo, Valencia, Spain; Epidemiology and Environmental Health Joint Research Unit, Foundation for the Promotion of Health and Biomedical Research in the Valencian Region, FISABIO-Public Health, FISABIO-Universitat Jaume I-Universitat de València, Av. Catalunya 21, Valencia, Spain; Spanish Consortium for Research on Epidemiology and Public Health (CIBERESP), Madrid, Spain; Institute of Reproductive and Developmental Biology, Imperial College London, United Kingdom; The Generation R Study Group, Erasmus MC, University Medical Center Rotterdam, Rotterdam, The Netherlands; Department of Pediatrics, Erasmus MC, University Medical Center Rotterdam, Rotterdam, The Netherlands; Department of Biomedicine – Human Genetics and the iSEQ Center, Aarhus University, Aarhus, Denmark; Center for Genomics and Personalized Medicine, Aarhus, Denmark; Bioinformatics Research Centre, Aarhus University, Aarhus, Denmark; PEDEGO Research Unit, Medical Research Center, University of Oulu and Oulu University Hospital, Oulu, Finland; Population Health Unit, Finnish Institute for Health and Welfare, Helsinki and Oulu, Finland; National Institute of Public Health, University of Southern Denmark, Copenhagen, Denmark; Institute of Clinical research, University of Southern Denmark, Odense, Denmark; Department of Health Science and Technology, Aalborg University, Denmark; Dept. of Clinical Immunology, Zealand University Hospital, Køge, Denmark; Dept. of Clinical Medicine, Faculty of Health and Medical Sciences, University of Copenhagen, Copenhagen, Denmark; Department for Genomics of Common Diseases, School of Medicine, Imperial College London, London United Kingdom; Department of Clinical and Molecular Medicine, Norwegian University of Science and Technology, Trondheim, Norway; Pediatric Research Centre, Helsinki University Hospital and University of Helsinki, Helsinki, Finlan; Norwegian Institute of Public Health, Oslo, Norway; Department of Global Public Health and Primary Care, University of Bergen, Bergen, Norway; Department of Internal Medicine, Division of Cardiology, University of Michigan, Ann Arbor, MI 48109, USA; Dept. of Clinical Immunology, Copenhagen University Hospital, Rigshospitalet, Copenhagen, Denmark; Statens Serum Institut, Copenhagen, Denmark; PEDEGO Research Unit, Medical Research Center, University of Oulu, Oulu, Finland; Department of Obstetrics and Gynaecology, Oulu University Hospital, Oulu, Finland; Department of Biostatistics and Center for Statistical Genetics, University of Michigan, Ann Arbor, MI, USA; Department of Human Genetics, University of Michigan, Ann Arbor, MI, USA; Department of Clinical Physiology and Nuclear Medicine, Kuopio University Hospital, Kuopio, Finland; Foundation for Research in Health Exercise and Nutrition, Kuopio Research Institute of Exercise Medicine, Kuopio, Finland; Psychotherapeutic Outpatient Clinic, Mental Health Services, Capital Region, Copenhagen Denmark; ISGlobal, Institute for Global Health, Barcelona, Spain; Universitat Pompeu Fabra (UPF), Barcelona, Spain; Dept. of Clinical Medicine, University of Copenhagen, Copenhagen, Denmark; Department of Clinical Medicine, University of Copenhagen, Copenhagen, Denmark; K.G. Jebsen Center for Genetic Epidemiology, Norwegian University of Science and Technology, Trondheim, Norway; Department of Genetics, Stanford University School of Medicine, Stanford, California, USA; Medical Research Council Integrative Epidemiology Unit, University of Bristol, Bristol, United Kingdom; Population Health Science, Bristol Medical School, University of Bristol, Bristol, UK; Institute of Biological Psychiatry, Mental Health Services, Copenhagen University Hospital, Copenhagen, Denmark; Lundbeck Center for Geogenetics, GLOBE Institute, University of Copenhagen, Copenhagen, Denmark; Centre for Fertility and Health, Norwegian Institute of Public Health, Oslo, Norway; MRC Centre for Environment and Health, School of Public Health, Imperial College London, London; Unit of Primary Health Care, Oulu University Hospital, OYS, Oulu, Finland; Division of Human Genetics, Cincinnati Children’s Hospital Medical Center, Department of Pediatrics, University of Cincinnati College of Medicine, Cincinnati, Ohio, United States of America; Center for Prevention of Preterm Birth, Perinatal Institute, Cincinnati Children’s Hospital Medical Center and March of Dimes Prematurity Research Center Ohio Collaborative, Cincinnati Children’s Hospital Medical Center, Department of Pediatrics, University of Cincinnati College of Medicine, Cincinnati, Ohio, United States of America; Department of Population Medicine, Harvard Medical School, Harvard Pilgrim Health Care Institute, Boston, MA; Diabetes Unit, Massachusetts General Hospital, MA, Boston; Department of Medical Genetics, Haukeland University Hospital, Bergen, Norway; Children and Youth Clinic, Haukeland University Hospital, Bergen, Norway

## Abstract

A well-functioning placenta is essential for fetal and maternal health throughout pregnancy. Using placental weight after term delivery as a proxy for placental growth, we report genome-wide association analyses in the fetal (*n* = 65,405), maternal (*n* = 61,228), and paternal (*n* = 52,392) genomes, yielding 40 independent association signals. Twenty-six signals are confidently classified as fetal only, four maternal only, and three fetal and maternal. A maternal parent-of-origin effect is seen near *KCNQ1*. Genetic correlation and colocalization analyses reveal overlap with birth weight genetics, but twelve loci are classified as predominantly or only affecting placental weight, with connections to placental development and morphology, and transport of antibodies and amino acids. Mendelian randomization analyses indicate that fetal genetically mediated higher placental weight is causally associated with risk of preeclampsia or shorter gestational duration. Moreover, these analyses support a role for insulin produced by the fetus in regulating the growth of the placenta, providing a key link between fetal and placental growth.

## Main

The placenta is a remarkable pregnancy organ formed after implantation of the blastocyst into the uterine wall. The intimate placental connection between mother and fetus provides nutrients and oxygen to the developing fetus, while removing waste products from the fetal blood. The placenta produces hormones, growth factors, and cytokines, and allows maternal IgG antibodies to be passed to the fetus, giving newborns innate immunity. Suboptimal placentation can lead to intrauterine growth restriction^1^, spontaneous abortion, preterm birth^2^, and preeclampsia^3,4^, and a poorly functioning placenta is associated with higher risk of growth restriction^5^, adverse neurodevelopment^6^, and cardiometabolic diseases^7–11^.

Placental weight (PW) is an easily available measure often used in epidemiological studies^12,13^ as a proxy for placental growth and function. The close relationship between placental and fetal growth is reflected by a positive correlation (*r* = 0.6) between PW and birth weight (BW) in large cohorts^12,14^. Several genome-wide association studies (GWAS) have identified genetic loci in the maternal and fetal genomes associated with BW^15,16^, and BW–associated loci are enriched for placental expression quantitative trait loci (eQTLs)^17^. However, no GWAS of PW is available to date, and it remains unclear how the genetics of placental growth relate to the genetics of fetal growth or to the genetics of adverse pregnancy outcomes such as preeclampsia. While the placenta is primarily composed of cells with fetal origin, it is intricately connected to maternal tissues and circulation^18,19^. Genetic analyses offer the opportunity for insight into the complex interplay of direct fetal, indirect maternal, and parent-of-origin effects (POE), which we hypothesize underlie placental growth and function.

We conducted the first GWAS of PW in term, singleton pregnancies, meta-analyzing data from 28 European studies. Analyses of 19,861 child-mother-father trios with PW measurements enabled a better understanding of the relationship between fetal and maternal effects, including POE. We also categorized loci according to their association with BW, examined genetic links between PW and pregnancy, perinatal and later-life outcomes, and used Mendelian randomization (MR) to assess causal relationships between maternal and offspring characteristics and PW.

## Results

### Meta-analyses of fetal, maternal, and paternal GWASs

We performed GWAS meta-analyses of PW adjusted for fetal sex and gestational duration against the fetal (*n* = 65,405), maternal (*n* = 61,228), and paternal genomes (*n* = 52,392) (Fig. 1.). Methods and Supplementary Tables 1-6 provide information on data collection and genotyping. After data cleaning and imputation, 11 million SNPs were analyzed. The fetal GWAS meta-analysis identified 32 independent loci at *P* < 5 × 10^−8^, the maternal analysis identified four loci, and the paternal identified two loci (Fig. 2, Table 1, Supplementary Table 7). Approximate conditional and joint analysis (COJO) further identified secondary association signals at three fetal loci (Methods; Table 1; Supplementary Table 7). A comparison of effect sizes against minor allele frequencies for those 41 association signals was in line with expectations from statistical power (Supplementary Fig. 1). We also conducted analyses adjusted only for sex (*i*.*e*. not gestational age), which showed high genetic correlations with our main analysis results (fetal *r_g* = 1.00, SE = 0.001, *P* ≤ 1 × 10^−16^; maternal *r_g* = 0.99, SE = 0.001, *P* ≤ 1 × 10^−16^; paternal *r_g* = 1.00, SE = 0.004, *P* ≤ 1 × 10^−16^). Four additional loci reached genome-wide significance in the fetal sex-adjusted analyses, all of which were close to genome-wide significance in our main fetal analysis (Supplementary Table 8). Regional association plots for all genome-wide significant loci in the main analyses are shown in Supplementary Fig. 2a-e.

**Table 1:** Variants associated with PW (adjusted for sex and gestational duration) in the fetal, maternal, and paternal GWAS meta-analyses.

| Locus Name | Chr | Position (b37) | rsid | EA | OA | Effect Allele Frequency | Beta | SE | P | N | Maternal Fetal Classification | PW/BW Classification |
| --- | --- | --- | --- | --- | --- | --- | --- | --- | --- | --- | --- | --- |
| **Fetal** |  |  |  |  |  |  |  |  |  |  |  |  |
| DCST2 | 1 | 155001281 | rs150138294 | A | G | 0.053 | 0.080 | 0.013 | 1.07E-09 | 65405 | Unclassified | PW & BW same direction |
| RPL31P11 | 1 | 161651064 | rs723177 | T | C | 0.296 | 0.039 | 0.006 | 1.02E-10 | 65405 | Fetal | Predominantly or only PW |
| TSNAX-DISC1, LINC00582 | 1 | 231733795 | rs1655296 | G | T | 0.393 | 0.042 | 0.006 | 4.86E-14 | 65405 | Fetal | Predominantly or only PW |
| TSNAX-DISC1, LINC00582* | 1 | 231794081 | rs140691414 | T | C | 0.007 | 0.261 | 0.039 | 2.54E-11 | 57158 | Fetal | PW & BW same direction |
| CHRM3 | 1 | 239822859 | rs10925945 | T | C | 0.936 | 0.067 | 0.011 | 1.69E-09 | 65405 | Unclassified | Predominantly or only PW |
| EPAS1 | 2 | 46567276 | rs4953353 | G | T | 0.622 | 0.042 | 0.007 | 1.38E-10 | 48809 | Unclassified | PW & BW same direction |
| ADCY5 | 3 | 123065778 | rs11708067 | G | A | 0.233 | 0.053 | 0.007 | 1.83E-16 | 65405 | Fetal & Maternal | PW & BW same direction |
| LOC339894/CCNL1 | 3 | 156795414 | rs9817452 | G | T | 0.614 | 0.040 | 0.006 | 1.72E-12 | 65405 | Fetal | PW & BW same direction |
| PDLIM5 | 4 | 95531563 | rs74457440 | A | G | 0.233 | 0.036 | 0.007 | 3.37E-08 | 65405 | Unclassified | PW & BW same direction |
| ACTBL2 | 5 | 57073666 | rs7722058 | T | C | 0.845 | 0.047 | 0.008 | 6.06E-10 | 65405 | Fetal | PW & BW same direction |
| HSPA4 | 5 | 132444128 | rs72801474 | A | G | 0.105 | 0.055 | 0.009 | 7.34E-09 | 65405 | Fetal | Predominantly or only PW |
| ARHGAP26 | 5 | 142429811 | rs3822394 | C | A | 0.263 | 0.036 | 0.006 | 5.80E-09 | 65405 | Fetal | Predominantly or only PW |
| EBF1 | 5 | 158433339 | rs67265526 | T | C | 0.628 | 0.032 | 0.006 | 1.12E-08 | 65405 | Fetal | Predominantly or only PW |
| NUDT3 | 6 | 34237188 | rs541641049 | A | G | 0.018 | 0.193 | 0.035 | 4.52E-08 | 55263 | Fetal | PW & BW same direction |
| FKBP5/MAPK13/TEAD3 | 6 | 35517390 | rs9800506 | T | G | 0.410 | 0.033 | 0.006 | 2.58E-09 | 65405 | Unclassified | PW & BW same direction |
| HACE1 | 6 | 105130521 | rs12529634 | C | T | 0.872 | 0.056 | 0.008 | 2.85E-11 | 65405 | Fetal | PW & BW same direction |
| ESR1 | 6 | 152042413 | rs11756568 | A | T | 0.703 | 0.052 | 0.006 | 1.95E-18 | 65405 | Fetal | PW & BW same direction |
| PDE10A | 6 | 166182483 | rs1021508 | C | T | 0.633 | 0.034 | 0.006 | 2.09E-09 | 65405 | Fetal | PW & BW same direction |
| PDE10A* | 6 | 166199513 | rs6456014 | C | A | 0.582 | 0.030 | 0.006 | 4.42E-08 | 65405 | Fetal | Predominantly or only PW |
| ISPD | 7 | 16193877 | rs7783810 | C | T | 0.422 | 0.039 | 0.006 | 1.85E-12 | 65405 | Fetal | PW & BW same direction |
| TBX20 | 7 | 35282931 | rs10486660 | A | C | 0.608 | 0.040 | 0.006 | 9.49E-13 | 65405 | Fetal | PW & BW same direction |
| YKT6 | 7 | 44246271 | rs138715366 | C | T | 0.991 | 0.202 | 0.035 | 7.91E-09 | 63786 | Fetal & Maternal | PW & BW same direction |
| ENTPD4 | 8 | 23342043 | rs6557677 | T | A | 0.166 | 0.044 | 0.007 | 2.66E-09 | 65405 | Fetal | Predominantly or only PW |
| SLC45A4 | 8 | 142247979 | rs12543725 | G | A | 0.585 | 0.032 | 0.006 | 9.20E-09 | 65405 | Unclassified | PW & BW same direction |
| KLF4 | 9 | 110822658 | rs1434836 | A | G | 0.575 | 0.035 | 0.006 | 2.87E-10 | 65405 | Fetal | PW & BW same direction |
| ADRB1 | 10 | 115805056 | rs1801253 | C | G | 0.743 | 0.052 | 0.006 | 8.06E-17 | 65405 | Fetal | PW & BW same direction |
| KCNQ1 | 11 | 2839751 | rs2237892 | T | C | 0.056 | 0.077 | 0.014 | 1.03E-08 | 48809 | Fetal & Maternal | Predominantly or only PW |
| SERPINA1 | 14 | 94838142 | rs112635299 | T | G | 0.024 | 0.149 | 0.019 | 1.42E-15 | 64541 | Fetal | Predominantly or only PW |
| FES/FURIN | 15 | 91428636 | rs7177338 | A | G | 0.536 | 0.035 | 0.006 | 3.28E-10 | 65405 | Unclassified | PW & BW same direction |
| NR2F2 | 15 | 96852638 | rs55958435 | A | G | 0.713 | 0.042 | 0.006 | 6.82E-12 | 65405 | Fetal | PW & BW same direction |
| GPR139/GPRC5B | 16 | 20006986 | rs57790054 | G | A | 0.302 | 0.035 | 0.006 | 5.54E-09 | 65405 | Fetal | PW & BW same direction |
| SLC6A2 | 16 | 55717569 | rs11866404 | C | G | 0.533 | 0.042 | 0.006 | 8.55E-14 | 65405 | Fetal | Predominantly or only PW |
| SLC7A5 | 16 | 87882209 | rs876987 | G | A | 0.306 | 0.038 | 0.007 | 1.24E-08 | 48809 | Fetal | PW & BW opposite directions |
| LOC339593 | 20 | 11200008 | rs6040436 | T | C | 0.444 | 0.054 | 0.006 | 1.55E-22 | 65405 | Fetal | PW & BW same direction |
| LOC339593* | 20 | 11428113 | rs6078190 | C | A | 0.319 | 0.040 | 0.006 | 1.36E-11 | 65405 | Fetal | Predominantly or only PW |
| **Maternal** |  |  |  |  |  |  |  |  |  |  |  |  |
| EBF1 | 5 | 157808173 | rs72804545 | A | T | 0.888 | 0.064 | 0.011 | 2.23E-08 | 41211 | Maternal | PW & BW same direction |
| LMO1 | 11 | 8255408 | rs2168101 | C | A | 0.696 | 0.047 | 0.008 | 2.13E-09 | 41356 | Maternal | PW & BW same direction |
| SLC38A4 | 12 | 47180370 | rs180435 | C | G | 0.190 | 0.051 | 0.007 | 1.68E-12 | 61153 | Maternal | PW & BW same direction |
| NLRP13 | 19 | 56423893 | rs303998 | G | A | 0.404 | 0.039 | 0.006 | 2.12E-11 | 61063 | Maternal | PW & BW same direction |
| **Paternal** |  |  |  |  |  |  |  |  |  |  |  |  |
| EBF1 | 5 | 158317602 | rs75512885 | T | A | 0.925 | 0.080 | 0.015 | 4.96E-08 | 31714 | Unclassified | PW & BW same direction |
| LOC339593 | 20 | 11207949 | rs2207099 | A | G | 0.448 | 0.038 | 0.006 | 8.77E-10 | 52271 | Fetal | PW & BW same direction |

The lead SNP is given in the rsid column. Where PW loci fell within 500 kb of BW loci, locus names were given the name of the BW locus in Warrington et al.<sup>15</sup> otherwise the locus was named by the nearest gene. Secondary signals at a locus are marked with an asterisk. Effect alleles are coded to correspond to increasing PW. Beta and SEs are given in SD units of PW. The subheadings under Locus Name indicate the GWAS analysis that the signal was identified in, and two last columns classify signals into fetal and/or maternal mode of association (using a weighted linear model<sup>15</sup>) and into whether effects were only/predominantly on PW, or also on BW. Abbreviations: Chr, chromosome; rsid, reference SNP cluster ID; EA, effect allele; OA, other allele; SE, standard error; P, P value; N, number; BW, birth weight; PW, placental weight.

**Fig. 1:**
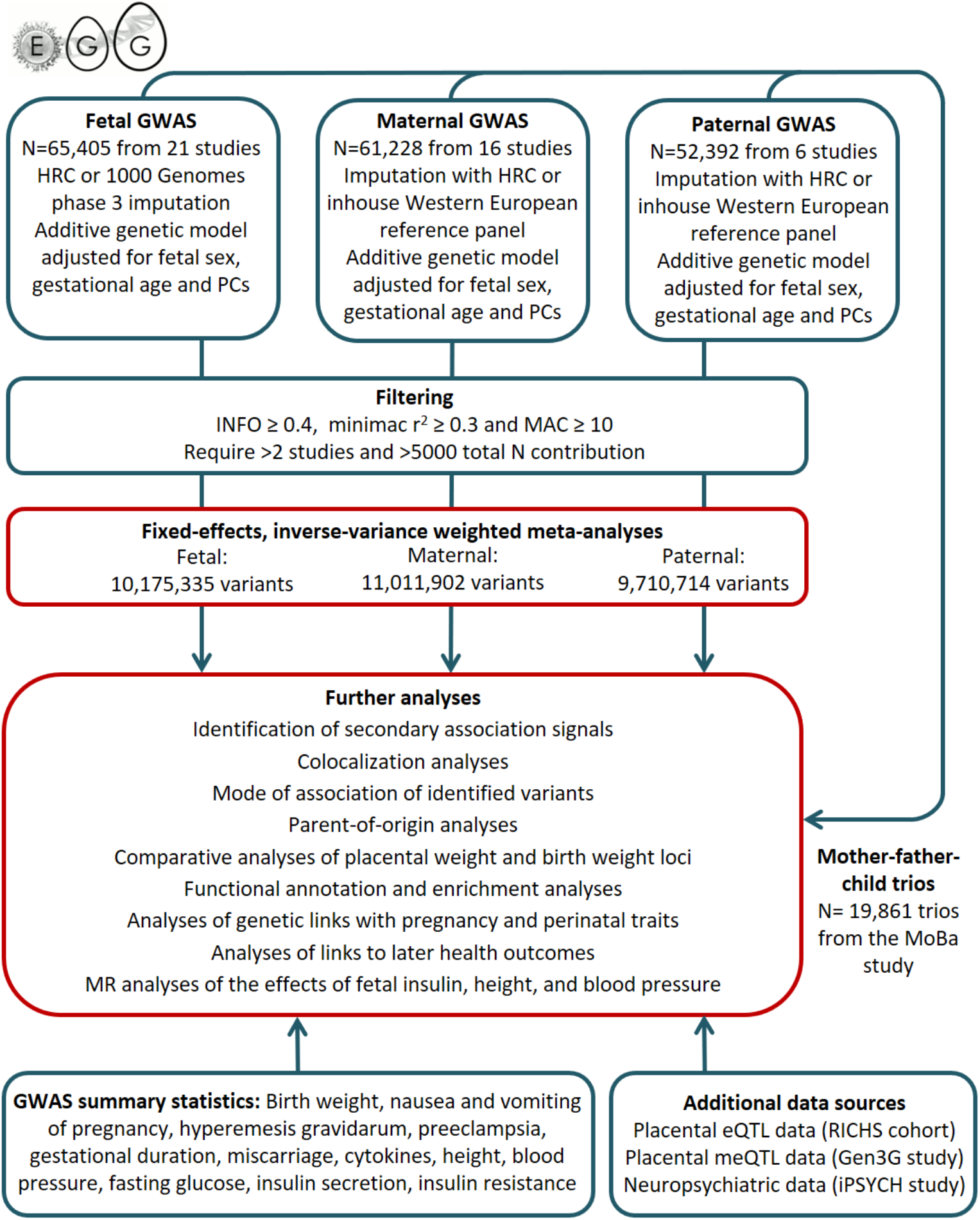
Flow chart of the study design. Abbreviations: eQTL, expression quantitative trait loci; GWAS, genome-wide association study; HRC, Haplotype Reference Consortium; MAC, minor allele content; meQTL: methylation quantitative trait loci; PC, principal component.

**Fig. 2:**
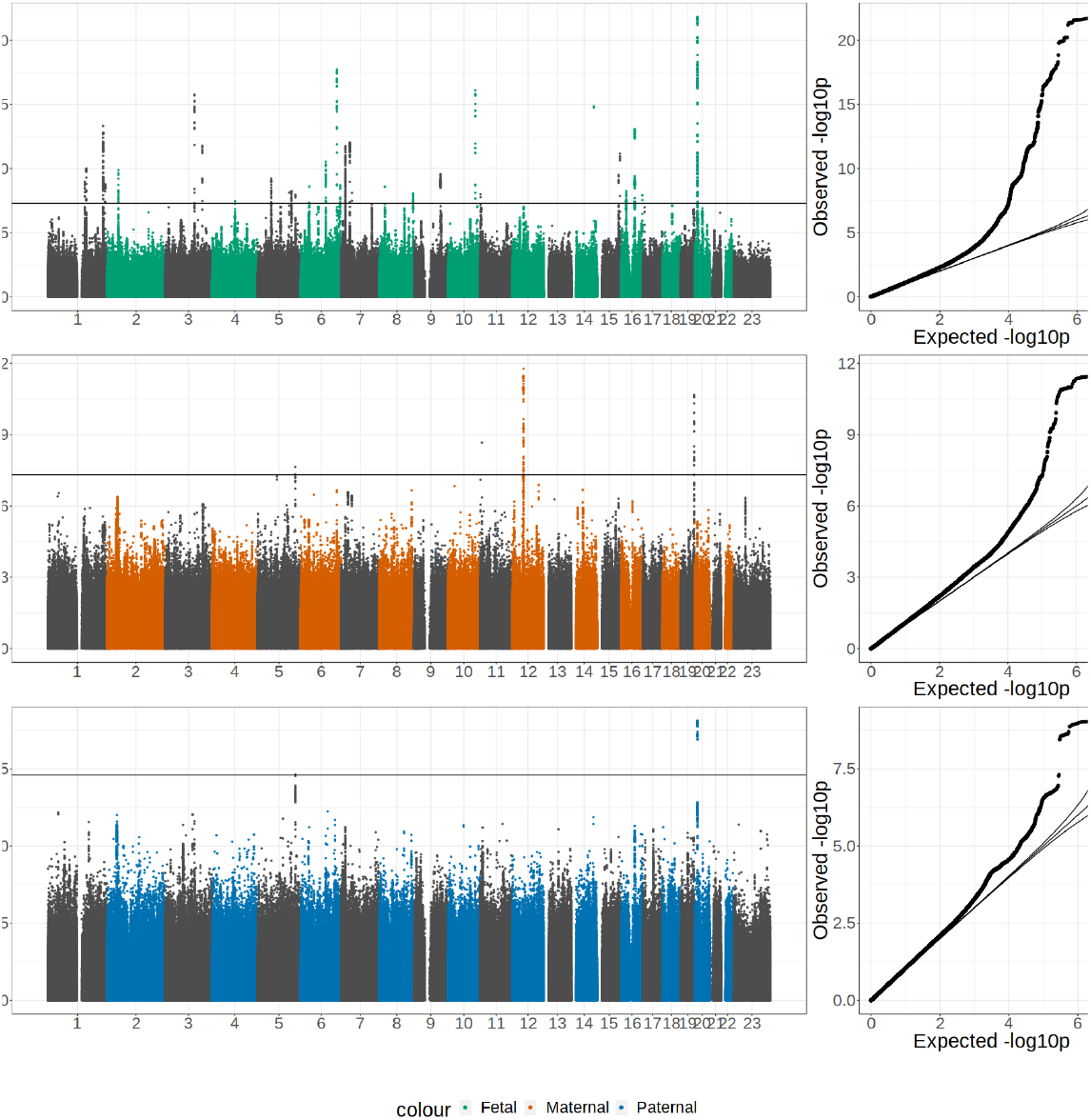
Genome-wide association results for PW. Manhattan plots of –log_10_ *P* values across the chromosomes and corresponding quantile–quantile plot of observed versus expected –log_10_ *P* values for meta-analyses of SNP associations with placental weight in **a**, the fetal GWAS; **b**, the maternal GWAS; and **c**, the paternal GWAS.

### Resolving fetal and parental contributions to association signals

Fetal genotype is correlated with both maternal and paternal genotype (each *r* = 0.5). While the fetal genotype may influence PW directly, the maternal genotype itself may have effect *via* the intrauterine environment. To calculate the SNP heritability of the maternal, paternal, and offspring contributions we used a framework within genomic structural equation modelling (SEM)^20^ developed by Moen et al.^21^ (Methods). We found evidence for substantial contributions from the fetal (h2=0.22 (SE=0.03)) and maternal (h2=0.12 (SE=0.02)) genomes to variation in placental weight, and a small component from the paternal genome (h2=0.06 (SE=0.02))(Supplementary Table 9, Supplementary Fig. 3). We also found genetic correlation between the three latent variables suggesting that the fetal effect on placental weight was negatively correlated with both maternal and paternal effects, while maternal and paternal effects on placental weight were positively correlated (Supplementary Table 9).

To estimate fetal-specific, maternal-specific, or paternal-specific effects on PW at each identified locus, we first applied a weighted linear model (WLM)^15^ to the GWAS meta-analysis summary statistics. The WLM can accurately approximate conditional effects (e.g. fetal effect conditional on any maternal effect, or vice versa) in absence of genotyped mother-child pairs by incorporating results from all samples, even those overlapping between the three GWAS meta-analyses (Methods), to maximize statistical power. We applied formulations of the WLM allowing for non-zero paternal effects, or fixing the paternal effects to zero (Supplementary Table 10). At the 41 association signals, we applied the classification method used for BW^15^ (Methods) to categorize the SNPs as having fetal or maternal effects. Hence, 26 SNPs were classified as fetal only, four as maternal only, two as fetal and maternal with effects in opposite directions, and one as fetal and maternal with effects in the same direction. We could not resolve the classification for the remaining eight loci (Supplementary Table 10, Supplementary Fig. 4a, Supplementary Fig.4b).

We complemented the WLM analysis with within-family analyses in the child-mother-father trio subset from the Norwegian Mother, Father, and Child Cohort (MoBa)^22^. First, the associations of fetal and parental genotypes with PW were conditioned against each other (*n* = 19,861 independent trios, Supplementary Table 10). There was good agreement between mode of association categories based on WLM results and those from conditional analyses in trios. Taking advantage of the availability of phased genotypes for MoBa children, we further decomposed the association signals into their mode of transmission (*i*.*e*., maternal transmitted, maternal non-transmitted, paternal transmitted, and paternal non-transmitted alleles). Since several loci were already known to be associated with BW (see below), we compared our results with a recent analysis of BW-mode of transmission^16^ (Supplementary Table 11). For loci associated with both traits, effect size estimates and mode of transmission were consistent between BW and PW analyses (Fig. 3a, Supplementary Fig. 5). Among the eight loci that remained unclassified after WLM analysis of PW, five were identified previously in BW GWAS and classified as fetal^16^, and one was classified as fetal and maternal with same direction of effect (Supplementary Table 11).

**Fig. 3:**
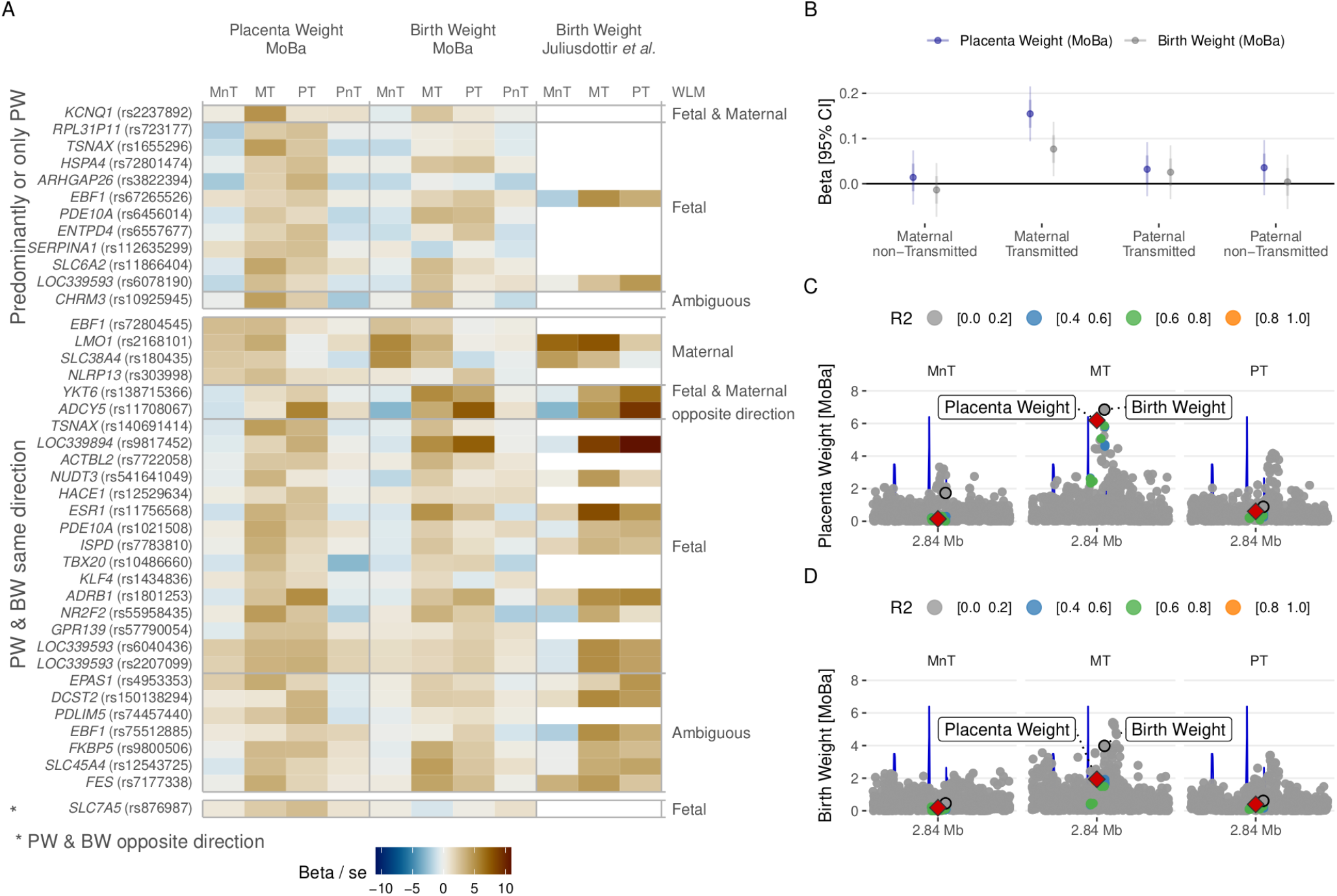
Resolving fetal and parental contributions to PW associations. **a**, Effect size estimates divided by standard error in the mode of association analyses for the 41 association signals and the association of the maternal non-transmitted, maternal transmitted, paternal transmitted, and paternal non-transmitted alleles with standardized PW and BW adjusted for sex and gestational duration in MoBa, and with BW according to Juliusdottir *et al*.^*16*^ (a blank value indicates that no proxy was found with *r*^*2*^ ≥ 0.2). **b**, Effect size estimates for standardized PW and BW adjusted for sex and gestational duration for rs2237892 near *KCNQ1* in MoBa, thin and thick error bars respectively represent 95% confidence intervals and one standard error on each side of the effect size estimate. **c**,**d**, Regional plots at the *KCNQ1* locus, plotting the negative base 10 logarithm of the unadjusted *P*-value against genomic location, for the maternal non-transmitted, maternal transmitted, paternal transmitted, and paternal non-transmitted alleles in their association with standardized placental (**c**) and BW (**d**) adjusted for sex and gestational duration in MoBa. The leading variants for PW and BW, rs2237892 and rs234864, are annotated with a diamond and a circle, respectively. Points are colored according to their LD with rs2237892. Abbreviations: MnT, maternal non-transmitted alleles; MT, maternal transmitted alleles; PT, paternal transmitted alleles; PnT, paternal non-transmitted alleles.

For the signal identified in the paternal GWAS near *EBF1* (rs75512885), our sample size did not allow resolving the mode of association using WLM or parent-offspring trio analyses, but this variant is in moderate LD with rs72813918 (*r*^*2*^ = 0.57), classified previously as fetal for BW^16^ (Supplementary Table 11). Colocalization analysis suggested that this signal for BW was independent of the nearby fetal association signal for PW (rs67265526; posterior probability for independent signals > 0.99). The lead SNP of the other paternal GWAS locus (rs2207099, near *LOC339593*) colocalized with the nearby fetal lead SNP (rs6040436; posterior probability for shared association = 0.97) (Supplementary Table 7), and was therefore excluded from subsequent analyses, leaving 40 independent association signals.

For all loci, we inspected the distance of the lead SNP to known imprinted genes according to www.geneimprint.com (Supplementary Table 11). Only one lead variant was found near imprinted genes; rs2237892 located in intron 10 of the imprinted gene *KCNQ1*. Indeed, the mode of transmission analysis in MoBa showed that the association with PW was only conferred by the maternally transmitted allele (Fig. 3b, Supplementary Fig. 5) consistent with the known maternal-only expression of *KCNQ1* and nearby *CDKN1* genes. This variant shows POE on type 2 diabetes risk^23^, where the maternally inherited risk allele (C) corresponds to the maternally inherited PW-decreasing allele identified here. Another nearby variant, rs234864, has been reported^16^ as a maternally-transmitted effect only for BW. The very low LD (*r*^*2*^ = 0.05) and very different minor allele frequencies (5.6% *vs*. 49.4%) between the two variants suggest that these may be independent signals, both imprinted (Fig. 3c). While the maternally transmitted allele of the BW variant rs234864 was strongly associated with both PW and BW in MoBa, the maternally transmitted allele of the PW variant rs2237892 showed directionally consistent, but weaker evidence of association with BW in MoBa (Fig. 3d). This is in agreement with the lower fetal effect size detected of rs234864 on BW in previous BW GWAS^16^ compared with the effect on PW (Supplementary Table 11).

### Correlations between PW and BW

The strong phenotypic correlation reported between PW and BW^12,14^ was confirmed in the MoBa samples in this study (Spearman’s *r* = 0.59, adjusted for sex and gestational duration). We applied LD score regression^24^ to estimate genetic correlations with published BW association summary statistics^15^, analyzing both main GWAS summary statistics, and WLM-adjusted estimates of the conditional fetal, maternal and paternal effects (Fig. 4). Using WLM-adjusted effects for both PW and BW, the fetal and maternal genetic correlations between PW and BW remained strong. We observed a negative genetic correlation between fetal-specific effects on BW and maternal-specific effects on PW (Fig. 4c), whereas fetal-specific effects on PW showed no genetic correlation with maternal-specific effects on BW (Fig. 4d). There was no genetic correlation between paternal-specific effects on PW and either maternal- or fetal-specific effects on BW after WLM-adjustment.

**Fig. 4:**
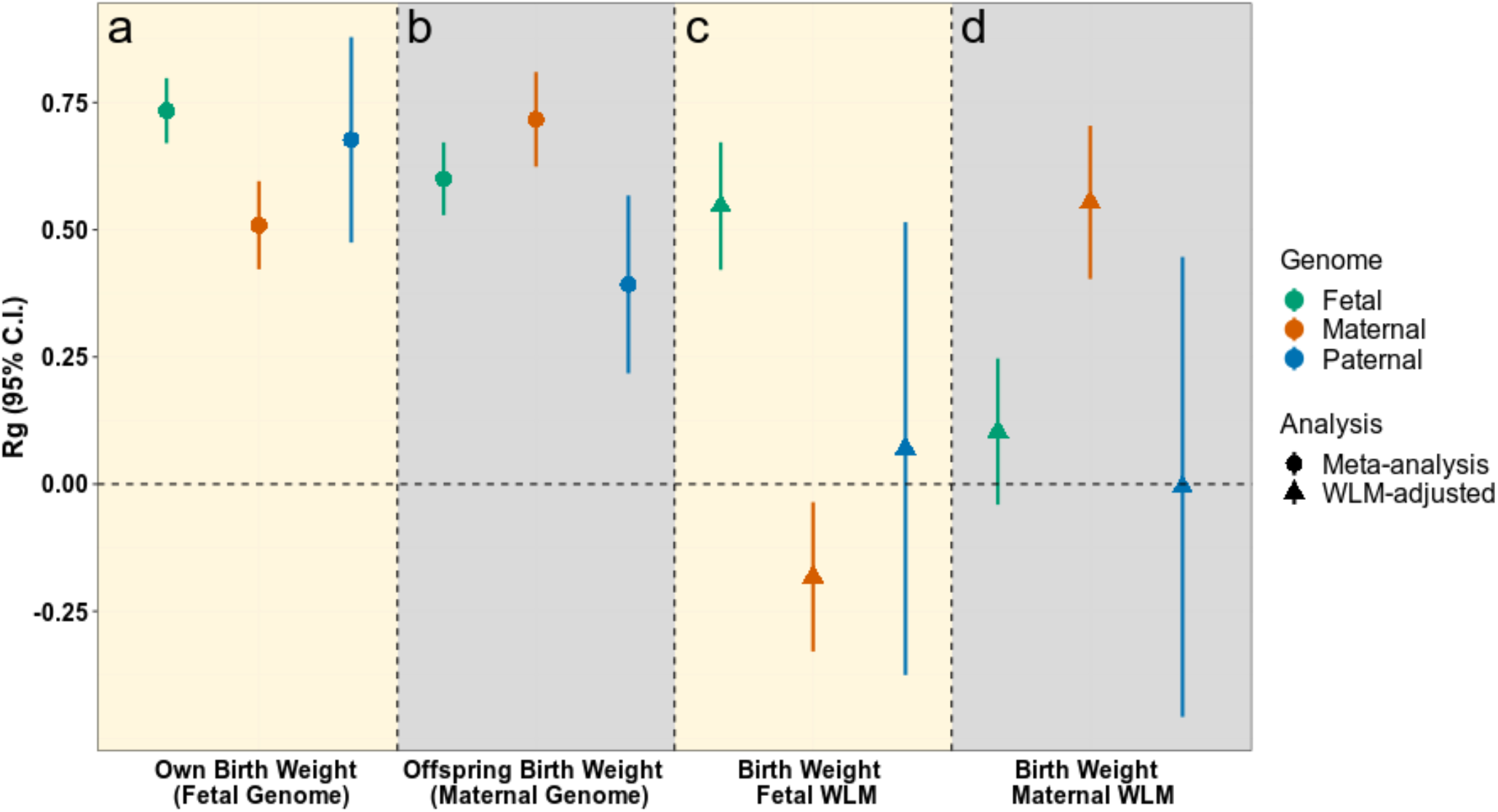
Genetic correlation between PW and BW. **a**, Genetic correlation estimates for PW (current study - illustrated by circles or triangles with 95% CI) and BW (using 4 types of estimate) from Warrington *et al*.^15^. “Own birth weight” means fetal GWAS of birthweight, and “offspring birth weight” means maternal GWAS of offspring birth weight. Values are provided for fetal, maternal, and paternal effects before and after WLM adjustment. Error bars represent 95% confidence intervals. Abbreviations: CI, confidence interval; Rg, genetic correlation; WLM, weighted linear model.

From the largest available GWAS of BW^16^, we selected BW loci within 500 kb of PW loci (Supplementary Table 7). Twenty-eight of the 40 independent PW signals were also reported for BW or had a BW lead SNP nearby. All these were within 100 kb of the PW lead SNP except rs876987 at *SLC7A5* (> 419 kb). Colocalization analysis suggested that 19 of these represent a shared underlying association (posterior probability for shared signal > 0.8), five signals were distinct (posterior probability for separate signals > 0.8), and the final four loci were uncertain (both posterior probabilities < 0.8).

Given the large proportion of signals colocalizing with BW loci, we aimed to classify loci showing evidence of associations with both PW and BW from those showing only (or predominantly) evidence of association with PW, by comparing PW and BW effect estimates (Supplementary Table 7, Methods). Twelve signals were classified as only or predominantly PW, and the remaining 28 were classified as both PW and BW signals, with one of these (near *SLC7A5*) showing opposite directions of effect both with *P* < 0.05. The results obtained from WLM-adjusted analysis were largely consistent (Fig. 5, Supplementary Fig. 6, Supplementary Table 7). Thus, 12 of the 40 independent signals only influence PW, or have effects on PW that are larger than any estimated effect on BW (Table 1).

**Fig. 5:**
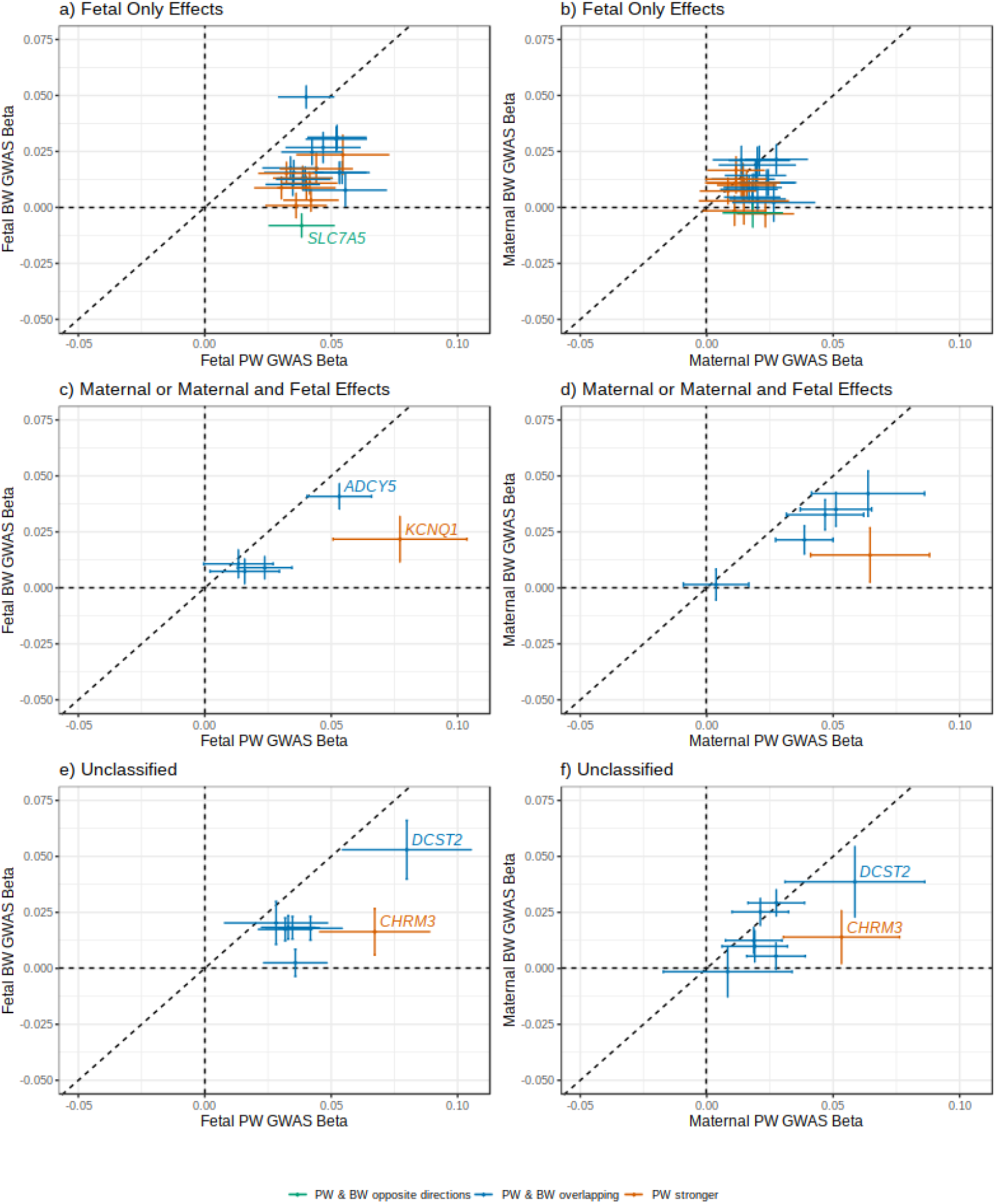
Scatter plots comparing effect sizes from PW and BW GWAS for placental weight SNPs. Scatter plots comparing effect size estimates from the PW GWAS with those from the BW GWAS^15^. The left column shows fetal genome associations, and the right shows maternal. The top row shows SNPs classified as fetal only effects on PW (Supplementary Table 7). The middle row shows SNPs classified as maternal, or maternal and fetal, and the bottom row shows unclassified SNPs. SNPs with minor allele frequency below 5% have been excluded to improve resolution. The full figure including the lower frequency SNPs is shown in Supplementary Fig. 6. Colors indicate the classifications, which are given in the key below the figure. Abbreviations: BW, birth weight; GWAS, genome-wide association study; PW, placental weight. Error bars represent 95% confidence intervals.

### Connections to placental development and morphology, and transport of antibodies and amino acids

To prioritize key tissues and cell types relevant to fetal genes implicated by our identified loci, we selected the 31 protein-coding genes closest to the index SNP of the 32 loci identified in the fetal GWAS (i.e. excluding *LOC339593*; Table 1) and examined their expression using tissue-specific mRNA abundance data from the Human Protein Atlas and a scRNA-seq data set of over 70,000 single cells at the decidual-placental interface in early pregnancy (Methods). Among 61 tissues, the 31 genes ranked higher in terms of mRNA abundance than other genes in placenta (*P* = 1.8 × 10^−4^) (Supplementary Fig. 7, Supplementary Table 12). In the single cell analysis among the 32 different cell types at the early maternal–fetal interface, expression of the 31 genes ranked significantly higher than other genes in cell types of fetal origin, including fetal endothelial cells (*P* = 6.2 × 10^−5^) and syncytiotrophoblasts (*P* = 1.9 × 10^−4^), but they also ranked higher in maternal innate lymphocyte cells (*P* = 1.2 × 10^−3^) (Supplementary Fig. 8, Supplementary Table 12).

To further investigate associations between PW loci and gene expression or methylation in placental tissue, we queried lead PW SNPs (and their proxies *r*^*2*^ ≥ 0.8) against eQTL data from placenta in the Rhode Island Child Health Study dataset^25^. Lead PW SNPs tagged placental eQTL at four loci (*HSPA4, TBX20, SLC7A5, JAG1*) (Supplementary Table 7). We additionally investigated whether any of the 40 independent lead SNPs were associated with placental DNA methylation at CpG sites within 0.5 Mb. Using Gen3G data^26,27^, we identified placental meQTLs at 21 of the 40 independent signals (FDR ≤ 0.05 extrapolated from the genome-wide beta distribution by TensorQTL; Supplementary Fig. 8). Among the 21 lead SNPs with identified meQTLs, individual SNPs were associated with placental DNA methylation at up to 15 CpG sites (Supplementary Fig. 9). Supplementary Table 7 shows meQTLs with lowest *P*-value for each lead SNP.

We combined the placental expression and methylation data with further look-ups of lead PW SNPs in available GWAS of gene expression, plasma protein levels, diseases and traits (Supplementary Tables 7, 13, 14, 15). These searches implicated several candidate genes and potential biological insights. For example, the PW-raising allele of rs723177 at the fetal *RPL31P11* locus (Supplementary Fig. 2a) is an eQTL and pQTL for *FCGR2A* and *FCGR2B* (receptors for the Fc region of IgG complexes) and associates with higher plasma protein levels (Supplementary Tables 14, 15), suggesting a potential role in maternal antibody transfer across the placenta^28^. The lead variant rs723177 is also the strongest placental meQTL in the region at the CpG site cg27514565 (overlapping DNase I Hypersensitive Sites [DHS] for primitive/embryonic and myeloid/erythroid tissues) located between *FCGR2C* and *FCGR3B*, and two meQTL (at cg14354529 and cg15531363) overlapping with trophoblast-specific DHS located downstream of *FCRLA* and *FCRLB* (Supplementary Fig. 6). Associations at lead SNP rs10486660, at another fetal locus, suggest a role for this locus in placental morphology: the SNP is intragenic in the T-box transcription factor gene, *TBX20* (Supplementary Fig. 2a) and marks an eQTL for *TBX20* in placenta (Supplementary Table 7). Approximately 50% of *TBX20* placental expression is found in trophoblast cells^29^, and *TBX20*-null mice are likely to show abnormal placental morphology compared to wild-type mice^30^. Third, the lead SNP rs112635299 is in LD (*r*^*2*^ = 1.0) with rs28929474, a missense variant in the serpin peptidase inhibitor gene *SERPINA1* (Supplementary Fig. 2a). The SNP is associated with several traits, including levels of >20 circulating plasma proteins^31^ (Supplementary Table 15) and the cause of autosomal recessive alpha-1 antitrypsin deficiency^32^. Within the placenta, *SERPINA1* expression regulates expression of inflammatory cytokines and serine protease *HTRA1*-induced trophoblast invasion through induction of endoplasmic reticulum stress^31^. Finally, lead fetal SNP, rs876987, being the only variant showing an inverse effect on PW relative to BW, is intragenic in *SLC7A5*, which encodes solute carrier family 7 member 5 (Supplementary Fig. 2a). The heterodimer encoded by *SLC7A5* and *SLC3A2* is a sodium-independent, high-affinity amino acid exchanger over membranes of several organs and is responsible for uptake of essential amino acids in placenta. Lead variant rs876987 is an eQTL for *SLC7A5* in placenta and a meQTL for 15 CpG sites, with the strongest association detected at cg03408354 (*P* = 4.7 × 10^−49^; located in intron 7 of *SLC7A5*) (Supplementary Table 7; Supplementary Fig. 9d).

### PW and pregnancy complications including preeclampsia

We tested whether the 40 independent lead SNPs were associated with pregnancy and perinatal traits, using published GWAS summary statistics for nausea and vomiting of pregnancy or hyperemesis gravidarum^33^, preeclampsia^34^, gestational duration^35^, miscarriage (recurrent and spontaneous)^36^, and ten cytokines assayed from neonatal blood spots^37^. The SNPs showed more associations with nausea and vomiting of pregnancy (maternal genotype effects), and with gestational duration, preeclampsia, and neonatal IgA levels (all fetal genotype effects) than expected under the null distribution (Supplementary Fig. 10, Supplementary Table 16)^35,36^. Scatter plots of effect sizes (Supplementary Fig. 11) suggested that fetal alleles predisposing to higher PW tended to associate with higher odds of preeclampsia and a shorter gestational duration. To formally test the effect of fetal PW raising alleles on preeclampsia and gestational duration, we performed Mendelian randomization (MR) analyses. These analyses showed that fetal genetic predisposition to a higher PW raises the risk of preeclampsia (OR 1.72 [95% CI: 1.19 - 2.47] per 1 SD higher fetal genetically predicted PW, *P* = 6 × 10^−3^), and shortens gestational duration (1.9 days [95% CI: 0.74 - 3.12] shorter per 1 SD higher fetal genetically predicted PW, *P* = 3 × 10^−3^) (Supplementary Table 17, Supplementary Fig. 12). We were unable to test causal relationships between fetal genetically predicted PW and nausea and vomiting of pregnancy or IgA, because only maternal effect estimates were available for the nausea and vomiting of pregnancy outcome^33^ and only unadjusted fetal effect estimates for IgA^37^. The gestational duration results were similar to those previously seen for BW^38^, where fetal genetic predisposition to higher BW is associated with shorter gestation, implying a general effect of fetoplacental growth^39^. However, known BW-associated SNPs^15^ with fetal only effects showed little evidence of association in the fetal GWAS of preeclampsia (Supplementary Fig. 9t), contrary to PW-associated SNPs with fetal only effects (Supplementary Fig. 9u). MR analysis did not support a causal relationship between fetal genetic predisposition to higher BW and odds of preeclampsia (*P* = 0.6; Supplementary Table 17, Supplementary Fig. 12), suggesting that the increase in preeclampsia risk is more specific to placental growth.

### Maternal and fetal characteristics and their effects on PW

We selected key maternal and fetal traits known to influence BW^15,16^ and used genetic instruments to assess whether there were similar causal relationships with PW.

#### Maternal glucose, fetal insulin and PW

Fetal insulin is a major determinant of fetal growth. Lower fetal insulin secretion due to rare fetal *GCK* mutations lead to reduced BW and PW compared with siblings without mutations^40,41^. To investigate the role of fetal insulin on PW in the general population, we used three SNP-sets in MR analyses: (i) 33 fasting glucose SNPs (maternal glucose crosses the placenta stimulating fetal insulin secretion; (ii) 18 insulin disposition index SNPs (estimated by insulin secretion multiplied by insulin sensitivity commonly used as a proxy for ß-cell function); and (iii) 53 BMI-adjusted fasting insulin SNPs (insulin resistance proxy in adults) (Supplementary Table 18)^15^. We used WLM estimates to ensure that maternal effects were adjusted for fetal effects, and vice versa. A genetic instrument representing 1 SD (0.4 mmol/l) higher maternal fasting glucose level was associated with a 47.4-g (95% CI: 23.7 - 71.2 g) higher placental weight (*P* = 4.46 × 10^−4^; Fig. 6; Supplementary Table 18). In addition, a 1-SD genetically higher fetal disposition index was associated with a 13.9-g (1.2 - 26.6) higher PW. Alleles that raise insulin secretion tend to lower glucose levels, and this was reflected in the opposite direction of the effect estimates for maternal disposition index (Fig. 6; Supplementary Table 18). The causal effect estimate for fetal insulin resistance was directionally consistent with causing lower PW, but the 95% CI crossed the null. These analyses support a role for insulin produced by the fetus, either directly or indirectly, in regulating growth of the placenta, providing a key link between fetal insulin and placental growth.

**Fig. 6:**
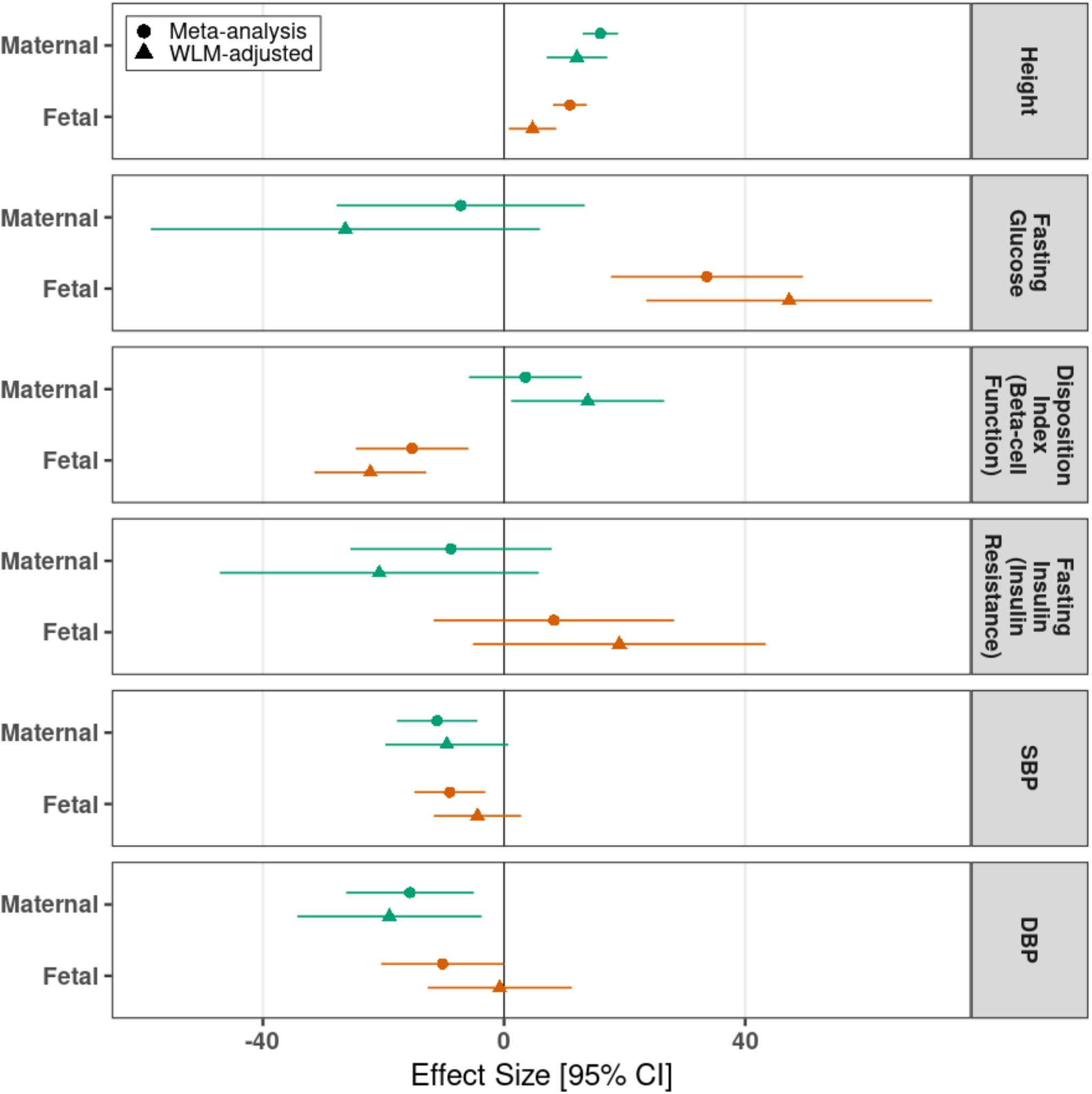
MR analyses testing effects of maternal exposures (and fetal genetic predisposition to those exposures) on PW. Results of two-sample MR analyses estimating the causal effects of maternal height, glycemic traits and blood pressure on PW, and the effects of fetal genetic predisposition to the same traits on PW. Maternal WLMs are adjusted for effects of the fetal genotypes, and fetal WLM analyses are adjusted for the effects of the maternal genotypes. Abbreviations: DBP, diastolic blood pressure; meta, meta analysis; SBP, systolic blood pressure; WLM, weighted linear model. Units for exposures are: height (SD), fasting glucose (SD), disposition index (SD), fasting insulin (SD), SBP (10 mmHg), DBP (10 mmHg). Causal estimates are g of placental weight per unit exposure.

#### Height and PW

Our analyses showed that both fetal and maternal genetic predisposition to a greater adult height were associated with greater PW, consistent with previous associations with higher BW and length^15,16^. A 1-SD (approx. 6 cm) higher genetically predicted adult height in the fetus was associated with a 12.1-g (95% CI: 7.1 - 17.2) higher PW (*P* = 2.88 × 10^−6^; Fig. 6; Supplementary Table 18) and a 1-SD higher maternal height was associated with a 4.7-g (95% CI: 0.8 - 8.7) higher PW, independent of direct fetal effects (*P* = 1.89 × 10^−2^; Fig. 6; Supplementary Table 18). These findings further emphasize key contributions of common fetal genetic factors to the close relationship between fetal and placental growth.

#### Blood pressure and PW

Higher maternal blood pressure in pregnancy is associated with reduced fetal growth^42^. To estimate the causal effect of maternal SBP or DBP on PW (adjusting for any direct fetal genetic effects), we used 68 known systolic blood pressure (SBP)-associated variants and 71 known diastolic blood pressure (DBP)-associated variants in two-sample MR analyses. We found no evidence of a causal effect of either maternal SBP (*P* = 0.23) or maternal DBP (*P* = 0.91; Fig. 6; Supplementary Table 18). However, analyses using direct fetal genotype effects (adjusted for maternal genotype effects) suggested that a 10-mmHg genetically higher DBP in the fetus caused a 19.0-g (95% CI: 3.8 - 34.3 g) lower PW, with a weaker effect of SBP in the same direction (9.5 g [95% CI: 0.5 - 19.7 g]; Fig. 6; Supplementary Table 18). Previous MR analyses of fetal blood pressure effects on BW have been inconsistent: those that used similar methods^15,43^ to the current analyses found no fetal effects, while others using transmitted and non-transmitted alleles^16,38^ supported fetal genetic predisposition to higher SBP being causally related to lower BW. Our findings for PW are similar to the latter.

### PW and later neuropsychiatric traits

The role of placenta in neurodevelopment and risk of later psychiatric disease in the offspring is a growing research field. Using data from the Initiative for Integrative Psychiatric Research (iPSYCH) cohort (*n* = 100,094), we analyzed observed fetal polygenic scores (PGS) for PW in groups of individuals who later developed neuropsychiatric disease. As expected, higher deciles in the PGS distribution corresponded to higher observed PW in a population representative sample, and we also observed similar patterns of association with PW in groups of individuals with autism spectrum disorder (ASD), attention deficit/hyperactivity disorder (ADHD), affective disorders, and schizophrenia (Supplementary Fig. 13). Logistic regression of each disease on fetal PGS for PW did not show any association, although there was a nominally significant association between higher observed PW and reduced risk of ADHD (*P* = 0.0247; Supplementary Fig. 14).

## Discussion

In this first GWAS of PW, we identify 40 independent association signals. These partially overlap with known BW loci, but 12 PW signals are related predominantly or only to PW, with connections to placental development and function. We observe a maternal POE near *KCNQ1*. Moreover, we find that fetal genetically mediated higher PW raises preeclampsia risk and shortens gestational duration, and we demonstrate a role for fetal insulin in regulating placental growth. Fig. 7 summarizes central results of the study.

**Fig. 7:**
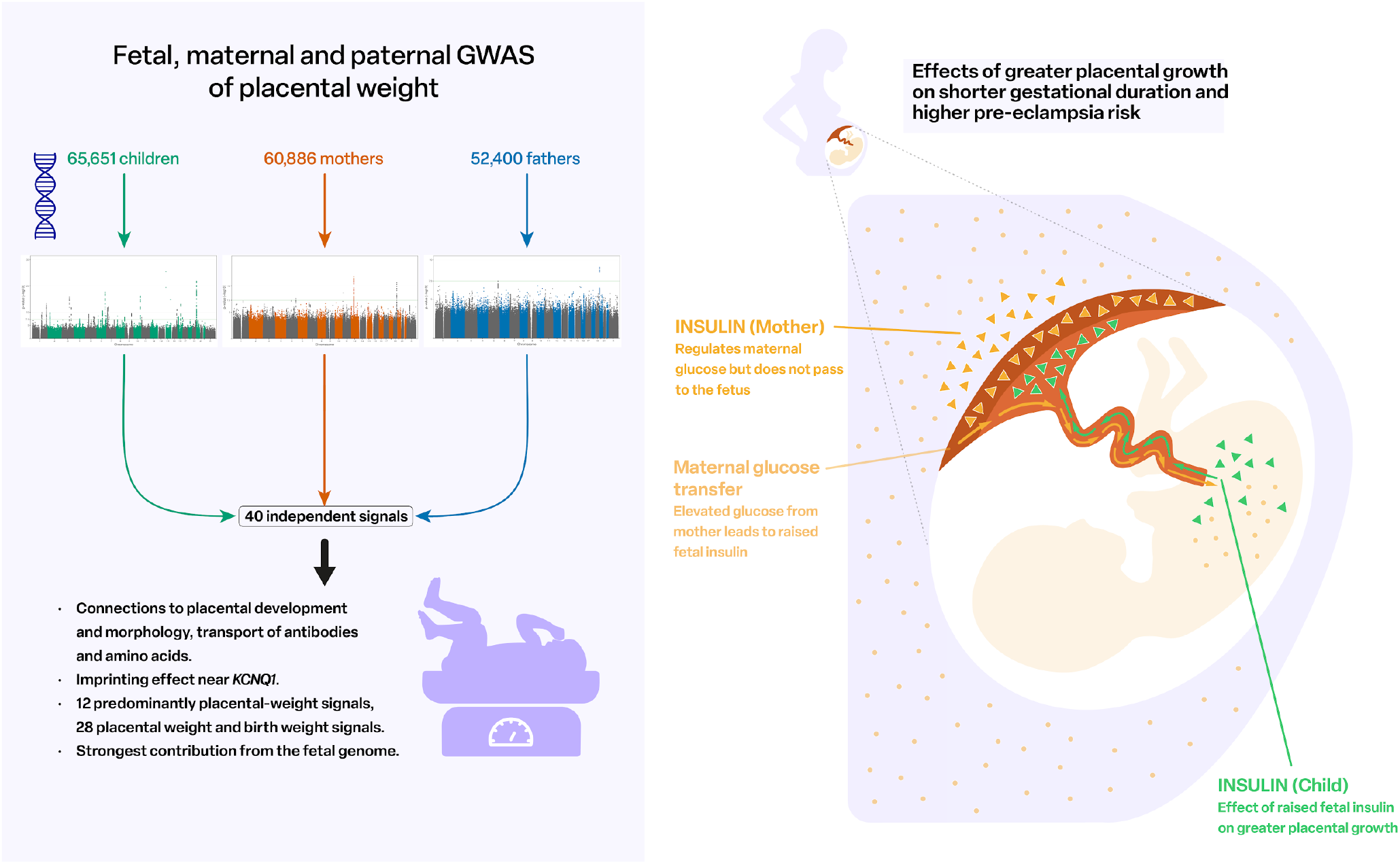
Overview of main study results. A salient feature of this study was the ability to address mode of association questions through analyses of fetal and parental genomes, application of WLM approaches, and analyses of almost 20,000 child-mother-father trios from the MoBa study. There was a clear predominance of fetal effect signals among the genome-wide significant loci, and fetal SNP heritability was almost double that of maternal SNP heritability (Supplementary Fig. 5). A total of 26 signals were classified as fetal only, and a further 3 as fetal and maternal. However, there were 4 loci that showed no fetal effect, and instead represent indirect maternal genetic effects acting on PW via the intrauterine environment. For loci also known to be associated with BW, our mode of association results were in good agreement with a previous classification^16^.

Investigations using MoBa trio data found a pronounced POE signal near *KCNQ1* carried by the maternally transmitted allele, in agreement with previously observed silencing of the paternal alleles of *KCNQ1* and the nearby *CDKN1C* gene. Notably, based on WLM analysis, we had classified the *KCNQ1* locus as having maternal and fetal effects in the same direction. Although the WLM may improve power to distinguish between maternal and fetal effects in available samples, family studies are still key to attaining clarity about how associations with neonatal phenotypes may arise, through fetal effects, POEs, maternal effects, or a combination. The *KCNQ1* finding has implications in fetal and placental growth through at least two pathways^44^. Placenta cells have a unique epigenetic profile that regulates its transcription patterns that may be associated with adverse pregnancy outcomes if disturbed. Beckwith-Wiedemann syndrome is an overgrowth syndrome with elevated PW and BW^44^. Approximately 50% of these patients have lost maternal-specific methylation at the putative Imprinting Control Region 2 (*ICR2*) within intron 10 of *KCNQ1* that is also the transcriptional start site of the regulatory long non-coding RNA *KCNQ1OT1*^*45*^ controlling expression of *CDKN1C* and *KCNQ1*. The location of the lead signal, rs2237892, to this region suggests it influences PW through an effect on methylation at *ICR2* in the placenta. Known associations between SNP rs2237892 and altered insulin secretion^46,47^ support a role for fetal insulin in mediating the association with PW. Additionally, placental DNA methylation at *KCNQ1* has been associated with maternal insulin sensitivity^48^, influencing maternal glucose availability across the placenta, and stimulating fetal insulin thus leading to fetal and placental growth.

Our results support a role for fetal genetic factors that predispose to a higher PW in raising preeclampsia risk: for every 1-SD higher genetically mediated PW, there was a more than 1.5-fold higher odds of preeclampsia. Preeclampsia is a complication that occurs in 3-5% of pregnancies^4,49^, but is poorly understood. The causal relationship we observed appears specific to placental growth as there was no similar effect of fetal genetic predisposition to higher BW. This finding is initially counter-intuitive, considering that preeclampsia is often linked with fetal growth restriction. However, the relationship between preeclampsia and placental weight is not consistent: early-onset preeclampsia, which is likely caused by defective placentation, is frequently associated with fetal growth restriction and low placental weight^50,51^, while the more common, “late-onset” or “term” preeclampsia (delivery in or after gestational week 37) has been associated observationally with both lower and higher PW^51^. Late-onset preeclampsia is thought to result largely from interactions between aging of the placenta and a maternal genetic susceptibility to cardiometabolic diseases^50^, but there is evidence that fetal trophoblast function is also important: the common genetic contributor to preeclampsia at the *FLT1* locus (lead SNP rs4769613) is fetal. Fetal *FLT1* likely mediates maternal endothelial dysfunction, but the variant was not associated with PW in our study, or with BW (*P* > 0.05)^15^. Our finding is supportive of a role for increased fetoplacental demands, which may result in uteroplacental mismatch, leading to late-onset preeclampsia^4^ and supports the role of fetally-mediated placental mechanisms in contributing to this condition.

Using PW as a proxy for placental growth had the advantage to reach the large sample size of this study, leveraging routine records in birth registers, and thereby yielding sufficient statistical power for discovery. One limitation, however, is that PW measured after delivery only crudely proxies placental growth and does not directly capture placental insults or other indications of placental dysfunction. Our study would therefore be complemented well by other approaches, e.g., involving single-cell transcriptomics of placental cells early in pregnancy^19^, characterization of placental mosaicism^52^, sequencing of cell-free RNA transcripts of placental origin in maternal circulation during pregnancy^53^, and RNA sequencing of placental samples without and with pregnancy complications^25,54^. Furthermore, while our results point to plausible candidate genes (e.g., *FCGR2A, FCGR2B, TBX20, SERPINA1, SLC7A5*) and potential effects on placental development and morphology, and transport of antibodies and amino acids, follow-up studies are required to establish causal links and to illuminate mechanisms related to placental growth.

Our study complements large-scale GWAS of BW. While genetic correlation and colocalization analyses revealed an overlap between signals associated with PW and BW, there were also notable differences. Twelve loci were classified as only or predominantly PW, and one signal showed opposite directions of effects between PW and BW, emphasizing the complexity of common and distinct genetic regulation and physiological processes affecting placental and fetal growth. Thus, our findings provide an improved understanding of the role of placenta for fetal growth and biological processes and complications such as gestational duration and preeclampsia. Future research may focus on additional proxies for placental function and environmental influences, as well as the role of placenta for later-life health outcomes.

## Methods

### Study cohorts and phenotype handling

Studies of individuals from European populations conducted genome-wide association analysis (GWAS) of placental weight as part of the EGG (Early Growth Genetics) Consortium. Where data was available, multiple births, congenital anomalies and babies born before 37 completed weeks of gestation or after 42 weeks and 6 days of gestation were excluded. Additionally, placental weight values < 200 g or > 1,500 g, or greater than five standard deviations from the mean were excluded. Placental weight was Z-score transformed for analysis. Details for phenotype exclusions, data collection, sample size, mean placental weight and gestational age for each cohort can be found in Supplementary Tables 1, 3, 5.

### Fetal, maternal, and paternal meta-analyses of PW GWAS

We performed separate GWAS to test associations between PW and fetal, maternal, and paternal genotypes, as detailed in equations (1) to (3).

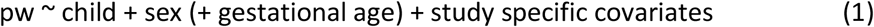

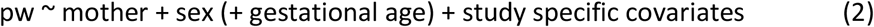

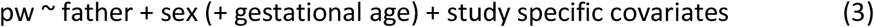

where ‘pw’ refers to the standardized PW, and ‘child’, ‘mother’ and ‘father’ refer to the number of tested alleles (as genotype probabilities) for a given variant in the child, mother, and father genomes, respectively.

Analyses were conducted twice, once adjusting for sex and gestational age and once adjusting for sex only. Adjustment was also made for ancestry principal components, and the number of principal components included was determined on a per-cohort basis. Genotypes were imputed to the HRC reference panel in most studies, with exceptions noted in Supplementary Tables 2, 4, 6. Details of imputation and analysis for individual studies are shown in Supplementary Tables 2, 4, 6. Association results from fetal, maternal, and paternal GWASs using both adjustment strategies were combined separately in fixed-effects meta-analyses implemented in METAL^55^, resulting in six meta-analyses. Meta-analyses were performed independently by two analysts. SNPs were excluded if they were present in fewer than two studies or the number of individuals for the SNP was < 5,000. Genome-wide significant loci were defined as regions with one or more SNP with *P* < 5 × 10^−8^, and these SNPs were defined as belonging to different loci if the distance between them was > 500 kb. The lead SNP at each locus was the one with the smallest *P*-value. Secondary signals within each locus were identified using approximate conditional and joint multiple-SNP (COJO) analysis performed using GCTA-COJO^56^. Independent SNPs were defined as those with conditional *P* < 5 × 10^−8^. The linkage disequilibrium reference panel was made up of 344,241 individuals from the UK Biobank defined as having British ancestry^57^.

### Genomic SEM

To calculate the SNP heritability of the maternal, paternal and offspring contributions we used a framework within genomic SEM^20^ developed by Moen et al^21^. In short the genomic SEM method^20^ involves two stages. In the first stage, LD score regression methods using pre-computed LD Scores from a European population provided by the original developers of LD score regression^24,58^ are applied to GWAS summary results statistics to estimate the genetic variance of each trait, and the genetic covariance between traits. In the second stage of genomic SEM, a user-defined SEM is fit to the genetic covariance matrix and parameters and their standard errors are estimated.

The genomic structural equation model we used to partition genetic covariances into maternal and offspring components is displayed in Supplementary Fig. 15. Results from the three placenta weight GWAS (squares) were modelled in terms of latent maternal, paternal and offspring genetic variables (circles). The lower part of this model reflects simple biometrical genetics principles (i.e. the fact that offspring and maternal genome are correlated 0.5) and consists of path coefficients fixed to the value one or one half. The top half of the model consists of free parameters requiring estimation - three SNP heritabilities (one for each trait), and three genetic covariances between the variables, representing commonalities in genetic action across the maternal, paternal and offspring genomes^21^.

It is important to realize that fitting a complicated SEM like the one in Supplementary Fig. 15 is necessary to obtain asymptotically unbiased estimates of SNP heritabilities and genetic correlations. The reason is that GWAS of perinatal traits represent a complicated mixture of maternal, paternal and offspring genetic effects. Our SEM disentangles these effects from each contributing GWAS. In contrast, the model underlying LD Score regression makes no allowance for this complication, and so naïve use will lead to biased estimates of SNP heritability and genetic correlations containing an unknown mixture of maternal, paternal and fetal effects^21^. Summary results statistics files from the GWASs described above were combined using genomic SEM^20^. The software was set to not exclude INDELs.

### Partitioning fetal, maternal, and paternal effects, and allele transmission analysis

Partitioning fetal, maternal, and paternal effects was performed from the summary statistics obtained from the GWAS (equations (1) to (3)) using a weighted linear model (WLM) similar to that applied in Warrington et al., extended to include paternal data and overlap in individuals between GWASs. The estimates of partitioned fetal, maternal, and paternal effects (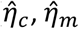, and 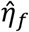 respectively), estimated from the GWAS estimates (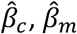, and 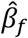 respectively) are

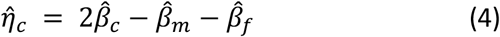

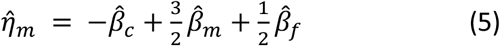

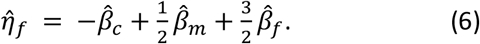

To account for sample overlap between GWASs, the covariance in estimates of the regression coefficients has the form

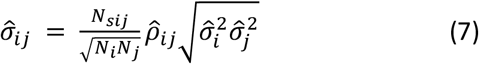

where *N*_*sij*_ is the number of individuals contributing to both analyses, *N*_*i*_ is the number of individuals contributing to analysis *i*, 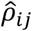 is the correlation between the estimates 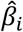 and 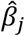 in the overlapping samples, and 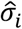 is the standard error of 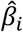. The term 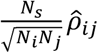 can be estimated as the intercept of a bivariate LDScore regression^24^. Standard errors for the partitioned effects are then estimated as

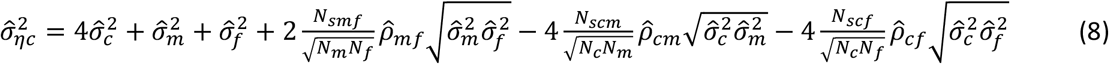

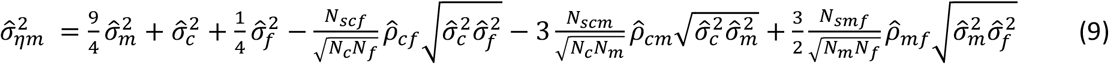

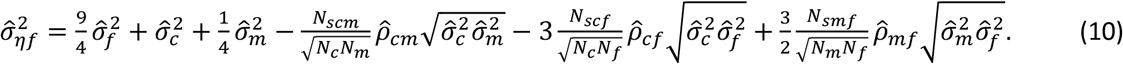

P-values are calculated from WLM estimates using a Z-test, with test statistic

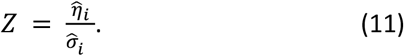

To complement this analysis, we utilized the independent child-mother-father trios available in MoBa to perform conditional analyses where the association of fetal and parental genotypes with PW are conditioned against each other, as detailed in equation (12).

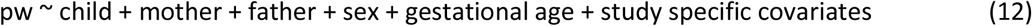

In this set of independent trios, using the phasing of the children’s genotypes, we inferred the parent-of-origin of the genotyped alleles as done by Chen *et al*.^*38*^. We then studied the association of the PW with maternal and paternal transmitted and non-transmitted alleles, as detailed in equation (13).

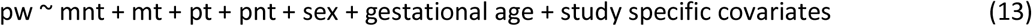

Where ‘mnt’ and ‘mt’ refer to the maternal non-transmitted and transmitted alleles, respectively; and similarly, ‘pt’ and ‘pnt’ refer to the paternal transmitted and non-transmitted alleles, respectively.

To estimate the significance of effects mediated by the maternal transmitted only, and hence test for parent-of-origin effects, we conditioned it on the genotypes of the child, mother, and father for the same variant, as detailed in equation (14).

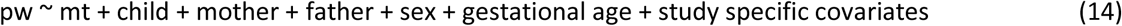

Where variables are coded as in equations (13) and (14). Note that trio and haplotype analyses in MoBa were conducted on hard-called genotypes. Sex, gestational age, and intercept were included in the model as well as study specific covariates, *i*.*e*., ten principal components and genotyping batches. The share of Mendelian errors was estimated using trios presenting at least a homozygous parent. If a variant presented more than 50 % Mendelian errors, children alleles were swapped. All estimates are provided in Supplementary Table 11.

To classify SNPs as either fetal, maternal or paternal, we used similar criteria to those used previously for BW SNPs^15^. We classified SNPs into three categories: (1) Fetal only: the P value for the fetal estimate is lower than a Bonferroni corrected threshold (0.05/37 = 0.00132), and the 95 % CI surrounding the estimate does not overlap the 95 % CI for the maternal estimate; (2) Maternal only: the P value for the maternal estimate is lower than a Bonferroni corrected threshold (0.00132), and the 95 % CI surrounding the estimate does not overlap the 95 % CI for the fetal estimate; and (3) Maternal and Fetal: the P value for both the maternal and fetal association estimates were less than the Bonferroni corrected threshold (0.00132). If a SNP did not fit any of these criteria it was marked as “unclassified”.

### Assessing colocalization of fetal, maternal or paternal association signals

To further supplement the aforementioned classifications, we performed colocalization analysis to determine overlap between GWAS signals from fetal, maternal, and paternal meta-analyses using the R library ‘coloc’^59^ with the default prior probability for colocalization. Signals were defined as colocalising if the posterior probability of shared association signals (P4) was > 0.8, distinct if the posterior probability of independent signals (P3) was > 0.8, and undetermined if neither P3 or P4 was > 0.8.

### LD score regression

To estimate the genetic correlation between PW and BW, linkage disequilibrium score regression^24,58^ was performed using summary statistics from gestational age and fetal sex adjusted analyses and BW summary statistics taken from Warrington *et al*^*15*^. Estimates for the results from the meta-analyses and WLM for both PW and BW were calculated for each of the fetal, maternal and paternal genomes.

### Testing whether lead PW SNPs were also associated with BW

Colocalization analysis was again utilized to determine whether PW loci close to those previously identified in GWAS ofBW represent the same association signal, using the same colocalization methods as above.

To classify SNPs as placental weight or BW, we calculated the 95 % CI in SD units for the PW lead SNP for both PW and BW. We then compared the 95% CIs; if these 95% CIs did not overlap the SNP was classified as “predominantly or only PW” unless either BW association (fetal or maternal) was in the opposite direction to the PW association and its associated P value was < 0.05, in which case it was classified as PW and BW in opposite directions. SNPs whose 95% CIs for PW and BW overlapped were classified as “PW and BW same direction”.

### Enrichment analysis using tissue-specific mRNA expression and scRNA data

We tested enrichment of expression of the PW associated genes in specific tissues or cell-types by comparing the ranks of gene expression across different tissue or cell-types (Supplementary Fig. 16). This method involved two steps: *first*, the tissue or cell-type specific expression levels from a reference expression data set were rank normalized across all tissues or cell-types for each gene; *second*, for a particular tissue or cell-type, expression enrichment of the test genes were compared against all other genes by the Wilcoxon rank-sum test. We used 31 protein-coding genes close to the index SNPs of the 32 PW-associated loci identified in the fetal GWAS (First section of Table 1; the *LOC339593* locus was not included because no protein-coding gene nearby). For the enrichment analysis in specific tissues, we used tissue-specific mRNA expression data from the Human Protein Atlas (RNA consensus tissue gene data)^29^. For the enrichment analysis in different cell-types, we used the scRNA-seq data of about 70,000 single cells at the decidual-placental interface in early pregnancy^19^. Cell-type specific gene expression of each gene was evaluated by percentage of cells with detectable scRNA reads. Significance levels for this enrichment analysis were Bonferroni corrected by number of tissues (*n* = 61) or cell-types (*n* = 32).

### Identification of placental meQTL

Using the 41 SNPs identified in the discovery meta-analyses GWASes of PW, we conducted placental methylation QTL (methQTL) analysis in 395 participants with European ancestry from the Canadian cohort of the study ‘Genetics of Glucose Regulation in Gestation and Growth’ (Gen3G)^26,27^. We measured DNA methylation using Illumina EPIC arrays from samples collected in the fetal-facing side of placenta. We conducted DNA methylation data processing, normalization, and cell type composition estimation as previously described^26^. Whole genome sequencing data were processed with the DNA-Seq v3.1.4 pipeline from GenPipes^60^ based on BWA_mem and GATK best practices to identify high quality SNPs on GRCh37 through joint genotyping over all samples. SNPs with a call rate above 20%, Hardy-Weinberg equilibrium p-value above 1e-6 and minor allele counts above 10 were included.

Methylation QTL were identified using beta values of 681 795 CpGs with TensorQTL^61^ in a cis-window of 0.5 Mb and the following covariates: four genotype PCs, fetal sex and cell type composition (estimates based on DNA methylation). In order to correct for the multiple CpGs tested genome-wide, a FDR threshold of ≤ 0.05 was applied on the q-values measured from the p-values extrapolated from the beta distribution by TensorQTL.

### Mapping to pathways and traits

Variants classified as PW through the SNP classification of PW or BW, were investigated further to identify relevant pathways and related biology. Systematic look-ups using Open Targets Platform, The Human Protein Atlas, and International Mouse Phenotyping Consortium. Each variant was assessed for which gene it is functionally implicated in through information gained regarding the variant’s involvement in molecular phenotype experiments, chromatin interaction experiments, *in silico* function predictions and distance between the variant and the gene’s canonical transcription start site. The most likely candidate gene was then used to investigate the placenta relevant cells that the gene is expressed in using information on the Open Targets Platform and The Human Protein Atlas websites. The international Mouse Phenotyping Consortium website was used to identify mouse models that found associations with placental pathology after removal of the gene in single-gene knockout mice. Finally, further information was garnered through searching publications for which the candidate gene was specifically implicated in placental biology.

### Lookups of SNPs in GWAS of other phenotypes

We looked at associations between our independent PW-associated SNPs and other phenotypes within the phenoscanner^62^. We looked for associations with the SNP itself, SNPs in LD (*r*^*2*^ = 1, and *r*^*2*^ ⋝ 0.8). We additionally looked at associations between the independent PW SNPs in GWAS for nausea and vomiting of pregnancy, hyperemesis gravidarum^33^, preeclampsia^34^, gestational duration^35^, miscarriage (recurrent and spontaneous)^36^, and 10 cytokines assayed from neonatal blood spots^37^ (Supplementary Table 17).

### Mendelian randomization analysis

We then performed two-sample Mendelian randomization analyses with own and offspring PW as outcomes. The exposures used included height, fasting glucose, disposition index of insulin secretion, insulin sensitivity, systolic blood pressure (SBP) and diastolic blood pressure (DBP) using the same genetic instruments as the previous GWAS of BW (Supplementary Table 18)^15^. We estimated both the effect of each maternal exposure on PW, and also the effect of the fetal genetic predisposition to each exposure on PW. For the latter, we made the assumption that SNP-exposure effects in the fetus would be the same as in the adult samples in which they were identified. Effect sizes were converted to g using the value of 1 SD of PW of 132.5 g. We additionally used two-sample Mendelian randomization with its own PW as the exposure, and gestational age, and preeclampsia as outcomes. The SNP-PW associations were taken from our GWAS of PW, and the WLM-adjusted PW analysis. The SNP associations for all other traits were taken from external sources (Supplementary Table 17). To estimate the independent fetal associations for gestational age and preeclampsia, we first transformed the preeclampsia log odds ratios to the liability scale^63^, then applied the WLM in the same way as for PW. The preeclampsia independent fetal effects were then transformed to log odds ratios for analysis. A population prevalence for preeclampsia of 4.6% was used for the transformations^64^. We applied the inverse-variance weighted Mendelian randomization method, with MR-Egger^65^, Weighted Median, and Penalised Weighted Median^66^ acting as sensitivity analysis to test for robustness to Mendelian randomization assumptions.

### Polygenic scores

To investigate a possible association between PW and future risk of neuropsychiatric disease, we conducted analyses in the Initiative for Integrative Psychiatric Research (iPSYCH) cohort. The iPSYCH cohort is a Danish population based case–cohort sample including 141,215 individuals, out of which 50,615 constitute a population-representative sample and the remainder are individuals with one or more neuropsychiatric disease diagnoses^67^. After restriction to individuals of European ancestries born at gestational ages between 37 and 42 weeks, 100,094 individuals remained for analysis, including 20,328 cases of ADHD, 28,672 cases of affective disorder, 17,362 cases of autism spectrum disorder, 4,052 cases of schizophrenia, and population-based sample of 32,995 individuals without any one of the four disorders. Placental weight data from the Danish Medical Birth Register were available for 33,035 of these individuals (9,200 cases of ADHD, 2,946 cases of affective disorder, 9,678 cases of autism spectrum disorder, 239 cases of schizophrenia, and 13,113 controls).

We redid the fetal meta-analysis for PW excluding the iPSYCH data, and based on the resulting summary statistics and a genetic correlation matrix created from the iPSYCH genotypes, we used LD-pred2-auto^68^ to generate polygenic scores (PGS) for PW. Next, we regressed observed PW on ten fetal PW PGS quantiles, including sex, gestational age and ten principal components as covariates in the model. The linear regressions were done separately for the population controls and the four neuropsychiatric diseases. We also used logistic regression to assess the association between either 1) observed PW, or 2) fetal PGS of PW, and future risk of neuropsychiatric disease, including sex, gestational age and ten principal components as covariates in the model. In these analyses, the observed PW and the fetal PGS were first standardized to have mean 0 and variance 1.

## Data Availability

Individual cohorts contributing to the meta-analysis should be contacted directly as each cohort has different data access policies. GWAS summary statistics from this study are available via the EGG website (https://egg-consortium.org/).

## URLs

https://geneimprint.com/

https://platform.opentargets.org/

https://www.proteinatlas.org/

https://www.mousephenotype.org/

https://www.ldsc.broadinstitute.org/

https://www.ebi.ac.uk/gwas/home

## Ethics

Details of ethical approvals for the contributing studies are found in the Supplementary Information.

## Author contributions

The central analysis and manuscript drafting team included R.N.B., C.F., M.Vaudel, X.W., L.S., G.-H.M., D.M.E., B.J., M.-R.J., G.Z., M.-F.H, S.J., R.M.F., B.F., P.R.N.

Sample collection, genotyping and/or phenotyping were performed and/or directed by C.F., M.Vaudel, G.- H.M., Ø.H., P.S.-N., K.B., D.W., J.R.B., H.C., K.Hao, O.A.A., B.O.Å., M.A., L.Bhatta, L.Bouchard, B.M.B, S.B., J.B.-G., P.E., L.E., C.Erikstrup, M.E., S.F., R.G., F.G.E., J.G., D.M.H., E.K., C.S.M., E.A.N., M.N., C.N.A.P., O.B.P., F.R., B.M.S., C.S., I.S., L.W.T., H.U., M.Vaarasmaki, B.J.V., C.J.W., T.A.L., D.J.G.-B., M.B., T.H., E.R.P., R.R., S.R.O., C.E.P., V.W.V.J., J.F.F., A.T.H., M.M., D.A.L., K.Hveem, T.W., H.S.N., P.M., B.J., M.-R.J., M.-F.H., S.J., R.M.F., B.F., P.R.N.

Statistical analysis was performed by R.N.B., C.F., M.Vaudel, X.W., J.C., G.-H.M., L.S., C.Albiñana, J.R., J.F., S.E.S., K.T., C.A.W., S.Srinivasan, C.S.S., J.R.B., C.Allard, M.G., T.K., D.J.L., F.W., H.C., K.Hao, M.A., F.G.E., J.G., B.J.V., T.A.L., M.B., J.F.F., D.M.E., G.Z., B.F.

Data was interpreted by R.N.B., C.F., M.Vaudel, X.W., L.S., G.-H.M., J.C., Ø.H., P.S.-N., C.Albiñana, J.R., K.T., S.Srinivasan, C.Allard, M.G., F.W., P.-É.J., B.O.Å., L.Bhatta, B.M.B., C.Ebbing, P.E., S.F., J.G., E.K., C.N.A.P., F.R., S.Sebert, M.Vaarasmaki, C.J.W., E.R.P., R.R., V.W.V.J., J.F.F., D.A.L., K.Hveem, D.M.E., B.J., M.-R.J., G.Z., M.-F.H., S.J., R.M.F., B.F., P.R.N.

All authors contributed to the final version of the manuscript.

## Acknowledgements

We are grateful to Drs. Mirjana Efremova and Sarah A. Teichmann who shared the maternal-fetal interface single cell RNA data. Full acknowledgements and details of supporting grants are found in the Supplementary Information.

