## Supplementary Material for "Genome-wide association study of placental weight in 179,025 children and parents reveals distinct and shared genetic influences between placental and fetal growth"

[15]Department of Clinical Sciences, Lund University Diabetes Centre, Malmö , Sweden, [16]Institute for Molecular Medicine Finland (FIMM), University of Helsinki, Helsinki, Finland, [17]Novo Nordisk Foundation Center for Basic Metabolic Research, Faculty of Health and Medical Sciences, University of Copenhagen, Copenhagen, Denmark, [18]Department of Internal Medicine, Erasmus MC, University Medical Center Rotterdam, Rotterdam, The Netherlands, [19]Department of Human Genetics, McGill University, Montréal, Québec, Canada., [20]School of Medicine and Public Health, College of Medicine, Public Health and Wellbeing, The University of Newcastle, Newcastle, New South Wales, Australia, [21]Hunter Medical Research Institute, New Lambton Heights, New South Wales, Australia 2305, [22]Dept. of Obstetrics and Gynecology, Copenhagen University Hospital, Hvidovre, Denmark, [23]Methods and Analysis, Statistics Denmark. Copenhagen, Denmark, [24]Division of Population Health & Genomics, School of Medicine, University of Dundee, Dundee, [25]Centre for Cardiovascular Science, The University of Edinburgh, Edinburgh, United Kingdom, [26]Dpt. of Genetics, Physical Anthropology and Animal Physiology, University of the Basque Country (UPV/EHU) Leioa, Spain, [27]Biocruces-Bizkaia Health Research Institute, Barakaldo, Spain, [28]Spanish Biomedical Research Center in Diabetes and Associated Metabolic Disorders (CIBERDEM), Spain, [29]Centre de recherche du Centre Hospitalier de l'Universite de Sherbrooke, Sherbrooke, QC, [30]Département de Biologie, Faculté des Sciences, Université de Sherbrooke, Sherbrooke, Québec, Canada, [31]Institute of Biomedicine, School of Medicine, University of Eastern Finland, Kuopio Campus, Finland, [32]Icahn School of Medicine at Mount Sinai, [33]NORMENT Centre, Institute of Clinical Medicine, University of Oslo, Oslo, Norway, [34]Division of Mental Health and Addiction, Oslo University Hospital, Oslo, Norway, [35]K.G. Jebsen Center for Genetic Epidemiology, Department of Public Health and Nursing, NTNU, Norwegian University of Science and Technology, Trondheim, 7491, Norway, [36]HUNT Research Centre, Department of Public Health and Nursing, NTNU, Norwegian University of Science and Technology, Levanger, Norway, [37]Department of Endocrinology, Clinic of Medicine, St. Olavs Hospital, Trondheim University Hospital, Trondheim, Norway., [38]Department of Biochemistry and Functional Genomics, Faculty of Medicine and Health Sciences, Université de Sherbrooke, Sherbrooke, Québec, Canada., [39]Clinical Department of Laboratory Medicine, Centre intégré universitaire de santé et de services sociaux (CIUSSS) du Saguenay–Lac-St-Jean – Hôpital Universitaire de Chicoutimi, Saguenay, Québec, Canada., [40]Clinic of Medicine, St. Olavs Hospital, Trondheim University Hospital, Trondheim, Norway, [41]Department for Congenital Disorders, Statens Serum Institut, Copenhagen, Denmark, [42]iPSYCH, The Lundbeck Foundation Initiative for Integrative Psychiatric Research, Aarhus, Denmark, [43]Department of Clinical Science, University of Bergen, Bergen, Norway, [44]Department of Gynecology and Obstetrics, Haukeland University Hospital, Bergen, Norway, [45]Department of Epidemiology and Biostatistics, School of Public Health, Imperial College London, London, United Kingdom, [46]Department of Obstetrics and Gynecology, Hillerød Hospital, Denmark, [47]Dept. Clinical Immunology, Aarhus University Hospital, Aarhus, Denmark, [48]Dept. Clinical Medicine, Aarhus University, Aarhus, Denmark, [49]Faculty of nursing and chiropody, Universitat de València, C/Menendez Pelayo, Valencia, Spain, [50]Epidemiology and Environmental Health Joint Research Unit, Foundation for the Promotion of Health and Biomedical Research in the Valencian Region, FISABIO-Public Health, FISABIO-Universitat Jaume I-Universitat de València, Valencia, Spain, [51]Spanish Consortium for Research on Epidemiology and Public Health (CIBERESP), Av. Monforte de Lemos, Madrid, Spain, [52]Institute of Reproductive and Developmental Biology, Imperial College London, United Kingdom, [53]The Generation R Study Group, Erasmus MC, University

Medical Center Rotterdam, Rotterdam, The Netherlands, [54]Department of Pediatrics, Erasmus MC, University Medical Center Rotterdam, Rotterdam, The Netherlands, [55]Department of Biomedicine - Human Genetics and the iSEQ Center, Aarhus University, Aarhus, Denmark, [56]Center for Genomics and Personalized Medicine, Aarhus, Denmark, [57]Bioinformatics Research Centre, Aarhus University, Aarhus, Denmark, [58]PEDEGO Research Unit, Medical Research Center, University of Oulu and Oulu University Hospital, Oulu, Finland, [59]Population Health Unit, Finnish Institute for Health and Welfare, Helsinki and Oulu, Finland, [60]National Institute of Public Health, University of Southern Denmark, Copenhagen, Denmark, [61]Institute of Clinical research, University of Southern Denmark, Odense, Denmark, [62]Department of Health Science and Technology, Aalborg University, Denmark, [63]Dept. of Clinical Immunology, Zealand University Hospital, Køge, Denmark, [64]Dept. of Clinical Medicine, Faculty of Health and Medical Sciences, University of Copenhagen, Copenhagen, Denmark, [65]A list of members and affiliations appears in the Supplementary Note, [66]Department for Genomics of Common Diseases, School of Medicine, Imperial College London, London United Kingdom, [67]Department of Clinical and Molecular Medicine, Norwegian University of Science and Technology, Trondheim, Norway, [68]Pediatric Research Centre, Helsinki University Hospital and University of Helsinki, Helsinki, Finlan, [69]Norwegian Institute of Public Health, Oslo, Norway, [70]Department of Global Public Health and Primary Care, University of Bergen, Bergen, Norway, [71]Department of Internal Medicine, Division of Cardiology, University of Michigan, Ann Arbor, MI, USA, [72]Dept. of Clinical Immunology, Copenhagen University Hospital, Rigshospitalet, Copenhagen, Denmark, [73]Statens Serum Institut, Copenhagen, Denmark, [74]PEDEGO Research Unit, Medical Research Center, University of Oulu, Oulu, Finland, [75]Department of Obstetrics and Gynaecology, Oulu University Hospital, Oulu, Finland, [76]Department of Biostatistics and Center for Statistical Genetics, University of Michigan, Ann Arbor, MI, USA, [77]Department of Human Genetics, University of Michigan, Ann Arbor, MI, USA, [78]Department of Clinical Physiology and Nuclear Medicine, Kuopio University Hospital, Kuopio, Finland, [79]Foundation for Research in Health Exercise and Nutrition, Kuopio Research Institute of Exercise Medicine, Kuopio, Finland, [80]Psychotherapeutic Outpatient Clinic, Mental Health Services, Capital Region, Copenhagen Denmark, [81]ISGlobal, Institute for Global Health, Barcelona, Spain, [82]Universitat Pompeu Fabra (UPF), Barcelona, Spain, [83]Spanish Consortium for Research on Epidemiology and Public Health (CIBERESP), Madrid, Spain, [84]Dept. of Clinical Medicine, University of Copenhagen, Copenhagen, Denmark, [85]Department of Clinical Medicine, University of Copenhagen, Copenhagen, Denmark, [86]K.G. Jebsen Center for Genetic Epidemiology, Norwegian University of Science and Technology, Trondheim, Norway, [87]Norwegian Institute of Public Health, Oslo, Norway, [88]Department of Genetics, Stanford University School of Medicine, Stanford, California, USA, [89]Medical Research Council Integrative Epidemiology Unit, University of Bristol, Bristol, United Kingdom, [90]Population Health Science, Bristol Medical School, University of Bristol, Bristol, UK, [91]Institute of Biological Psychiatry, Mental Health Services, Copenhagen University Hospital, Copenhagen, Denmark, [92]Lundbeck Center for Geogenetics, GLOBE Institute, University of Copenhagen, Copenhagen, Denmark, [93]Centre for Fertility and Health, Norwegian Institute of Public Health, Oslo, Norway., [94]MRC Centre for Environment and Health, School of Public Health, Imperial College London, London, [95]Unit of Primary Health Care, Oulu University Hospital, OYS, Oulu, Finland, [96]Division of Human Genetics, Cincinnati Children's Hospital Medical Center, Department of Pediatrics, University of Cincinnati College of Medicine, Cincinnati, Ohio, United States of America, [97]Center for Prevention of Preterm Birth, Perinatal Institute, Cincinnati Children's

Hospital Medical Center and March of Dimes Prematurity Research Center Ohio Collaborative, Cincinnati Children's Hospital Medical Center, Department of Pediatrics, University of Cincinnati College of Medicine, Cincinnati, Ohio, United States of America, [98]Department of Population Medicine, Harvard Medical School, Harvard Pilgrim Health Care Institute, Boston, MA, [99]Diabetes Unit, Massachusetts General Hospital, MA, Boston, [100]Department of Medical Genetics, Haukeland University Hospital, Bergen, Norway, [101]Children and Youth Clinic, Haukeland University Hospital, Bergen, Norway

*These authors contributed equally to this work
**These authors jointly directed this work

### Content

| Item | Text | Page |
|---|---|---|
| Supplementary Figure 1 | Effect sizes and minor allele frequencies for placental weight-associated lead SNPs | 8 |
| Supplementary Figure 2a | Graphical display of locus-specific association results from the fetal GWAS: rs150138294, rs723177, rs1655296, rs10925945, rs4953353, rs11708067, rs9817452, rs74457440, rs7722058, rs72801474, rs3822394, rs67265526 | 9 |
| Supplementary Figure 2b | Graphical display of locus-specific association results from the fetal GWAS: rs9800506, rs12529634, rs11756568, rs1021508, rs7783810, rs10486660, rs138715366, rs6557677, rs12543725, rs1434836, rs1801253, rs2237892 | 10 |
| Supplementary Figure 2c | Graphical display of locus-specific association results from the fetal GWAS: rs112635299, rs7177338, rs55958435, rs57790054, rs11866404, rs876987, rs6040436 | 11 |
| Supplementary Figure 2d | Graphical display of locus-specific association results from the maternal GWAS: rs72804545, rs2168101, rs180435, rs303998 | 12 |
| Supplementary Figure 2e | Graphical display of locus-specific association results from the paternal GWAS: rs75512885, rs2207099 | 13 |
| Supplementary Figure 3 | Heritability estimates for PW and BW from genomicSEM analysis | 14 |
| Supplementary Figure 4a | Fetal classified loci effects from meta analysis and WLM for each genome | 15 |
| Supplementary Figure 4b | Classifications of remaining loci and effects from meta analysis and WLM for each genome | 16 |
| Supplementary Figure 5a | Conditioned effect size comparison between placental and birth weight, loci 1-16: *DCST2* (rs150138294), RPL31P11 (rs723177), *TSNAX−DISC1,LINC00582* (rs1655296), *TSNAX−DISC1,LINC00582** (rs140691414), *CHRM3* (rs10925945), *EPAS1* (rs4953353), *ADCY5* (rs11708067), *LOC339894/CCNL1* (rs9817452), *PDLIM5* (rs74457440), *ACTBL2* (rs7722058), *HSPA4* (rs72801474), *ARHGAP26* (rs3822394), *EBF1* (rs72804545), *EBF1* (rs75512885), *EBF1* (rs67265526), *NUDT3* (rs541641049) | 17 |
| Supplementary Figure 5b | Conditioned effect size comparison between placental and birth weight, loci 17-32: *FKBP5/MAPK13/TEAD3* (rs9800506), *HACE1* (rs12529634), *ESR1* (rs11756568), *PDE10A* (rs1021508), *PDE10A* (rs6456014), *ISPD* | 20 |

| | | |
|---|---|---|
| | (rs7783810), *TBX20* (rs10486660), *YKT6* (rs138715366), *ENTPD4* (rs6557677), *SLC45A4* (rs12543725), *KLF4* (rs1434836), *ADRB1* (rs1801253), *KCNQ1* (rs2237892), *LMO1* (rs2168101), *SLC38A4* (rs180435), *SERPINA1* (rs112635299) | |
| Supplementary Figure 5c | Conditioned effect size comparison between placental and birth weight, loci 33-41: *FES/FURIN* (rs7177338), *NR2F2* (rs55958435), *GPR139/GPRC5B* (rs57790054), *SLC6A2* (rs11866404), *SLC7A5* (rs876987), *NLRP13* (rs303998), *LOC339593* (rs6040436), *LOC339593* (rs2207099), *LOC339593** (rs6078190) | 23 |
| Supplementary Figure 6 | Scatter plots comparing effect sizes from placental and birth weight GWAS for placental weight SNPs | 24 |
| Supplementary Figure 7 | Tissue enrichment by mRNA data | 25 |
| Supplementary Figure 8 | Cell-type enrichment by scRNA-seq data | 26 |
| Supplementary Figure 9a | Graphical display of methylated locus-specific association results, loci: rs723177, rs1655296, rs4953353, rs11708067, rs9817452, rs74457440 | 27 |
| Supplementary Figure 9b | Graphical display of methylated locus-specific association results, loci: rs3822394, rs541641049, rs9800506, rs12529634, rs11756568, rs1021508 | 30 |
| Supplementary Figure 9c | Graphical display of methylated locus-specific association results, loci: rs10486660, rs6557677, rs12543725, rs1801253, rs2168101, rs180534 | 33 |
| Supplementary Figure 9d | Graphical display of methylated locus-specific association results, loci: rs7177338, rs55958435, rs876987 | 36 |
| Supplementary Figure 10a-l | Quantile–quantile (QQ) plots of observed versus expected $-\log_{10}$ *P* values, various pregnancy and perinatal traits | 37 |
| Supplementary Figure 10m-u | Quantile–quantile (QQ) plots of observed versus expected $-\log_{10}$ *P* values, additional pregnancy and perinatal traits | 38 |
| Supplementary Figure 11 | Scatter plots | 39 |
| Supplementary Figure 12 | Mendelian randomization analysis | 40 |
| Supplementary Figure 13 | Polygenic score analysis | 42 |
| Supplementary Figure 14 | Analyses in the iPSYCH cohort of placental weight and risk of neuropsychiatric diseases | 43 |
| Supplementary Figure 15 | Genomic SEM model | 44 |

| | | |
|---|---|---|
| Supplementary Figure 16 | Enrichment test using tissue or cell-type specific expression data | 45 |
| Supplementary Table 1 | Study descriptions (fetal GWAS, cohort data) | 46 |
| Supplementary Table 2 | Study descriptions (fetal GWAS, genetic data) | 46 |
| Supplementary Table 3 | Study descriptions (maternal GWAS, cohort data) | 46 |
| Supplementary Table 4 | Study descriptions (maternal GWAS, genetic data) | 46 |
| Supplementary Table 5 | Study descriptions (paternal GWAS, cohort data) | 46 |
| Supplementary Table 6 | Study descriptions (paternal GWAS, genetic data) | 46 |
| Supplementary Table 7 | Summary statistics from the GWAS of fetal, maternal, and paternal genotype with birth weight | 46 |
| Supplementary Table 8 | Association results for the lead SNPs from Supplementary Table 7 in the sex-only adjusted meta-analysis | 47 |
| Supplementary Table 9 | Estimates of variance, and correlation of the different variables in the genomic SEM model | 47 |
| Supplementary Table 10 | Summary statistics from the WLM and parent-offspring conditional analysis in MoBa for the 41 lead SNPs | 47 |
| Supplementary Table 11 | Fetal and parental contributions to association signals based on MobBa data compared to results on birth weight by Juliusdottir *et al*[1]. | 47 |
| Supplementary Table 12 | Ranks of gene expression of the 31 placental weight associated genes with fetal effect in placenta and the three cell-types reached multiple-testing corrected significance level in enrichment analysis. | 47 |
| Supplementary Table 13 | Annotation of SNPs at placental weight loci with respect to GWAS Catalog associations | 48 |
| Supplementary Table 14 | Annotation of SNPs at placental weight loci with respect to expression quantitative trait loci (eQTL) | 48 |
| Supplementary Table 15 | Annotation of SNPs at placental weight loci with respect to protein quantitative trait loci (pQTL) | 48 |
| Supplementary Table 16 | Lookups of placental weight lead SNPs in external GWAS of preeclampsia, recurrent and spontaneous miscarriage, fetal cytokines, birth weight, gestational age, hyperemesis/nausea and vomiting of pregnancy, and endometriosis | 48 |
| Supplementary Table 17 | Mendelian randomization analyses for effects of placental and birth weight on pregnancy outcomes | 48 |
| Supplementary Table 18 | Mendelian randomization analyses for effects of maternal exposures on placental weight | 48 |

| | | |
|---|---|---|
| Acknowledgements | | 49 |
| References | | 58 |
| EGG Membership | | 59 |

### Supplementary Figures

**Supplementary Fig. 1: Effect sizes and minor allele frequencies for placental weight-associated lead SNPs.**

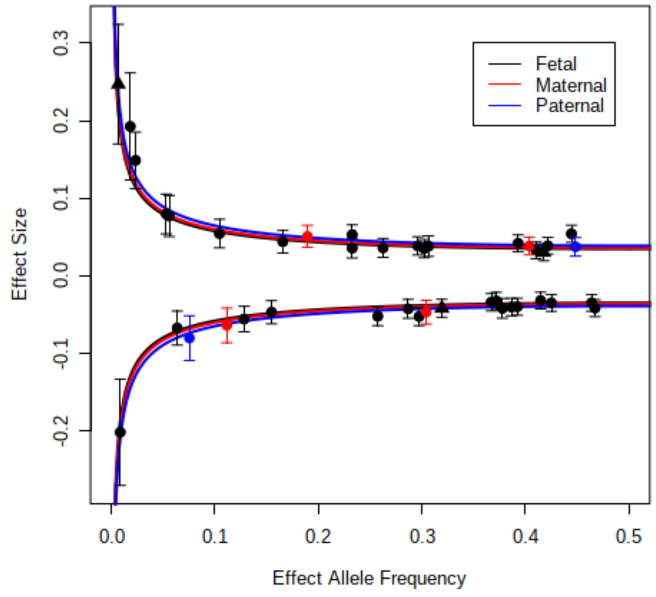

Variants identified in the fetal, maternal and paternal analyses are shown in black, red, and blue, respectively. The lines indicate effect sizes needed to have 80% power to detect variants at genome-wide significance with the sample sizes of the fetal, maternal, and paternal analyses. Circles indicate main signals and triangles indicate secondary signals.

**Supplementary Fig. 2a: Graphical display of locus-specific association results from the fetal GWAS: rs150138294, rs723177, rs1655296, rs10925945, rs4953353, rs11708067, rs9817452, rs74457440, rs7722058, rs72801474, rs3822394, rs67265526.**

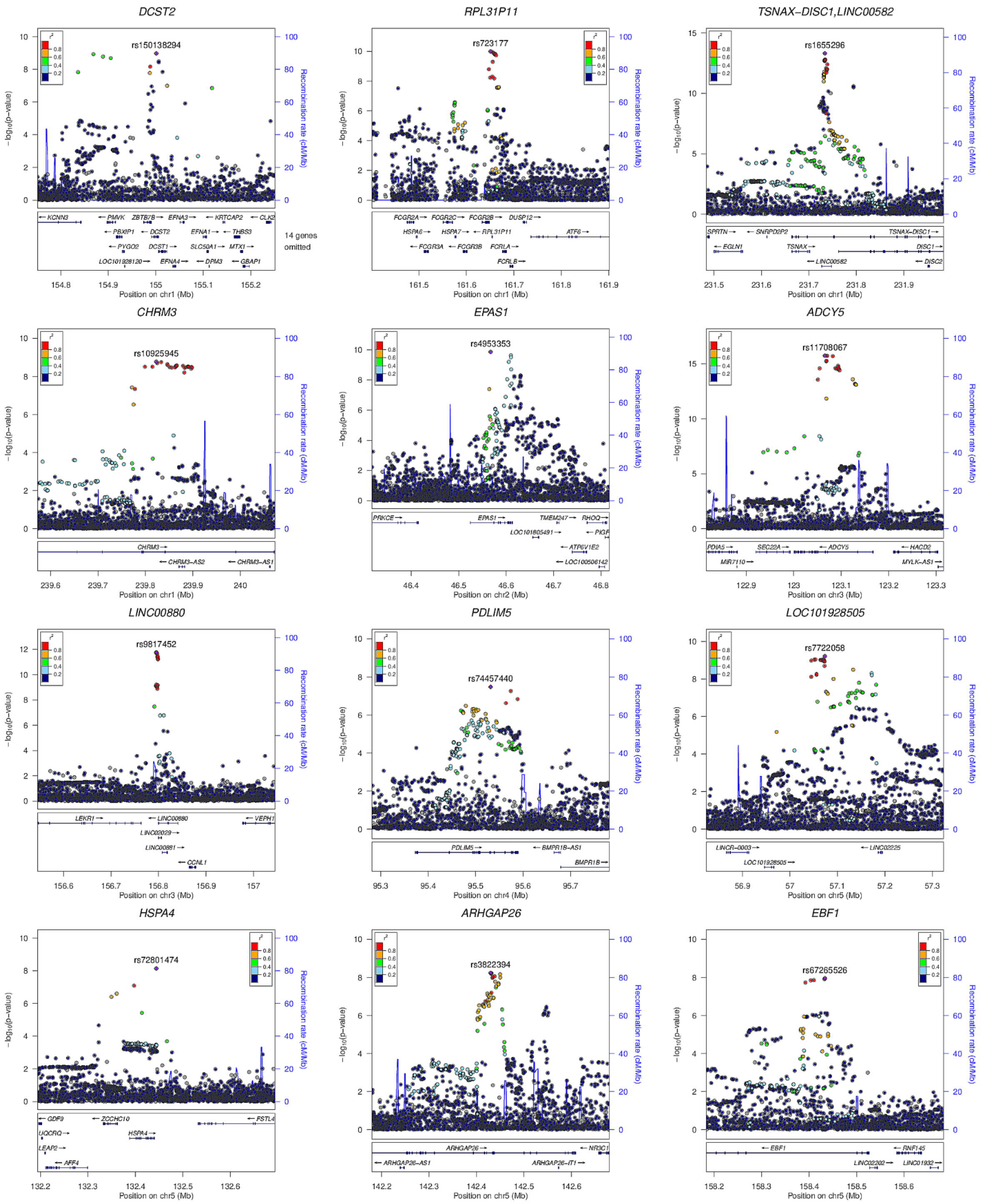

Shown are LocusZoom plots providing overview of the extent of LD and the position relative to nearby genes and local recombination hotspots for the fetal loci rs150138294, rs723177, rs1655296, rs10925945, rs4953353, rs11708067, rs9817452, rs74457440, rs7722058, rs72801474, rs3822394, rs67265526. Abbreviation: Chr, chromosome.

**Supplementary Fig. 2b: Graphical display of locus-specific association results from the fetal GWASi: rs9800506, rs12529634, rs11756568, rs1021508, rs7783810, rs10486660, rs138715366, rs6557677, rs12543725,  rs1434836, rs1801253, rs2237892.**

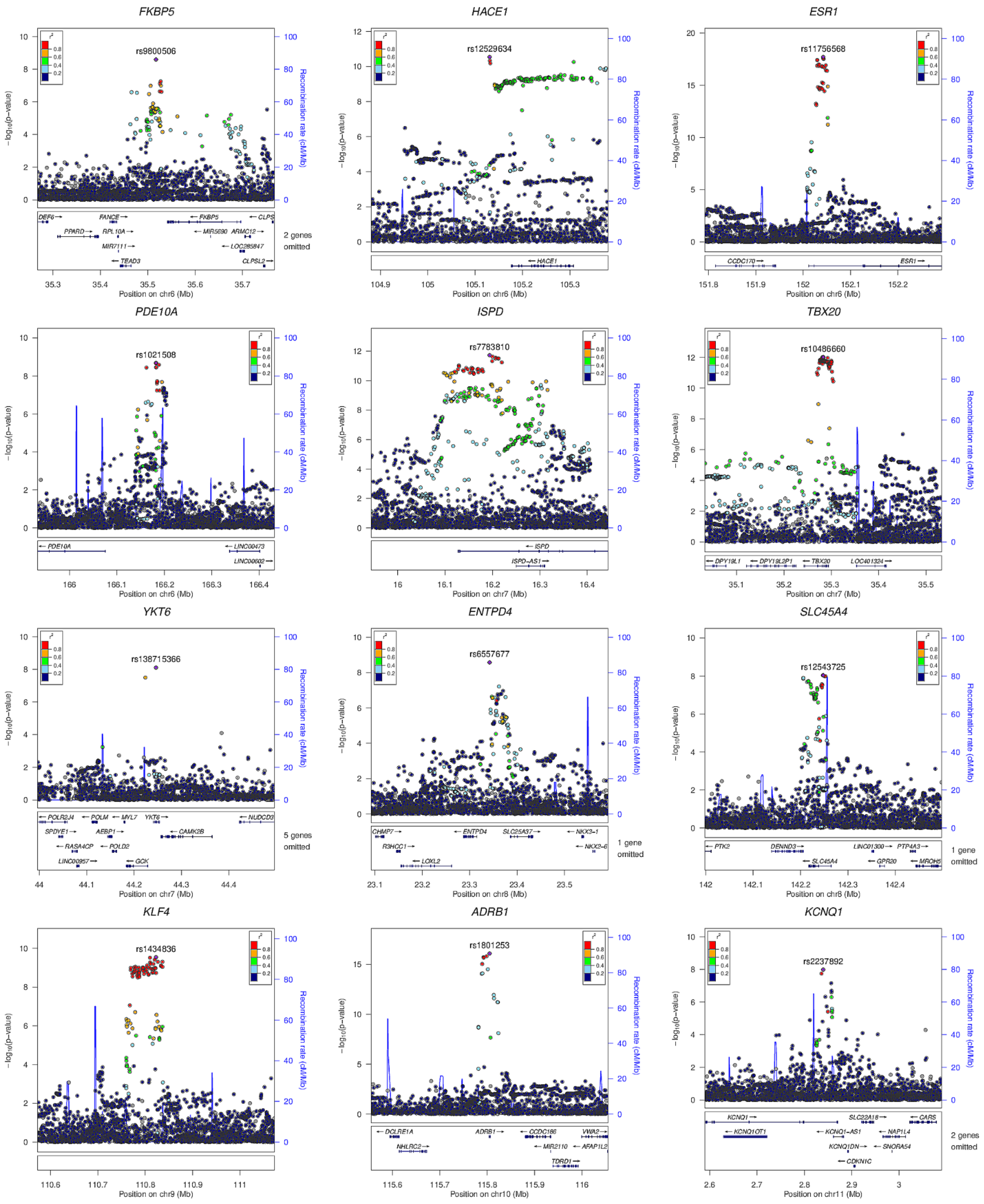

Shown are LocusZoom plots providing overview of the extent of LD and the position relative to nearby genes and local recombination hotspots for the fetal loci rs9800506, rs12529634, rs11756568, rs1021508, rs7783810, rs10486660, rs138715366, rs6557677, rs12543725,  rs1434836, rs1801253, rs2237892. Abbreviation: Chr, chromosome.

**Supplementary Fig. 2c: Graphical display of locus-specific association results from the fetal GWAS: rs112635299, rs7177338, rs55958435, rs57790054, rs11866404, rs876987, rs6040436.**

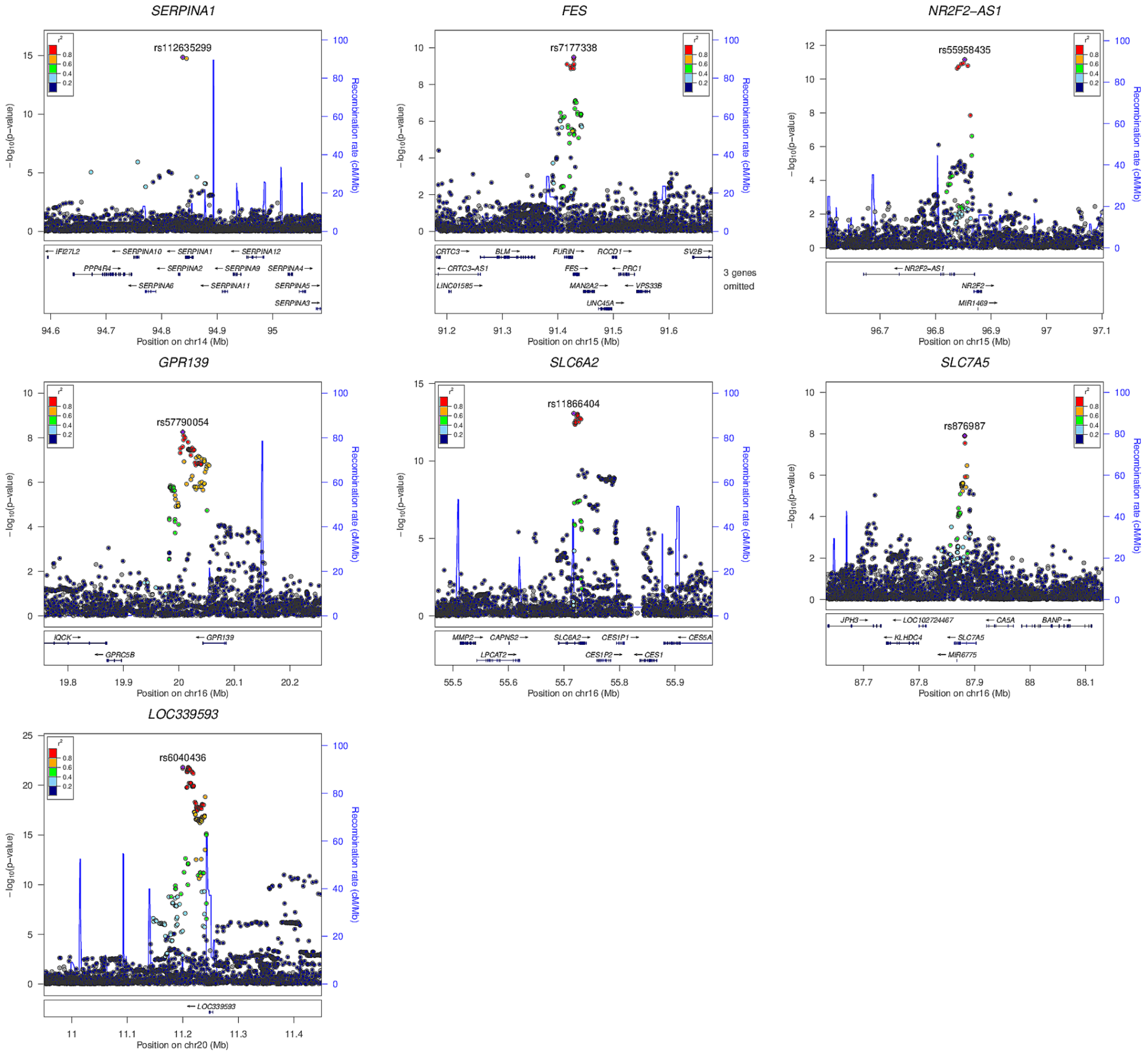

Shown are LocusZoom plots providing overview of the extent of LD and the position relative to nearby genes and local recombination hotspots for the fetal loci rs112635299, rs7177338, rs55958435, rs57790054, rs11866404, rs876987, rs6040436. Abbreviation: Chr, chromosome.

**Supplementary Fig. 2d: Graphical display of locus-specific association results from the maternal GWAS: rs72804545, rs2168101, rs180435, rs303998.**

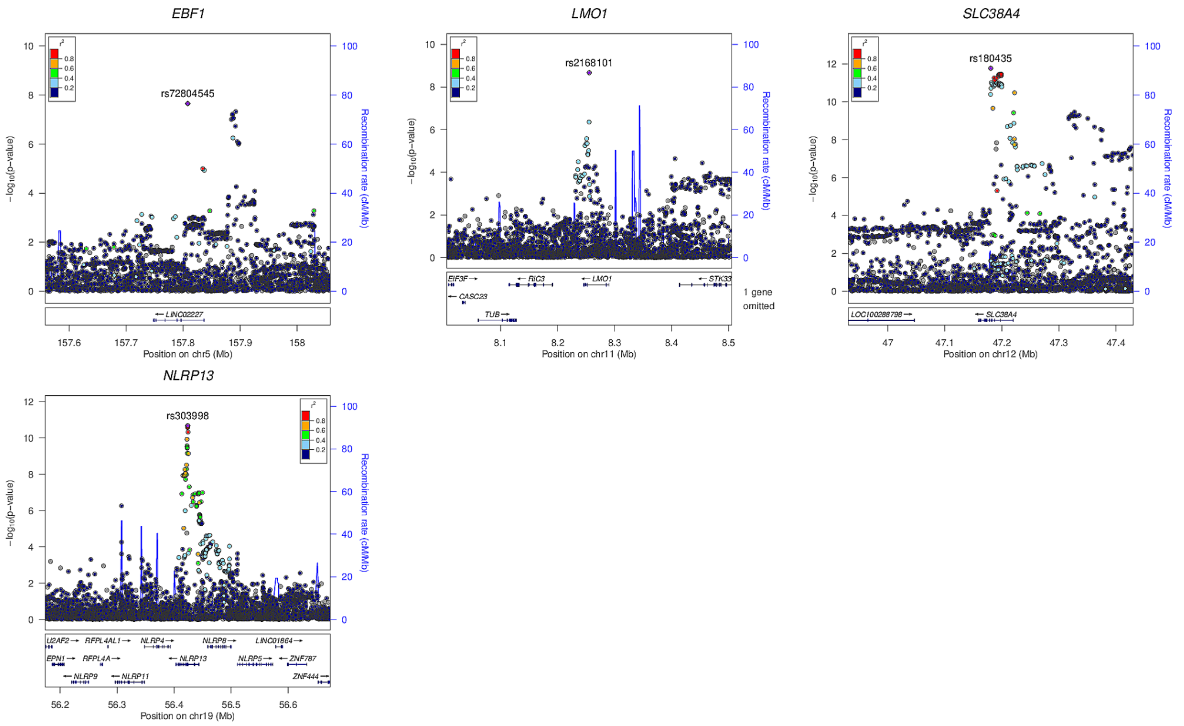

Shown are LocusZoom plots providing overview of the extent of LD and the position relative to nearby genes and local recombination hotspots for the maternal loci rs72804545, rs2168101, rs180435, rs303998. Abbreviation: Chr, chromosome.

**Supplementary Fig. 2e: Graphical display of locus-specific association results from the paternal GWAS: rs75512885, rs2207099.**

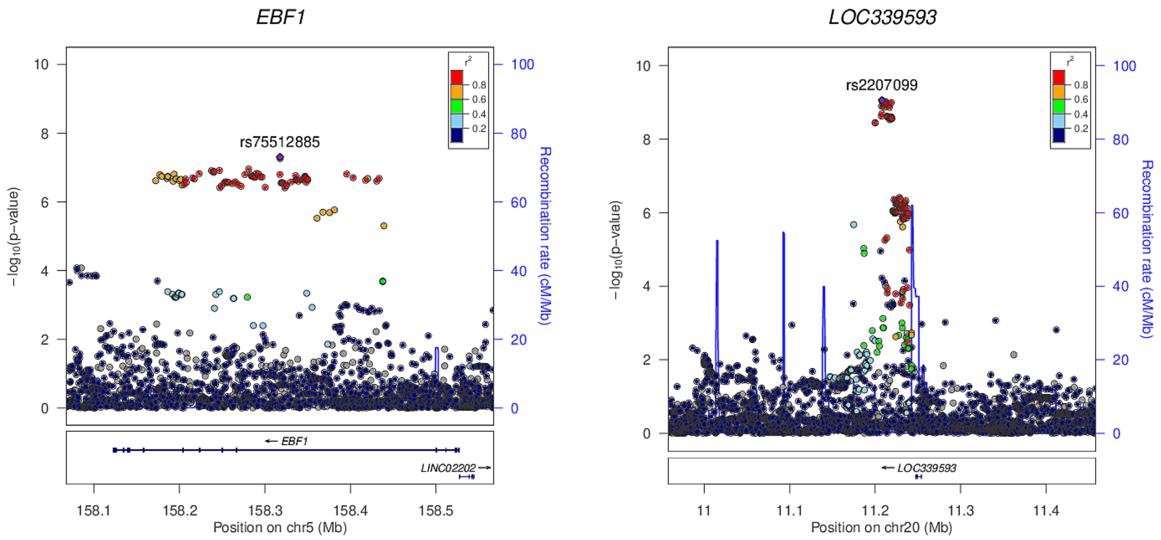

Shown are LocusZoom plots providing overview of the extent of LD and the position relative to nearby genes and local recombination hotspots for the two paternal loci rs75512885, rs2207099. Abbreviation: Chr, chromosome.

**Supplementary Fig. 3: Heritability estimates for PW and BW from genomic SEM analysis.**

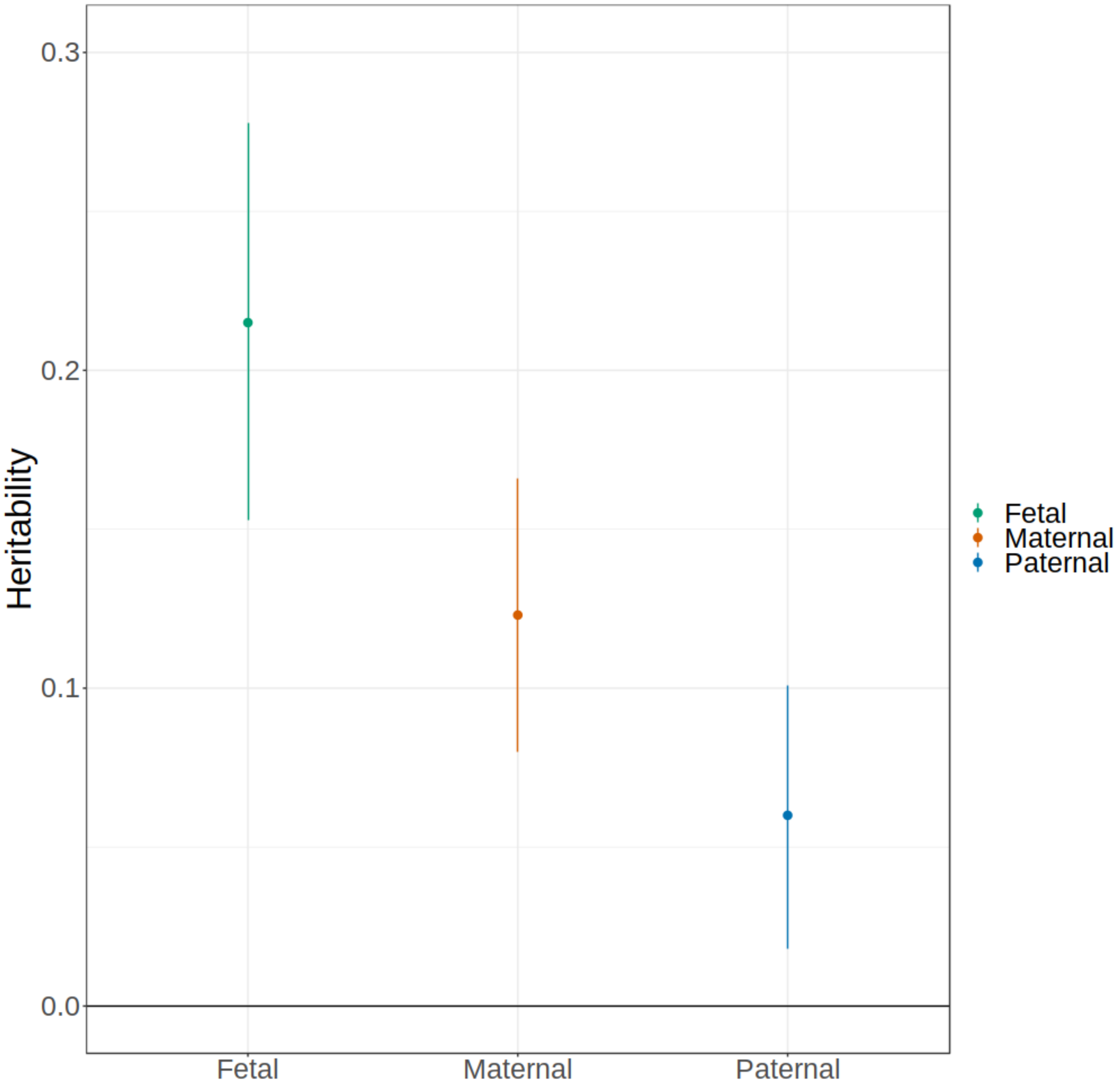

Scatter plot showing SNP heritability estimates ($h^2$) for fetal, maternal and paternal genomes estimated using genomic SEM.

**Supplementary Fig. 4a: Fetal classified loci: effects from meta-analysis and weighted linear model for each genome.**

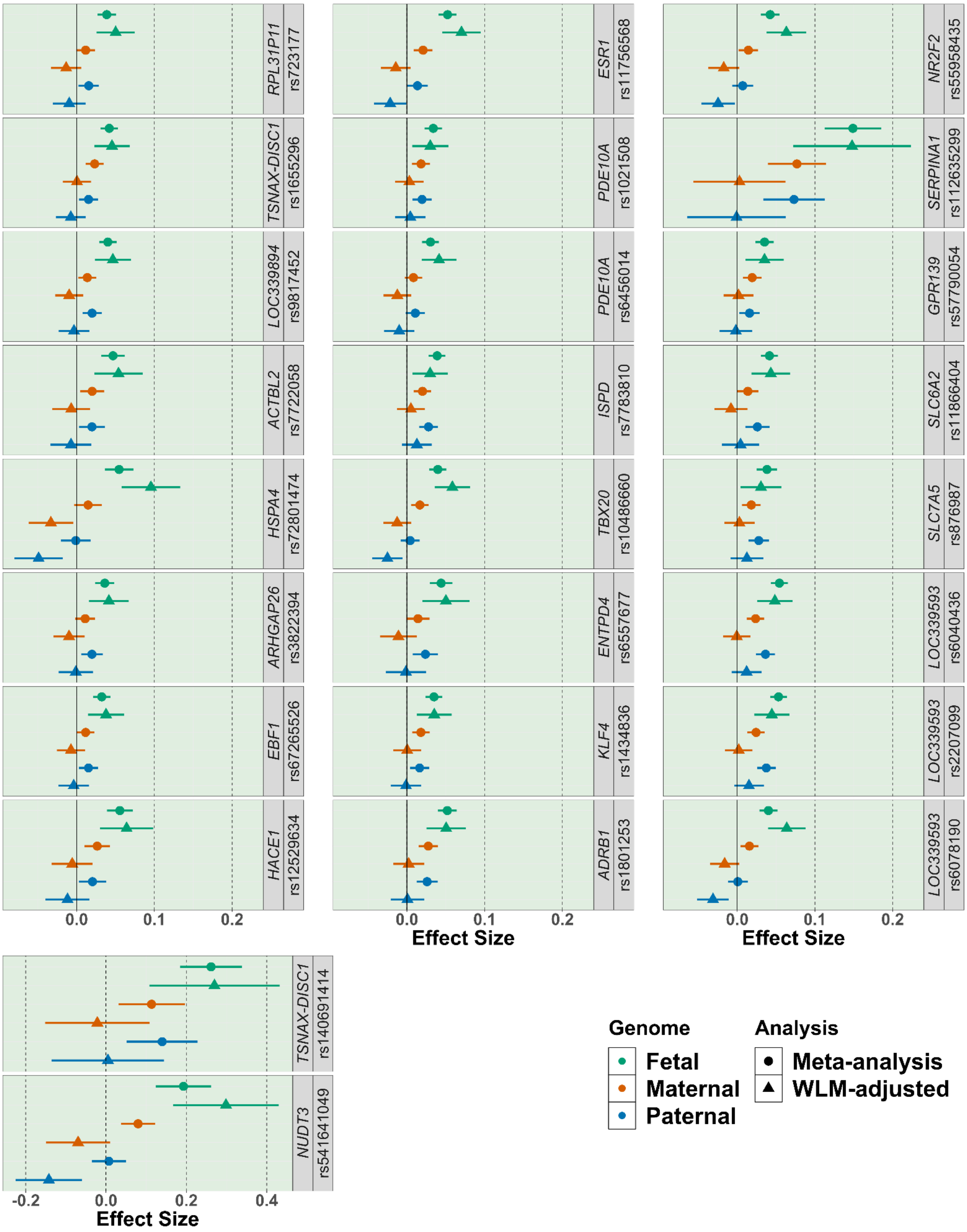

Shown are the variants which were classified as having a fetal effect (represented by the green background). Estimates are provided for fetal, maternal, and paternal effects for the meta-analysis results and after weighted linear model adjustment. Error bars represent 95% confidence intervals. Abbreviation: WLM, weighted linear model. ***Note different scale on x-axis for rs140691414 - *TSNAX-DISC1* & rs541541049 *NUDT3***

**Supplementary Fig. 4b: Classifications of remaining loci and effects from meta analysis and weighted linear model for each genome.**

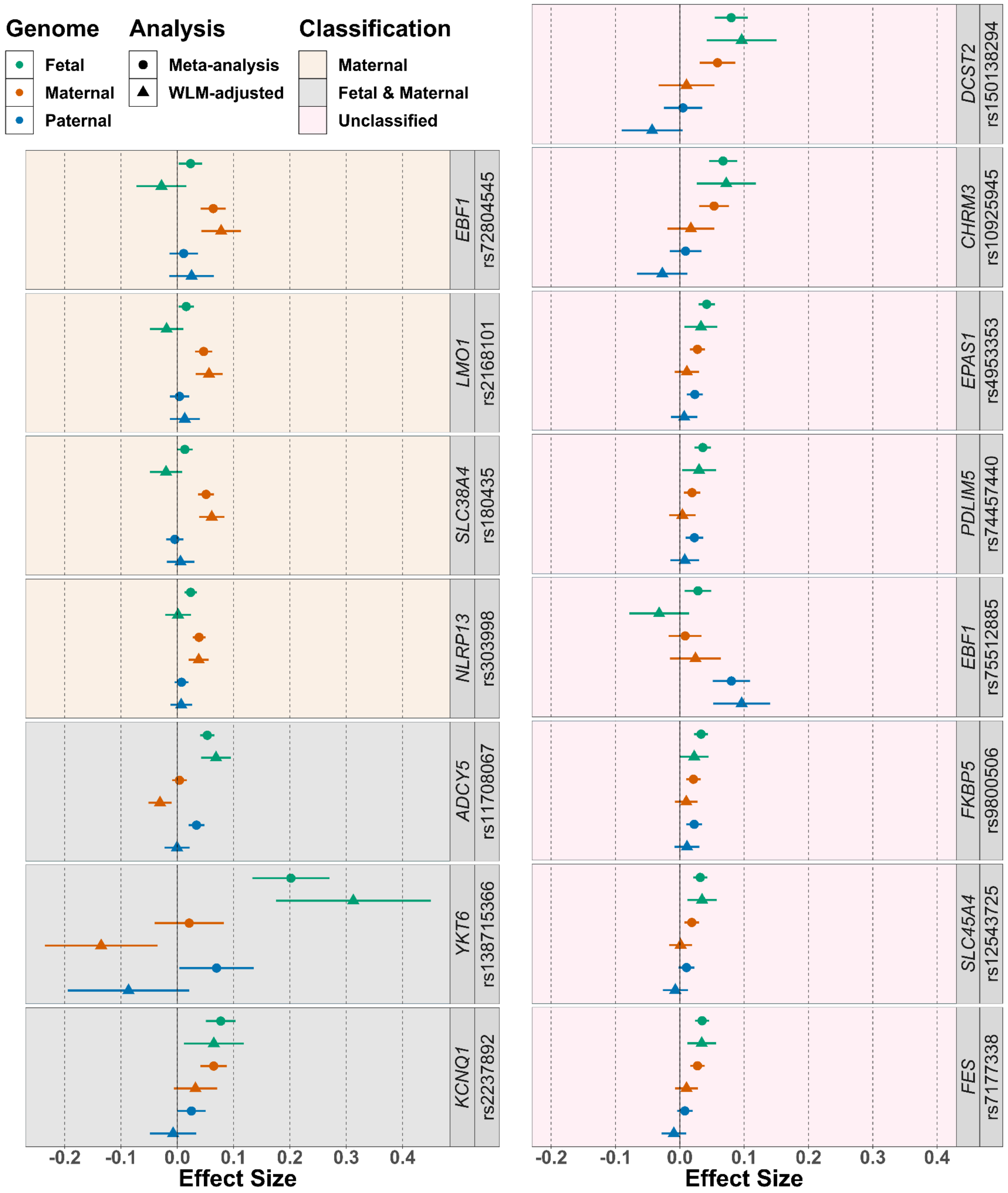

Shown are the remaining variants, with backgrounds indicating the classification of effect. Estimates are provided for fetal, maternal, and paternal effects for the meta-analysis results and after weighted linear model adjustment. Error bars represent 95% confidence intervals. Abbreviation: WLM, weighted linear model.

**Supplementary Fig. 5a: Conditioned effect size comparison between placental and birth weight, in MoBa trios, loci 1-16: *DCST2* (rs150138294), *RPL31P11* (rs723177), *TSNAX−DISC1,LINC00582* (rs1655296), *TSNAX−DISC1,LINC00582** (rs140691414), *CHRM3* (rs10925945), *EPAS1* (rs4953353), *ADCY5* (rs11708067), *LOC339894/CCNL1* (rs9817452), *PDLIM5* (rs74457440), *ACTBL2* (rs7722058), *HSPA4* (rs72801474), *ARHGAP26* (rs3822394), *EBF1* (rs72804545), *EBF1* (rs75512885), *EBF1* (rs67265526), *NUDT3* (rs541641049).**

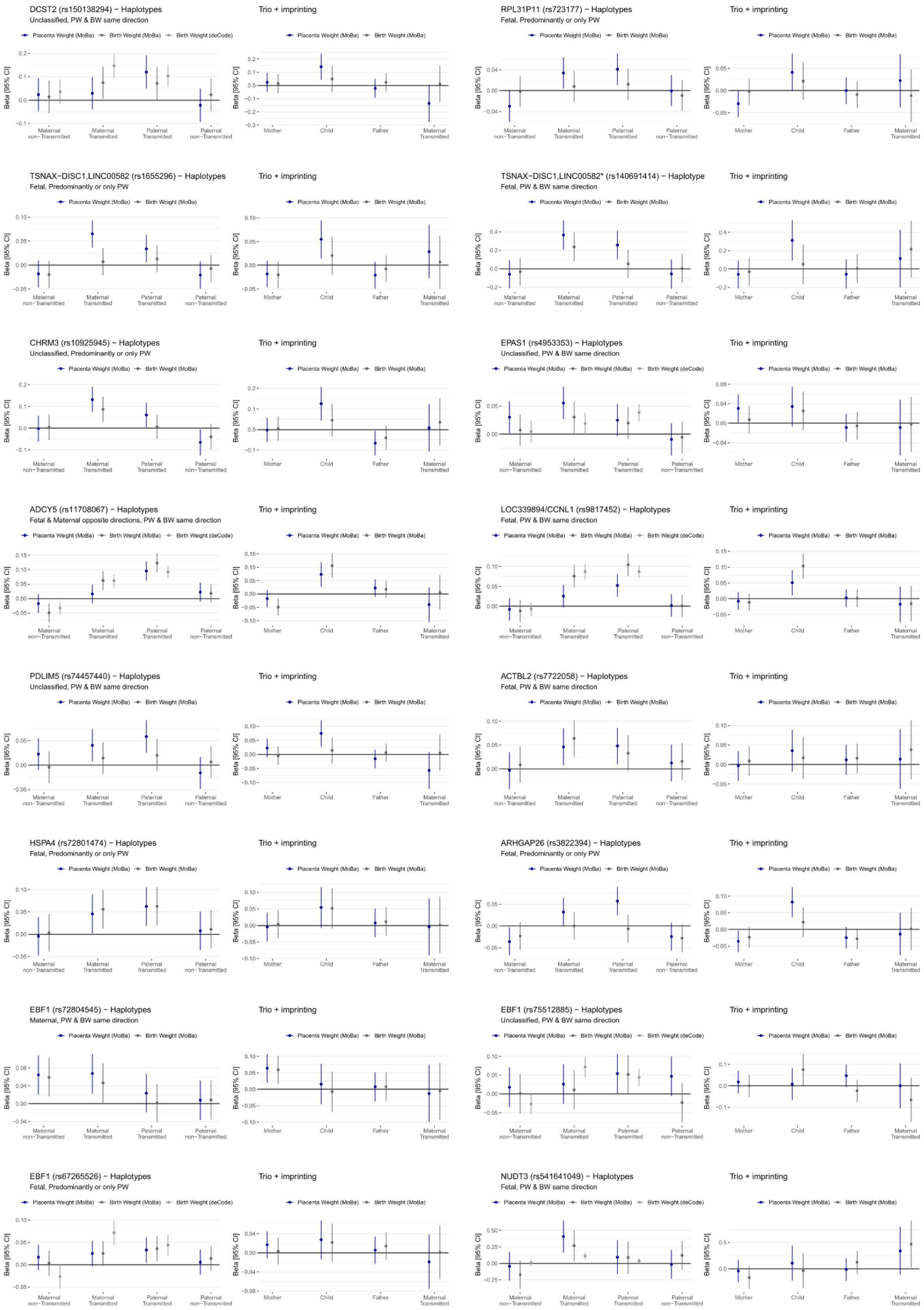

Effect size estimates with 95% confidence intervals of the genetic association with placental and birth weight conditioning child, mother, and father genotypes against each other. For all loci, estimates were computed on unrelated {child, mother, father} trios of the MoBa cohort using two models: (1) left, maternal and paternal alleles separated based on their transmission status; and (2) right, individual genotypes and the maternal transmitted allele. Where a proxy was found, effect sizes obtained for modes of transmission of birth weight by Juliusdottir *et al.*[1] are displayed.

**Supplementary Fig. 5b: Conditioned effect size comparison between placental and birth weight, loci 17-32:*FKBP5/MAPK13/TEAD3* (rs9800506), *HACE1* (rs12529634), *ESR1* (rs11756568), *PDE10A* (rs1021508), *PDE10A* (rs6456014), *ISPD* (rs7783810), *TBX20* (rs10486660), *YKT6* (rs138715366), *ENTPD4* (rs6557677), *SLC45A4* (rs12543725), *KLF4* (rs1434836), *ADRB1* (rs1801253), *KCNQ1* (rs2237892), *LMO1* (rs2168101), *SLC38A4* (rs180435), *SERPINA1* (rs112635299).**

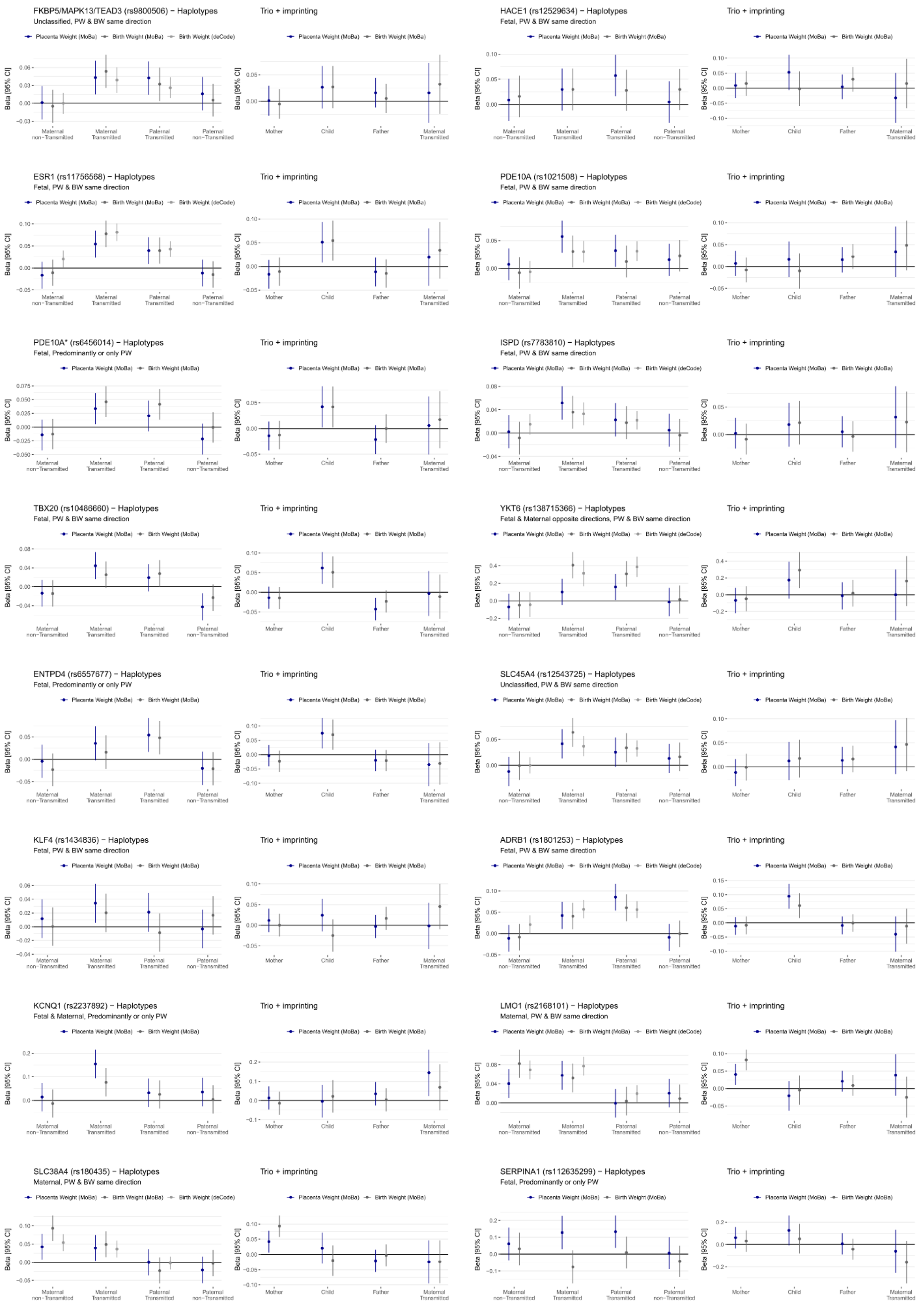

Effect size estimates with 95% confidence intervals of the genetic association with placental and birth weight conditioning child, mother, and father genotypes against each other. For all loci, estimates were computed on unrelated {child, mother, father} trios of the MoBa cohort using two models: (1) left, maternal and paternal alleles separated based on their transmission status; and (2) right, individual genotypes and the maternal transmitted allele. Where a proxy was found, effect sizes obtained for modes of transmission of birth weight by Juliusdottir *et al.*[1] are displayed.

**Supplementary Fig. 5c: Conditioned effect size comparison between placental and birth weight, loci 23-41: *FES/FURIN* (rs7177338), *NR2F2* (rs55958435), *GPR139/GPRC5B* (rs57790054), *SLC6A2* (rs11866404), *SLC7A5* (rs876987), *NLRP13* (rs303998), *LOC339593* (rs6040436), *LOC339593* (rs2207099), *LOC339593** (rs6078190).**

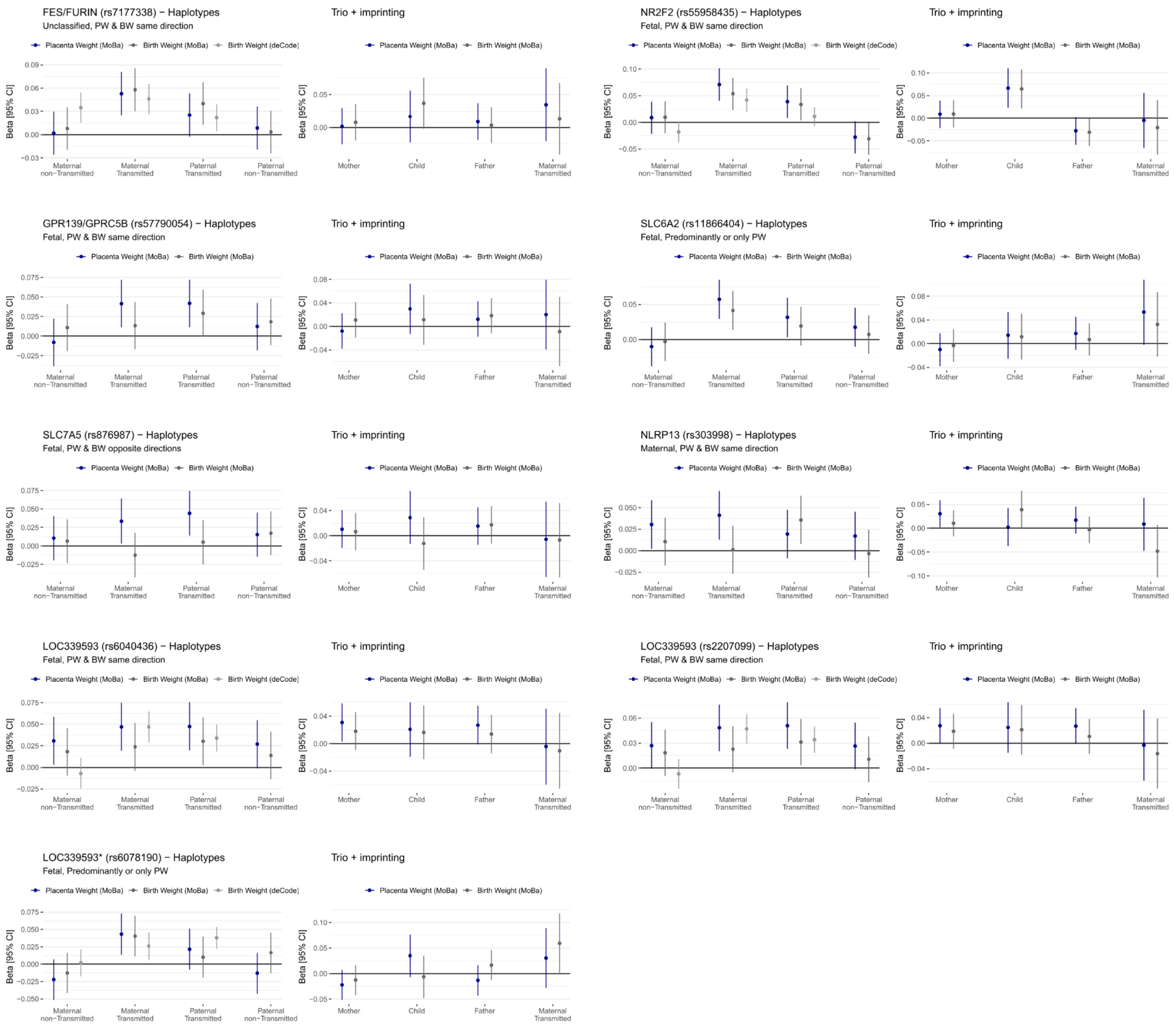

Effect size estimates with 95% confidence intervals of the genetic association with placental and birth weight conditioning child, mother, and father genotypes against each other. For all loci, estimates were computed on unrelated {child, mother, father} trios of the MoBa cohort using two models: (1) left, maternal and paternal alleles separated based on their transmission status; and (2) right, individual genotypes and the maternal transmitted allele. Where a proxy was found, effect sizes obtained for modes of transmission of birth weight by Juliusdottir *et al.*[1] are displayed.

**Supplementary Fig. 6: Scatter plots comparing effect sizes from PW and BW GWAS for placental weight SNPs.**

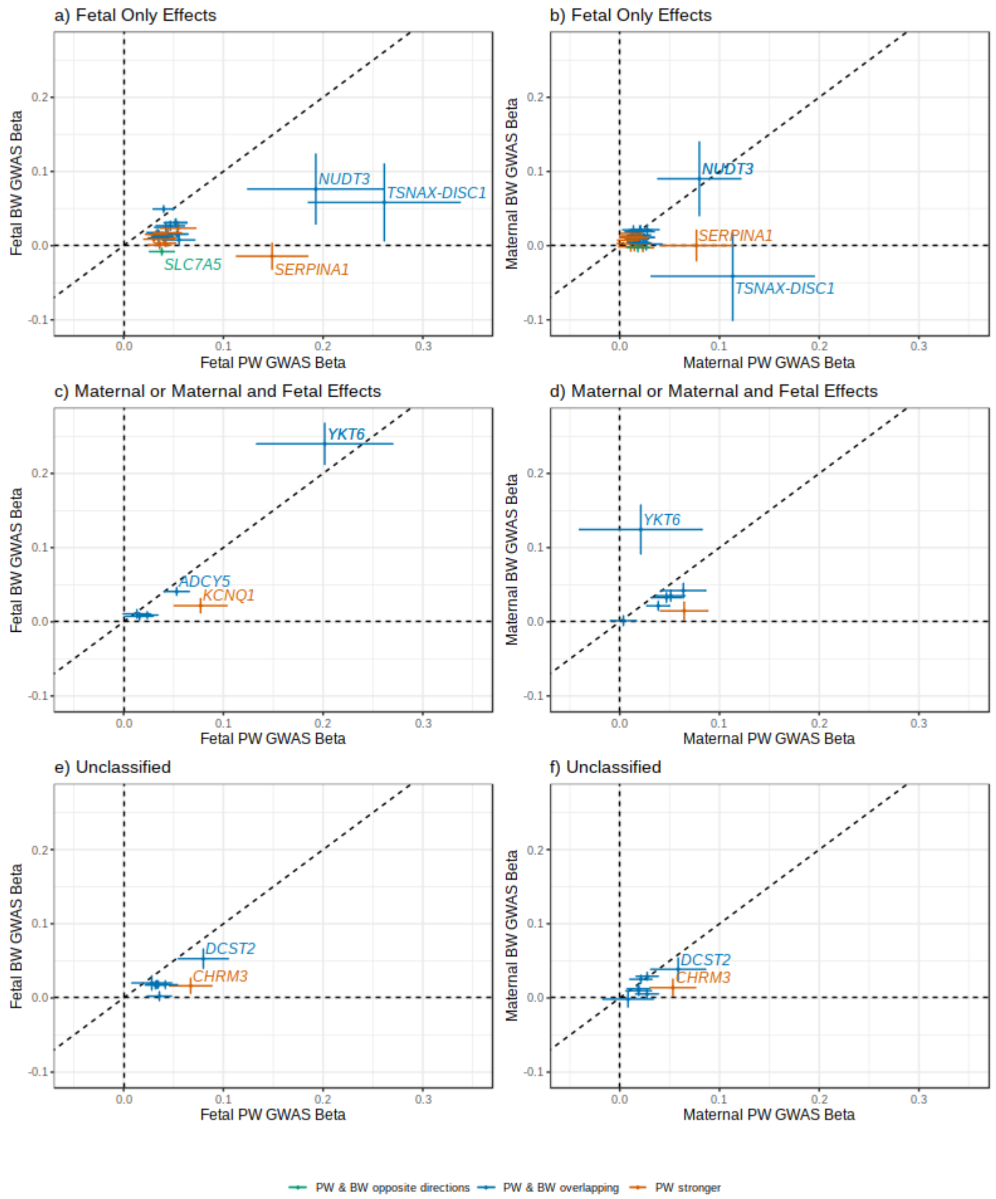

Scatter plots comparing effect size estimates from the PW GWAS with those from the BW GWAS[15]. The left column shows fetal genome associations, and the right shows maternal. The top row shows SNPs classified as fetal only effects on PW (Supplementary Table 7). The middle row shows SNPs classified as maternal, or maternal and fetal, and the bottom row shows unclassified SNPs. Colors indicate classifications, which are given in a key below the figure. Abbreviations: BW, birth weight; GWAS, genome-wide association study; PW, placenta weight. Error bars represent 95% confidence intervals.

**Supplementary Fig. 7: Tissue enrichment by mRNA data.**

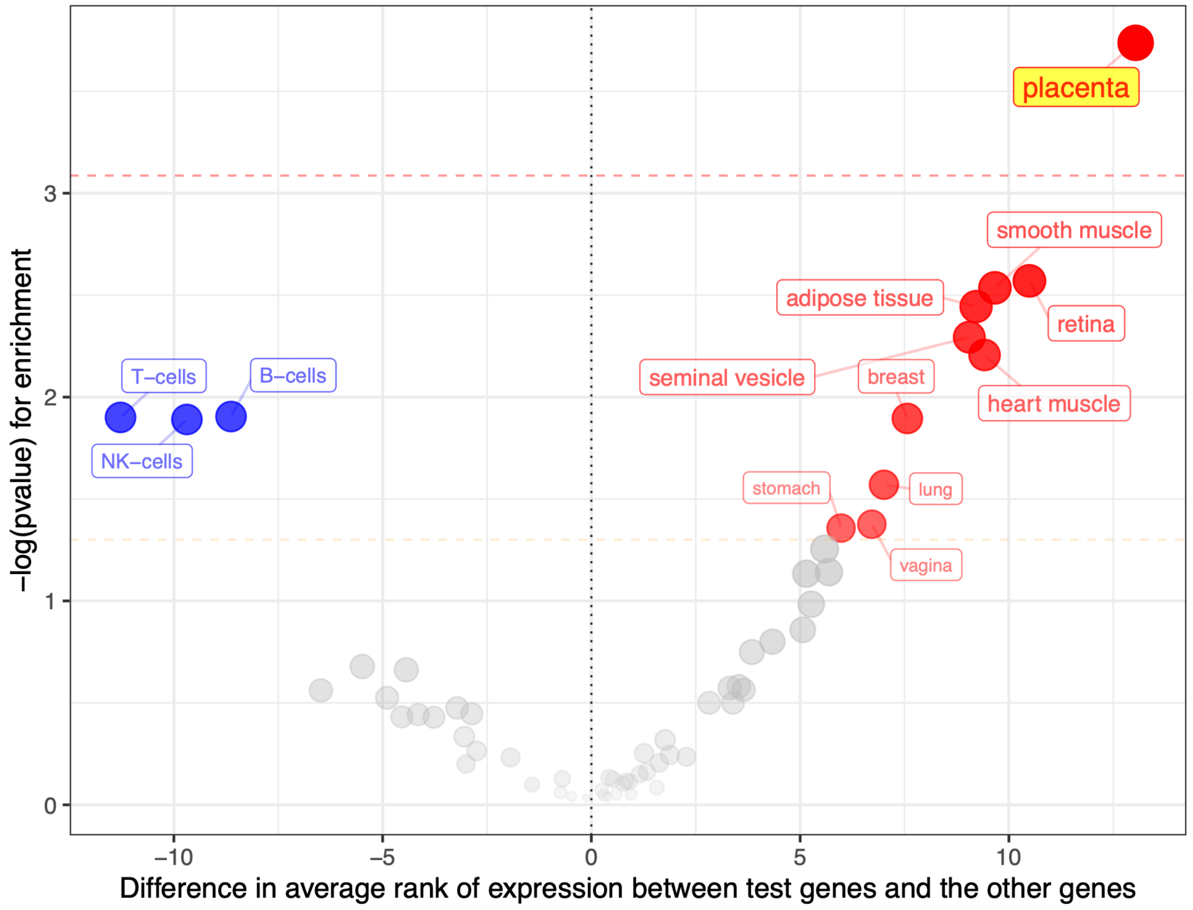

Plot illustrating the enrichment or depletion of RNA expression of the 31 placental weight associated protein-coding genes identified in the fetal GWAS, in 61 different tissues. Each dot represents a specific tissue and plots the difference in average rank of expression levels between the 31 placental weight associated genes and all the other genes (x-axis) with associated -log(*P* value) based on the Wilcoxon rank-sum test (y-axis). The two dashed horizontal lines represent significance levels with (red) or without (orange) Bonferroni correction ($n$ = 61). Tissues with nominally significant (*P* value < 0.05) higher or lower expression of the test genes are plotted and labeled as red or blue dots, respectively. Tissues with Bonferroni corrected significance are highlighted by labels with yellow background.

**Supplementary Fig. 8: Cell-type enrichment by scRNA-seq data.**

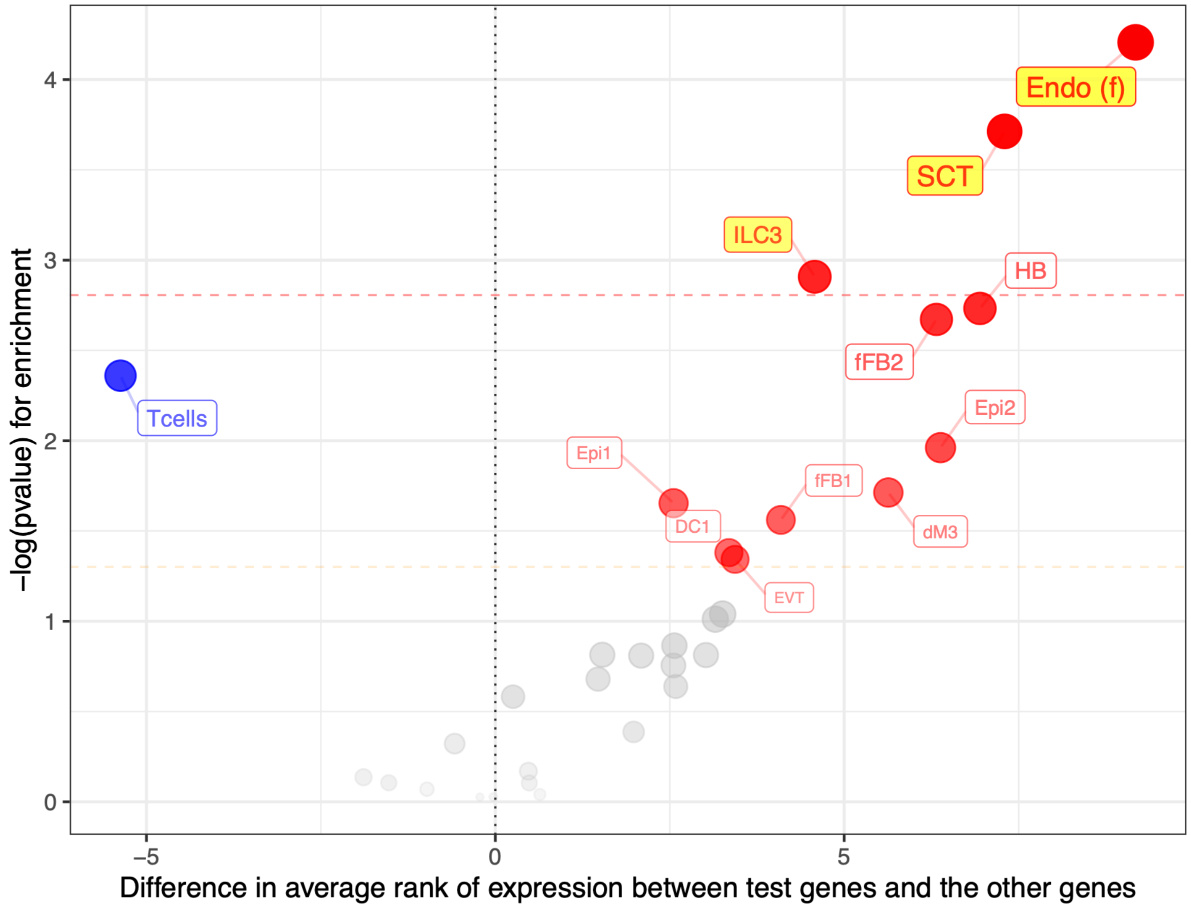

Plot illustrating the enrichment or depletion of RNA expression of the 31 nearest protein-coding genes at placental weight loci identified in the fetal GWAS in 32 different cell types at the early maternal-fetal interface. Each dot represents a specific cell-type and plots the difference in average rank of expression between the 31 placental genes and all the other genes (x-axis) with associated -log(*P* value) based on the Wilcoxon rank-sum test (y-axis). The two dashed horizontal lines represent significance levels with (red) or without (orange) after Bonferroni correction ($n$ = 32). Cell types with nominally significant (*P* value < 0.05) higher or lower expression of the test genes are plotted and labeled as red or blue dots, respectively. Cell types with Bonferroni corrected significance are highlighted by labels with yellow background. Abbreviations of cell types with nominally significant difference in expression: Endo (f), endothelial cells (fetal); SCT, syncytiotrophoblast (fetal); ILC, innate lymphocyte cells (maternal); HB, Hofbauer cells (fetal); fFB1 and fFB2, fibroblasts (fetal); Epi1 and Epi2, epithelial glandular cells (unassigned or maternal); dM3, Maternal macrophages (maternal cell in placenta); EVT, extravillous trophoblast; DC1, dendritic cells; Tcells, T cells (maternal or fetal).

**Supplementary Fig. 9a: Graphical display of methylated locus-specific association results, loci: rs723177, rs1655296, rs4953353, rs11708067, rs9817452, rs74457440**

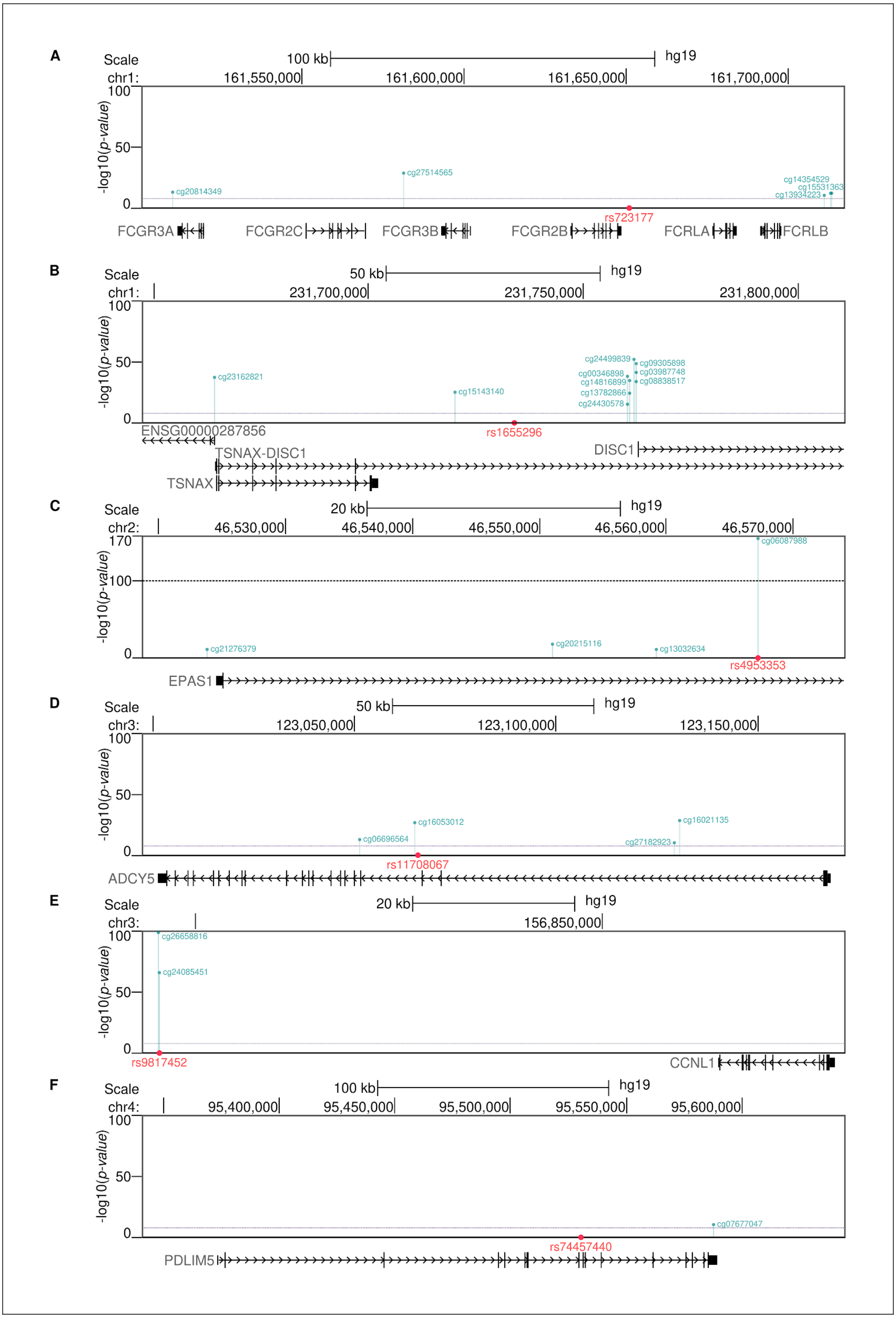
A
Scale
100 kb
hg19
chr1:
161,550,000
161,600,000
161,650,000
161,700,000
-log10(p-value)
100
50
0
cg20814349
cg27514565
cg14354529
cg15531363
cg13934223
rs723177
FCGR3A
FCGR2C
FCGR3B
FCGR2B
FCRLA
FCRLB
B
Scale
50 kb
hg19
chr1:
231,700,000
231,750,000
231,800,000
-log10(p-value)
100
50
0
cg23162821
cg15143140
cg24499839
cg09305898
cg00346898
cg03987748
cg14816899
cg08838517
cg13782866
cg24430578
rs1655296
ENSG00000287856
TSNAX-DISC1
DISC1
TSNAX
C
Scale
20 kb
hg19
chr2:
46,530,000
46,540,000
46,550,000
46,560,000
46,570,000
-log10(p-value)
170
100
0
cg06087988
cg21276379
cg20215116
cg13032634
rs4953353
EPAS1
D
Scale
50 kb
hg19
chr3:
123,050,000
123,100,000
123,150,000
-log10(p-value)
100
50
0
cg16053012
cg16021135
cg06696564
cg27182923
rs11708067
ADCY5
E
Scale
20 kb
hg19
chr3:
156,850,000
-log10(p-value)
100
50
0
cg26658816
cg24085451
rs9817452
CCNL1
F
Scale
100 kb
hg19
chr4:
95,400,000
95,450,000
95,500,000
95,550,000
95,600,000
-log10(p-value)
100
50
0
cg07677047
rs74457440
PDLIM5

Shown are associations between lead SNPs (red dots) and nearby differentially methylated CpG sites (blue lines) in placental tissue using 395 subjects from the Genetics of Glucose Regulation in Gestation and Growth study (Gen3G) and placental DNA methylation levels at 681 795 CpGs using Tensor QTL software[3]. Associations were tested within in a cis-window of 0.5 Mb of each lead SNP, and we illustrated methQTL with FDR threshold of ≤ 0.05 from the beta distribution by TensorQTL. Here, placental methQTLs at 6 of the 40 independent signals are illustrated. Abbreviations: Chr, chromosome; hg19, Human Genome version 19 corresponding to Genome Reference Consortium Human Build 37 (GRCh37); kb, kilo bases; Mb, mega bases.

**Supplementary Fig. 9b: Graphical display of methylated locus-specific association results, loci: rs3822394, rs541641049, rs9800506, rs12529634, rs11756568, rs1021508**

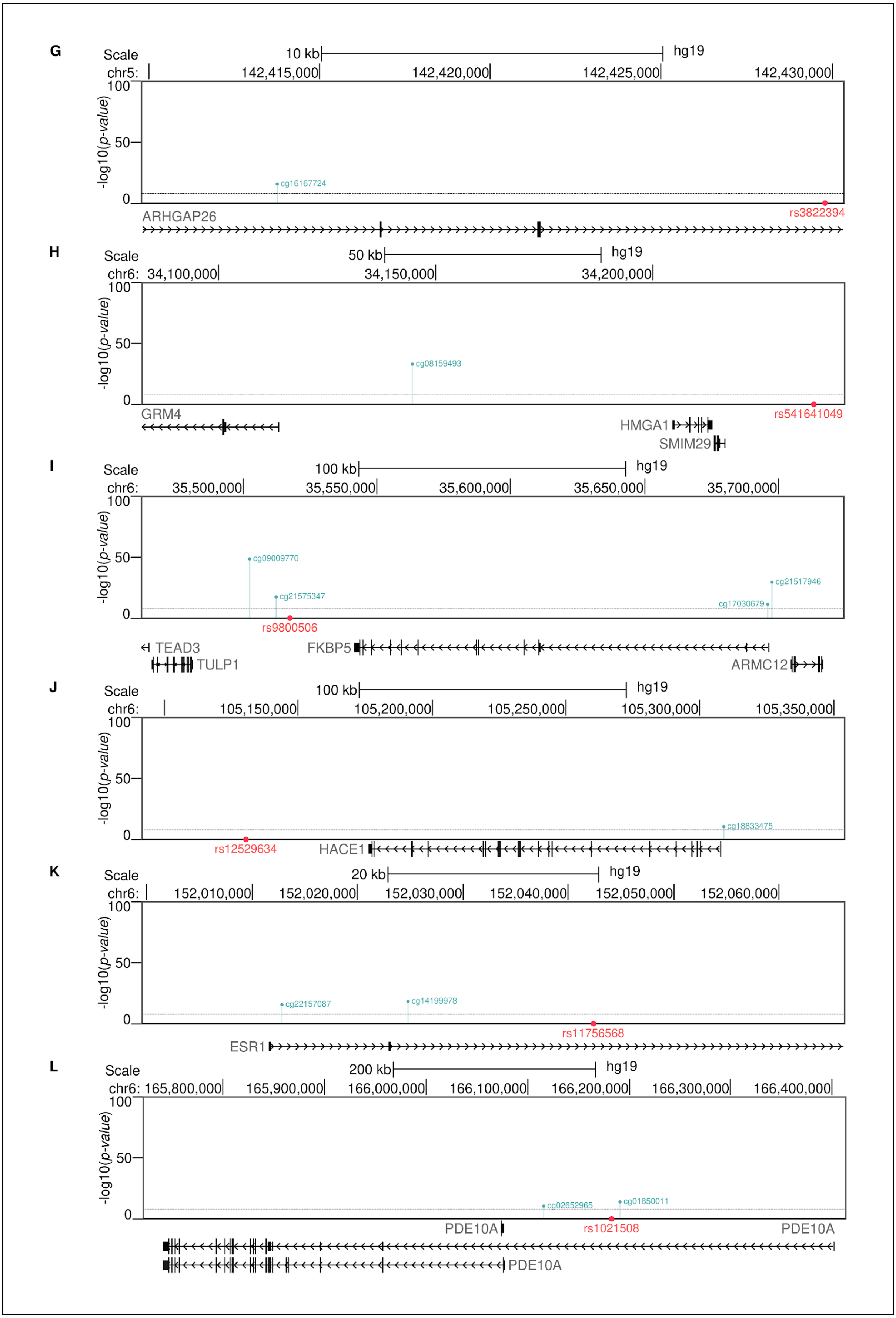
G
Scale
10 kb
hg19
chr5:
142,415,000
142,420,000
142,425,000
142,430,000
-log10(p-value)
cg16167724
rs3822394
ARHGAP26
H
Scale
50 kb
hg19
chr6:
34,100,000
34,150,000
34,200,000
-log10(p-value)
cg08159493
rs541641049
GRM4
HMGA1
SMIM29
I
Scale
100 kb
hg19
chr6:
35,500,000
35,550,000
35,600,000
35,650,000
35,700,000
-log10(p-value)
cg09009770
cg21575347
cg21517946
cg17030679
rs9800506
TEAD3
FKBP5
TULP1
ARMC12
J
Scale
100 kb
hg19
chr6:
105,150,000
105,200,000
105,250,000
105,300,000
105,350,000
-log10(p-value)
cg18833475
rs12529634
HACE1
K
Scale
20 kb
hg19
chr6:
152,010,000
152,020,000
152,030,000
152,040,000
152,050,000
152,060,000
-log10(p-value)
cg22157087
cg14199978
rs11756568
ESR1
L
Scale
200 kb
hg19
chr6: 165,800,000
165,900,000
166,000,000
166,100,000
166,200,000
166,300,000
166,400,000
-log10(p-value)
cg02652965
cg01850011
rs1021508
PDE10A
PDE10A
PDE10A

Shown are associations between lead SNPs (red dots) and nearby differentially methylated CpG sites (blue lines) in placental tissue using 395 subjects from the Genetics of Glucose Regulation in Gestation and Growth study (Gen3G) and placental DNA methylation levels at 681 795 CpGs using Tensor QTL software[3]. Associations were tested within in a cis-window of 0.5 Mb of each lead SNP, and we illustrated methQTL with FDR threshold of ≤ 0.05 from the beta distribution by TensorQTL. Here, placental methQTLs at 6 of the 40 independent signals are illustrated. Abbreviations: Chr, chromosome; hg19, Human Genome version 19 corresponding to Genome Reference Consortium Human Build 37 (GRCh37); kb, kilo bases; Mb, mega bases.

**Supplementary Fig. 9c: Graphical display of methylated locus-specific association results, loci: rs10486660, rs6557677, rs12543725, rs1801253, rs2168101, rs180534**

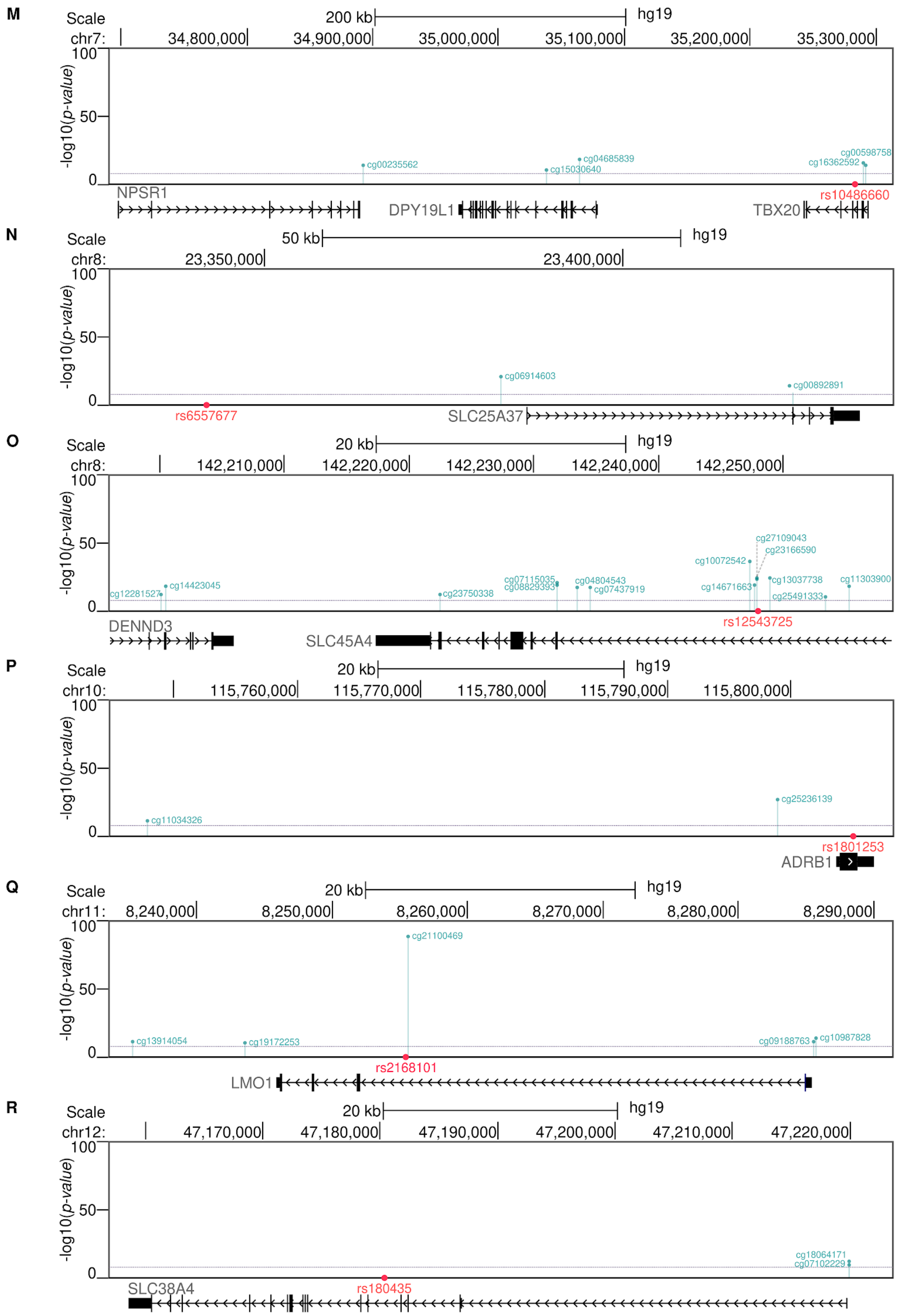
M
Scale
200 kb
hg19
chr7:
34,800,000
34,900,000
35,000,000
35,100,000
35,200,000
35,300,000
-log10(p-value)
100
50
0
cg00235562
cg15030640
cg04685839
cg16362592
cg00598758
rs10486660
NPSR1
DPY19L1
TBX20
N
Scale
50 kb
hg19
chr8:
23,350,000
23,400,000
cg06914603
cg00892891
rs6557677
SLC25A37
O
Scale
20 kb
hg19
chr8:
142,210,000
142,220,000
142,230,000
142,240,000
142,250,000
cg12281527
cg14423045
cg23750338
cg07115035
cg08829393
cg04804543
cg07437919
cg10072542
cg27109043
cg23166590
cg14671663
cg13037738
cg25491333
cg11303900
rs12543725
DENND3
SLC45A4
P
Scale
20 kb
hg19
chr10:
115,760,000
115,770,000
115,780,000
115,790,000
115,800,000
cg11034326
cg25236139
rs1801253
ADRB1
Q
Scale
20 kb
hg19
chr11:
8,240,000
8,250,000
8,260,000
8,270,000
8,280,000
8,290,000
cg21100469
cg13914054
cg19172253
cg09188763
cg10987828
rs2168101
LMO1
R
Scale
20 kb
hg19
chr12:
47,170,000
47,180,000
47,190,000
47,200,000
47,210,000
47,220,000
cg18064171
cg07102229
SLC38A4
rs180435

Shown are associations between lead SNPs (red dots) and nearby differentially methylated CpG sites (blue lines) in placental tissue using 395 subjects from the Genetics of Glucose Regulation in Gestation and Growth study (Gen3G) and placental DNA methylation levels at 681 795 CpGs using Tensor QTL software[3]. Associations were tested within in a cis-window of 0.5 Mb of each lead SNP, and we illustrated methQTL with FDR threshold of ≤ 0.05 from the beta distribution by TensorQTL. Here, placental methQTLs at 6 of the 40 independent signals are illustrated. Abbreviations: Chr, chromosome; hg19, Human Genome version 19 corresponding to Genome Reference Consortium Human Build 37 (GRCh37); kb, kilo bases; Mb, mega bases.

**Supplementary Fig. 9d: Graphical display of methylated locus-specific association results, loci: rs7177338, rs55958435, rs876987**

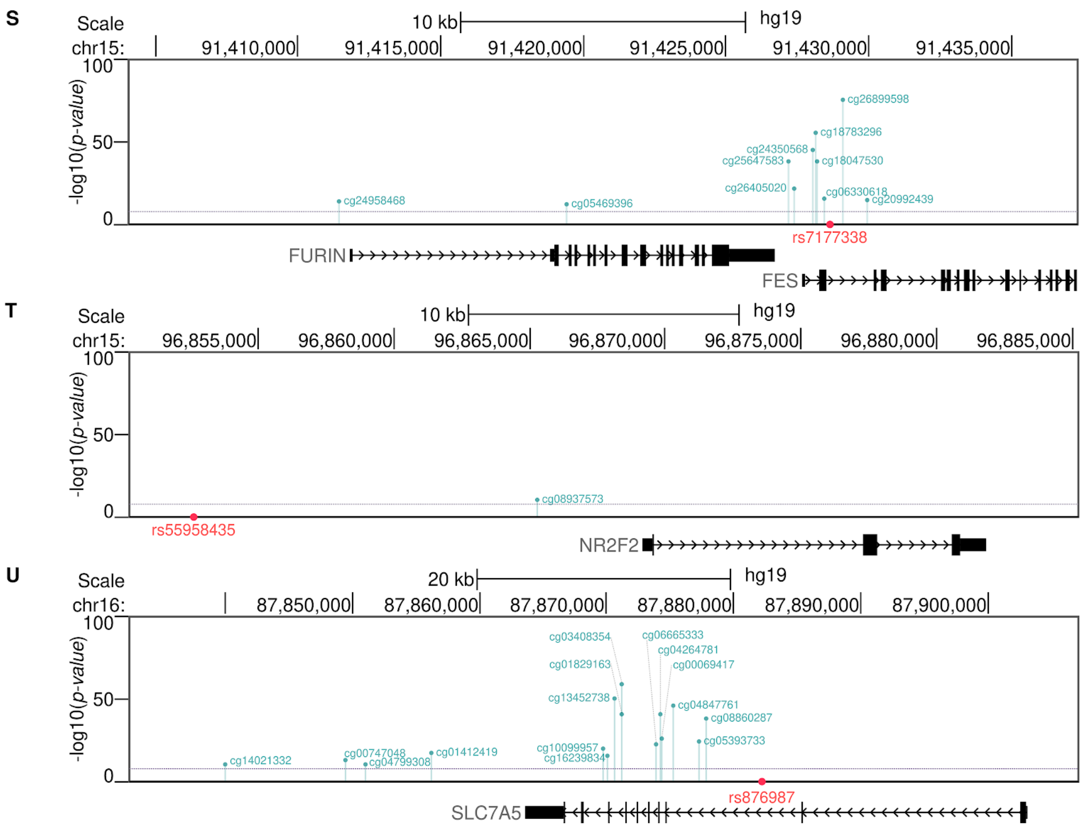

Shown are associations between lead SNPs (red dots) and nearby differentially methylated CpG sites (blue lines) in placental tissue using 395 subjects from the Genetics of Glucose Regulation in Gestation and Growth study (Gen3G) and placental DNA methylation levels at 681 795 CpGs using Tensor QTL software[3]. Associations were tested within in a cis-window of 0.5 Mb of each lead SNP, and we illustrated methQTL with FDR threshold of ≤ 0.05 from the beta distribution by TensorQTL. Here, placental methQTLs at 3 of the 40 independent signals are illustrated. Abbreviations: Chr, chromosome; hg19, Human Genome version 19 corresponding to Genome Reference Consortium Human Build 37 (GRCh37); kb, kilo bases; Mb, mega bases.

**Supplementary Fig. 10a-l: Quantile–quantile (QQ) plots of observed versus expected –$\log_{10}$ *P* values, various pregnancy and perinatal traits.**

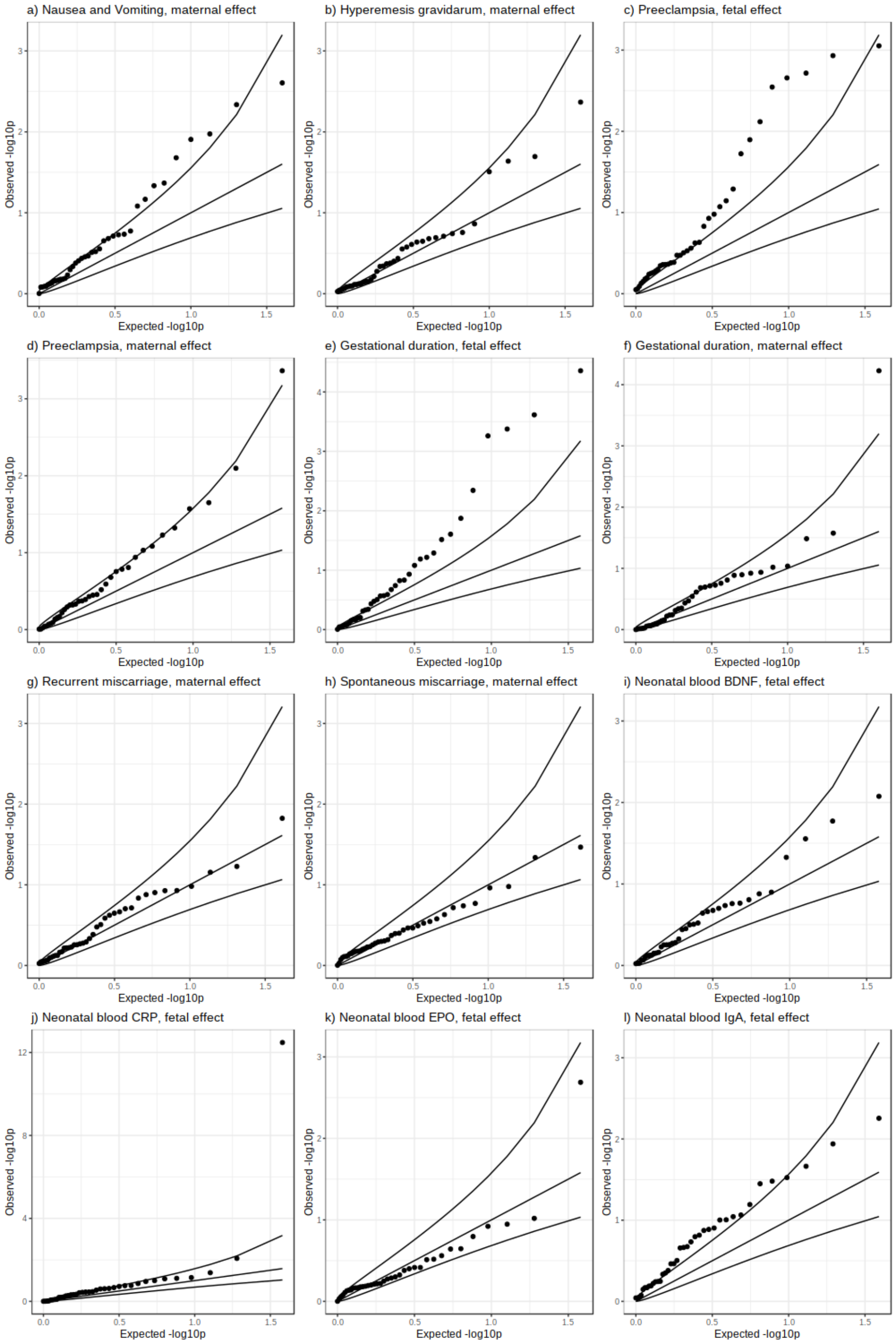

Shown are QQ plots of associations between 40 independent lead placental weight SNPs and various pregnancy and perinatal traits from published GWAS studies (**a-l**). The lines represent the expected, and upper- and lower-95% confidence intervals under the null distribution. Nausea and Vomiting of Pregnancy is abbreviated to Nausea and Vomiting.

**Supplementary Fig. 10m-u: Quantile–quantile (QQ) plots of observed versus expected –log$_{10}$ *P* values, additional pregnancy and perinatal traits.**

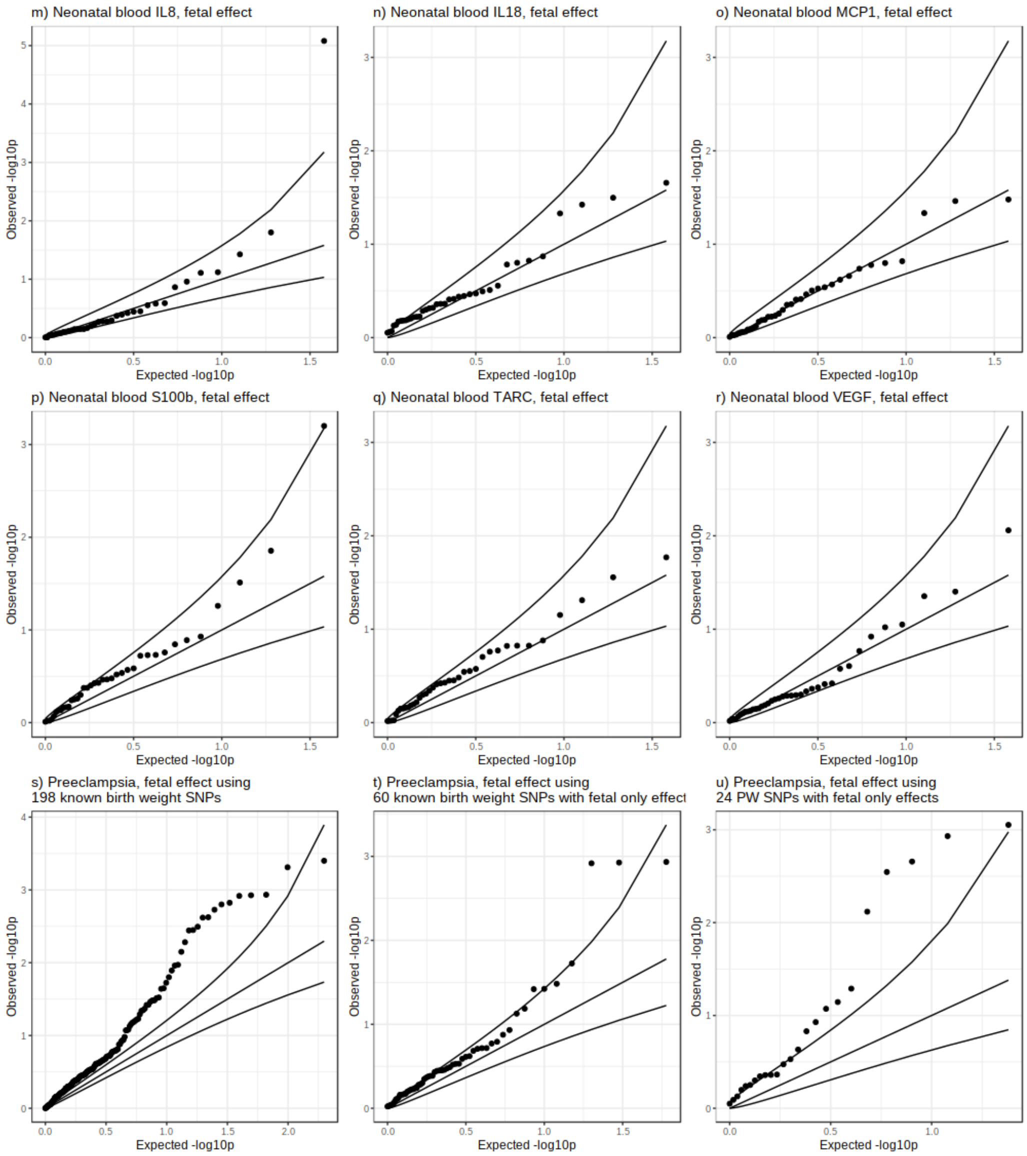

Shown are QQ plots of associations between 40 independent lead placental weight SNPs and various pregnancy and perinatal traits from published GWAS studies (**m-r**), and between 198 birth weight associated SNPs and preeclampsia (**s**), 60 birth weight SNPs with fetal effects only and preeclampsia (**t**), and 24 PW SNPs with fetal effects only and preeclampsia (**u**). The lines represent the expected, and upper- and lower-95% confidence intervals under the null distribution.

**Supplementary Fig. 11: Scatter plots.**

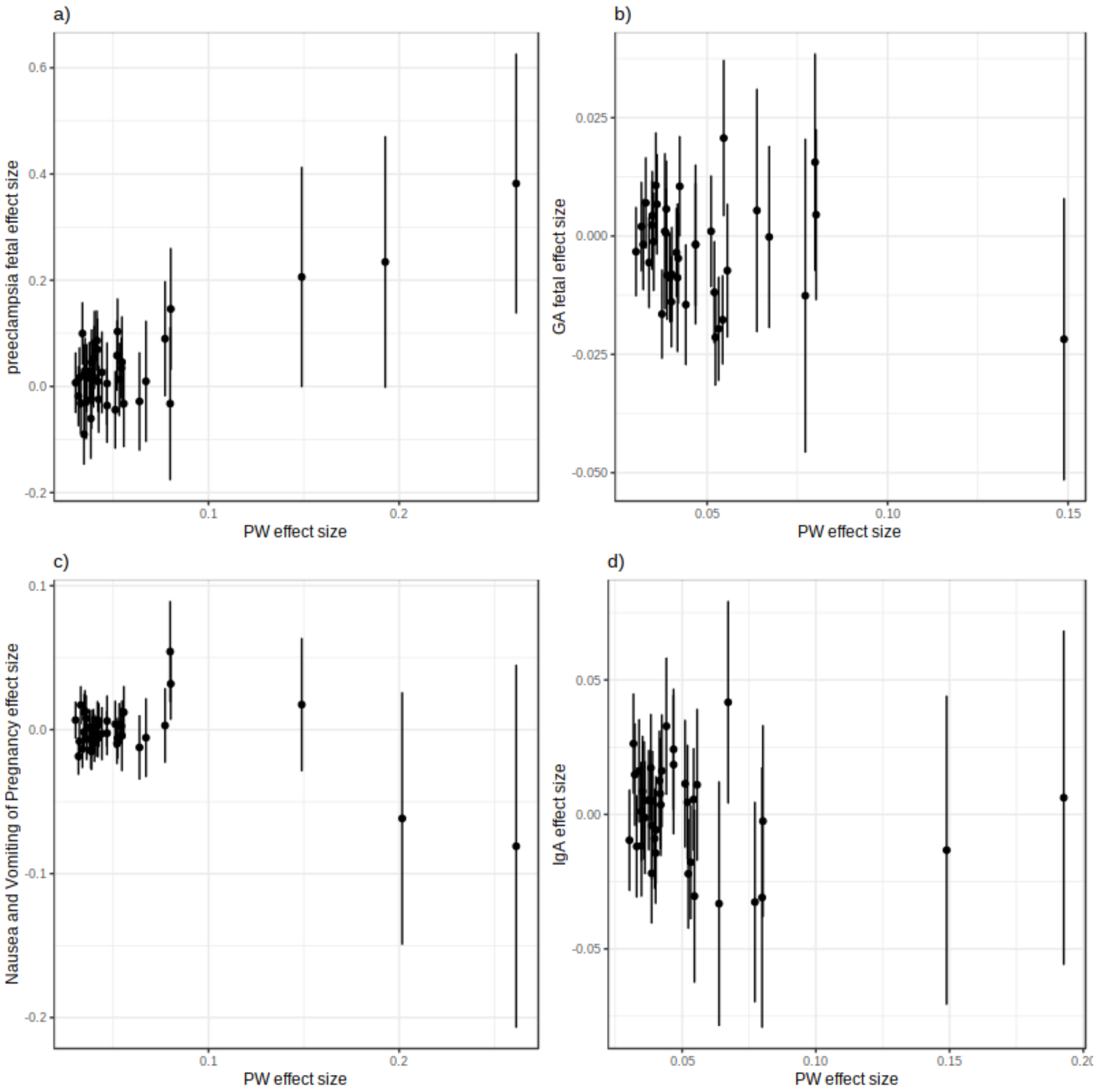

Shown are effects (y-axis) of lead placental weight SNPs on preeclampsia (**a**, fetal effect), gestational duration (**b**, fetal effect), morning sickness (**c**, maternal effect), and neonatal IgA levels (**d**, fetal effect) vs. effect on placental weight (from fetal GWAS; x-axis). Y-error bars show 95% CI. Abbreviation: PW, placental weight.

**Supplementary Fig. 12: Mendelian randomization analyses.**

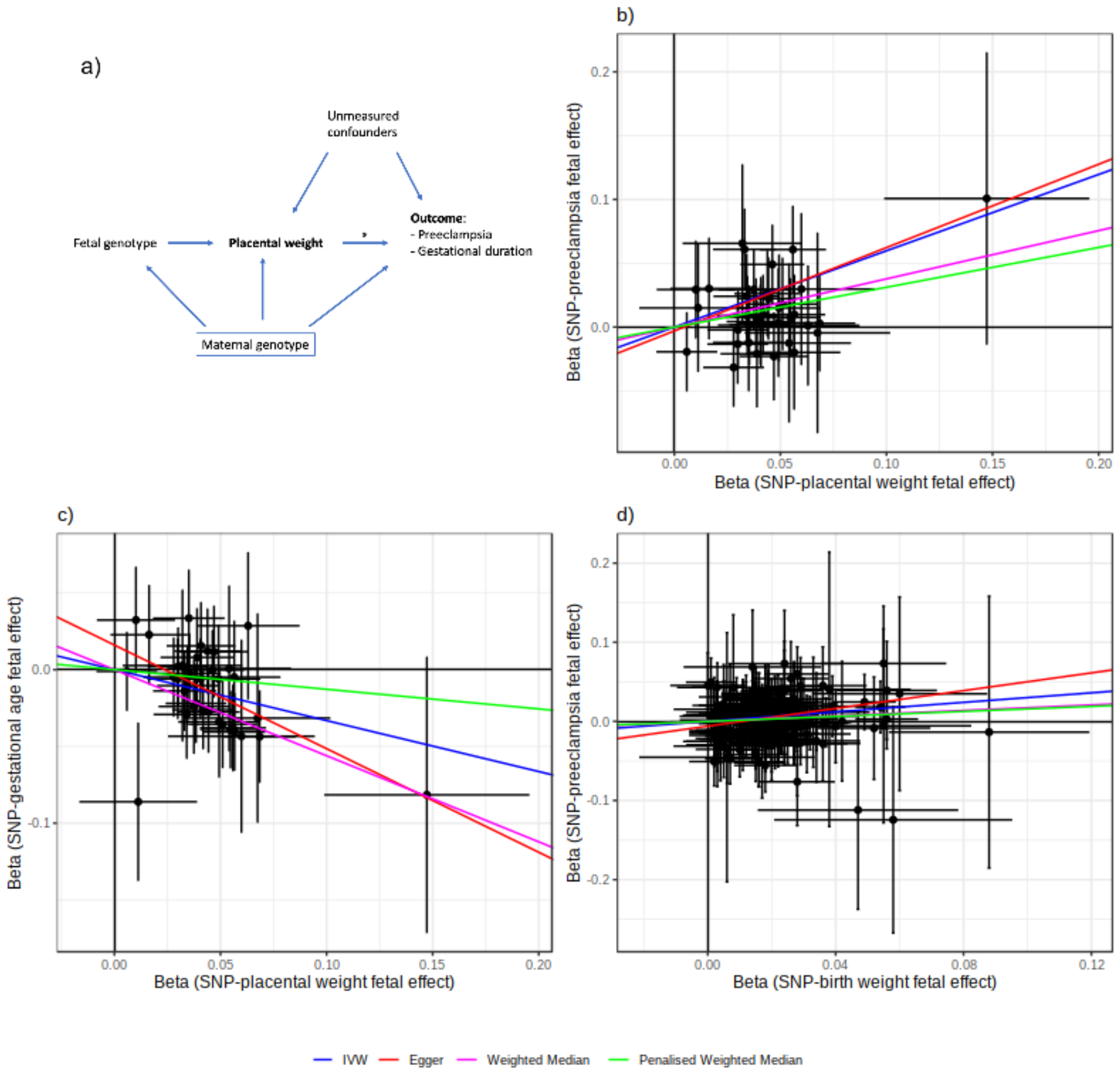

**a**, Diagram illustrating the Mendelian randomization analyses used to test for a causal relationship (*) between higher placental weight (exposure; a proxy for faster placental growth) and either preeclampsia, or gestational duration (outcomes). Key assumptions are (i) that the fetal genotype (genetic instrumental variable) is robustly related to placental weight, (ii) that potential confounders of the causal relationship of interest are not associated with the genetic instrumental variable, and (iii) that the genetic instrumental variable can only be related to the outcome via its effect on the exposure (placental weight), and not through any other pathway. Since maternal genotype is correlated with fetal genotype and may additionally influence placental weight and the outcome variables, it is a potential confounder and should be adjusted for in the analyses (indicated by the box around it). We were able to adjust for maternal genotype using weighted linear model (WLM) estimates of fetal genetic effects on placental weight and on preeclampsia and gestational duration, since both maternal and fetal GWAS summary statistics are available for those outcomes. To check for deviation from assumption (iii) above, we used the MR Egger, weighted median and penalized weighted median sensitivity analyses. **b-d**, Results of two-sample Mendelian randomization analyses testing the effect of (**b**) higher placental weight using fetal genetic instruments on preeclampsia, (**c**) higher placental weight using fetal genetic instruments on gestational duration, and (**d**) higher birth weight using fetal genetic instruments on preeclampsia.

**Supplementary Fig. 13: Polygenic score analyses.**

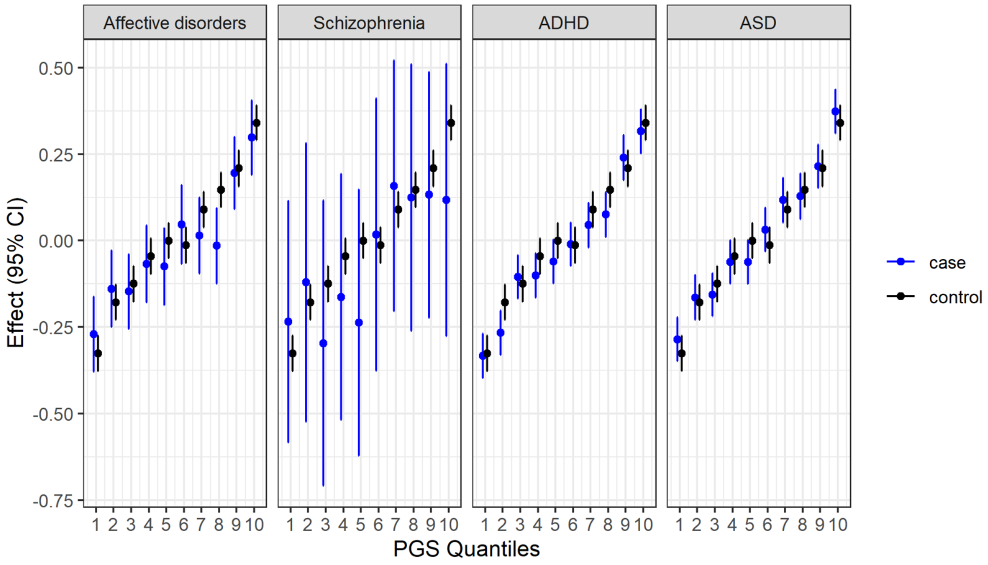

The panels show associations between quantiles of fetal polygenic scores for placental weight and standardized observed placental weight in the iPSYCH cohort. Black dots show associations for the population representative sample used as controls in iPSYCH, and blue dots show associations for cases of four different neuropsychiatric diseases. Abbreviations: CI, confidence interval; ADHD, attention deficit/hyperactivity disorder; ASD, autism spectrum disorder; PGS, polygenic score.

**Supplementary Fig. 14: Analyses in the iPSYCH cohort of placental weight and risk of neuropsychiatric diseases.**

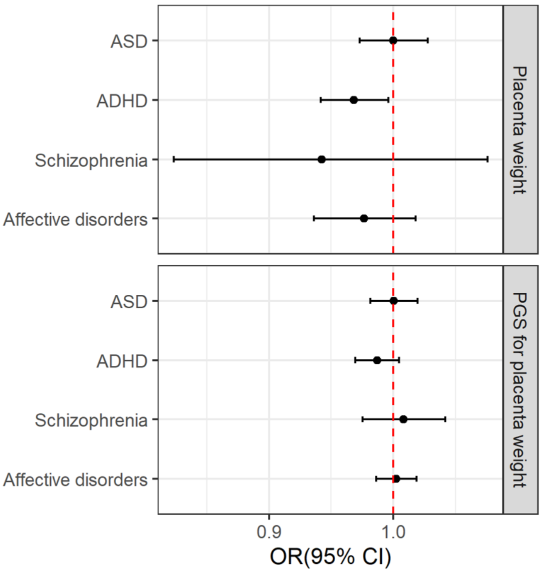

The figure shows odds ratios (ORs) from logistic regressions of four neuropsychiatric diseases on standardized observed placental weight (upper panel) and fetal polygenic score of placental weight (lower panel). The ORs correspond to a change of one standard deviation in standardized observed placental weight or PGS for placental weight. Abbreviations: CI, confidence interval; ADHD, attention deficit/hyperactivity disorder; ASD, autism spectrum disorder; OR, odds ratio; PGS, polygenic score.

**Supplementary Fig. 15: Genomic SEM model**

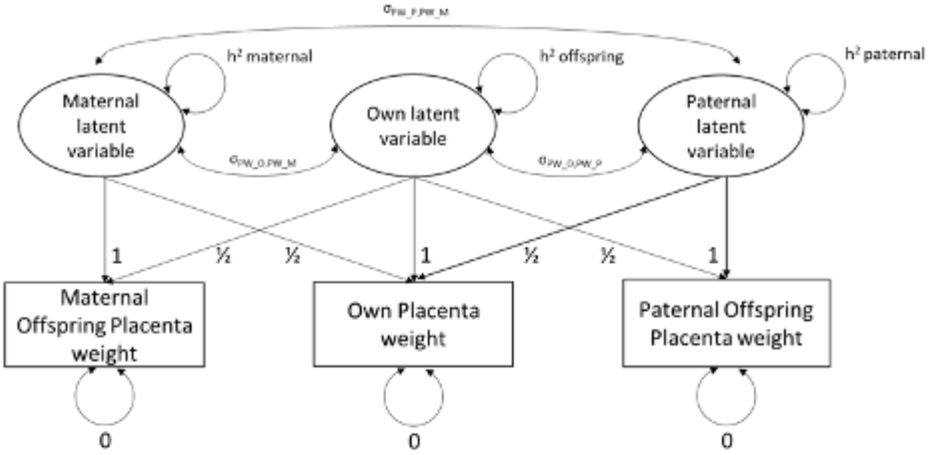

GWAS summary results statistics from the three placental weight (squares) are modeled in terms of latent variables representing the fetal genome, the maternal genome and the paternal genome (circles). The lower part of this model reflects simple biometrical genetics principles (i.e. the fact that offspring and maternal/paternal genomes are correlated 0.5) and consists of path coefficients fixed to the value of one or one half. The top half of the model consists of free parameters requiring estimation- three SNP heritabilities (maternal, paternal and own), and three genetic covariances between the different variables, representing commonalities in gene action across the maternal, paternal and fetal genomes.

**Supplementary Fig. 16: Enrichment test using tissue or cell-type specific expression data.**

Step 1. Rank normalization across all tissue/cell types

| | Gene 1 | Gene 2 | .... | Gene *g* |
|---|---|---|---|---|
| Tissue/Cell Type 1 | $x_{1,1}$ | $x_{1,2}$ | | $x_{1,g}$ |
| Tissue/Cell Type 2 | $x_{2,1}$ | $x_{2,2}$ | | $x_{2,g}$ |
| Tissue/Cell Type t | $x_{t,1}$ | $x_{t,2}$ | | $x_{t,g}$ |

Step 2. Enrichment test in a tissue/cell type based on rank

| | Gene 1 | Gene 2 | .... | Gene *g* |
|---|---|---|---|---|
| Tissue/Cell Type 1 | $r_{1,1}$ | $r_{1,2}$ | | $r_{1,g}$ |
| Tissue/Cell Type 2 | $r_{2,1}$ | $r_{2,2}$ | | $r_{2,g}$ |
| Tissue/Cell Type t | $r_{t,1}$ | $r_{t,2}$ | | $r_{t,g}$ |

Our gene expression enrichment analysis included two steps. First, for each gene (red box), we rank normalized the expression levels of a reference data set ($x_{i,j}$: expression level of gene *j* in tissue or cell-type *i*) to $r_{i,j}$. For any gene *j*, $r_{i,j}$ takes a value from 1 to *t*, indicating the order of gene *j*'s expression level in tissue or cell-type *i* among *t* different tissue or the cell-types. In the second step, for a specific tissue or cell-type under examination, the ranks of gene expression of a set of test genes (blue box) were compared against all other genes (green box) using Wilcoxon rank-sum test.

### Supplementary Tables

**Supplementary Table 1: Description of studies (fetal); cohort data.**

Descriptive phenotypic statistics for studies contributing to fetal GWAS meta-analysis: country of origin, years of birth, sample size, phenotypic exclusion, data collection data, mean (SD) or placental weight, median and interquartile range of gestational age at delivery, and study references. The sample sizes reported are the total number of individuals who contributed to any analysis.

**Supplementary Table 2: Description of studies (fetal); genetics.**

Descriptive genotyping statistics for studies contributing to fetal GWAS meta-analysis: genotyping, quality control, pre-phasing, imputation, and association analysis.

**Supplementary Table 3: Description of studies (maternal); cohort data.**

Descriptive phenotypic statistics for studies contributing to maternal GWAS meta-analysis: country of origin, years of birth, sample size, phenotypic exclusion, data collection data, mean (SD) or placental weight, median and interquartile range of gestational age at delivery, and study references. The sample sizes reported are the total number of individuals who contributed to any analysis.

**Supplementary Table 4: Description of studies (maternal); genetics.**

Descriptive genotyping statistics for studies contributing to maternal GWAS meta-analysis: genotyping, quality control, pre-phasing, imputation, and association analysis.

**Supplementary Table 5: Description of studies (paternal); cohort data.**

Descriptive phenotypic statistics for studies contributing to paternal GWAS meta-analysis: country of origin, years of birth, sample size, phenotypic exclusion, data collection data, mean (SD) or placental weight, median and interquartile range of gestational age at delivery, and study references. The sample sizes reported are the total number of individuals who contributed to any analysis.

**Supplementary Table 6: Description of studies (paternal); genetics.**

Descriptive genotyping statistics for studies contributing to paternal GWAS meta-analysis: genotyping, quality control, pre-phasing, imputation, and association analysis.

**Supplementary Table 7: Summary statistics from the genome-wide association analyses of fetal, maternal, and paternal genotype with birth weight.**

Summary statistics for loci reaching $P < 5\text{x}10^{-8}$ in any of the fetal, maternal or paternal genome-wide association meta-analyses of placental weight, including details of birth weight or gestational age loci within 500 kb of the placental weight lead SNP, placental eQTL lookups, meQTL lookups, and results from classifications of loci into placental weight only, or showing evidence of association with both placental weight and birth weight. Colocalisation results showing posterior probabilities of placental weight and birth weight lead SNPs tag the same signal (column AL) or separate association signals (column AK) are shown in bold where posterior probabilities as > 0.8 for either shared or separate signals.

**Supplementary Table 8: Association results for the lead SNPs from Supplementary Table 7 in the sex-only adjusted meta-analysis.**

Summary statistics from the 41 lead SNPs from Supplementary Table 7 in the sex-only adjusted meta-analysis, along with details of the lead SNP in the sex-only adjusted meta-analysis if this is different to the sex and gestational age adjusted meta-analysis lead SNP.

**Supplementary Table 9: Estimates of variance (heritability) and correlation between the maternal, paternal and fetal effect using the genomic SEM model.**

Estimates of variance (heritability) of the maternal effect, paternal effect and fetal effect on placental weight and the correlation between the three variables using the meta-analysis GWAS of placental weight and obtaining direct effects using the model depicted in Supplementary Fig. 14.

**Supplementary Table 10: Summary statistics from the WLM and parent-offspring conditional analysis in MoBa for the 41 lead SNPs.**

Association summary statistics from the fetal, maternal, and paternal meta-analyses, WLM, and parent-offspring conditional analysis in the MoBa cohort for the 41 lead SNPs from Supplementary Table 7. Trio WLM refers to the WLM allowing for non-zero paternal effects, POE WLM refers to the WLM where paternal effect are set to zero, and instead the paternal GWAS is used to estimate parent-of-origin effects, and Pair WLM refers to the WLM only allowing for maternal and fetal effects. MoBa GWAS shows results from the GWAS in Moba, MoBa Conditional Trio Analysis shows the results of multivariable analysis regressing maternal, paternal and fetal genotype on placental weight simultaneously, and MoBa Conditional Duo Analysis shows the results of multivariable analysis regressing maternal and fetal genotype on placental weight. Maternal fetal classification shows classifications when comparing betas and 95% CIs for fetal and maternal associations in the WLM and MoBa as described in the methods.

**Supplementary Table 11: Haplotypes based on MobBa data compared to published study by Juliusdottir *et al.*[1]**

Closest imprinted genes to the 41 lead SNPs according to www.geneimprint.com. Summary statistics of the (child, mother, father) models run in the Norwegian Mother, Father and Child (MoBa) cohort for placental and birth weight, as well as mode of association results for these variants from Juliusdottir *et al.*[1]

**Supplementary Table 12: Ranks of gene expression of the 31 placental weight associated genes with fetal effect in placenta and the three cell-types reached multiple-testing corrected significance level in enrichment analysis.**

Ranks of gene expression levels of the 31 genes (the closest protein-coding genes to the index SNP of the genomic loci with fetal effect on placental weight) in placenta and the three cell-types reached multiple-testing corrected significance level in enrichment test (please see Supplementary Figure 8 and 9). For a particular gene, the rank is the order of expression level in a specific tissue or cell-type among all tissues ($n$ = 61) or cell-types ($n$ = 32). For example, *FCGR2B* ranked 1 in placenta means the expression of *FCGR2B* is the highest in placenta among the 61 tissues. The lowest rank is 61 in tissue-specific analysis (placenta) and 32 in cell-type analysis (last three columns). Abbreviations for the three significant cell-types: Endo (f), endothelial cells (fetal); SCT, syncytiotrophoblast (fetal); ILC, innate lymphocyte cells (maternal).

#### Supplementary Table 13: Annotation of SNPs at the placental weight loci with respect to GWAS Catalog associations.

Associations from the GWAS Catalog are restricted to SNPs in LD ($r^2 > 0.6$) with a lead placental weight SNP and with $P < 5 \times 10^{-8}$ reported in the GWAS Catalog.

#### Supplementary Table 14: Annotation of SNPs at the placental weight loci with respect to expression quantitative trait loci (eQTL).

The eQTL results are based on the PsychENCODE, BIOSQTL, EyeGEx, DICE, eQTL Catalogue, eQTLGen, and GTEx/v8 databases.

#### Supplementary Table 15: Annotation of SNPs at the placental weight loci with respect to protein quantitative trait loci (pQTL).

The pQTL results are based on Sun *et al*.[4],Suhre *et al*.[5], Emilsson *et al*.[6], and Kilpeläinen *et al*.[7].

#### Supplementary Table 16: Lookups of placental weight lead SNPs in external GWAS of preeclampsia, recurrent and spontaneous miscarriage, fetal cytokines, birth weight, gestational age, hyperemesis/nausea and vomiting of pregnancy, and endometriosis.

Associations statistics for placental weight lead SNPs in external GWASs of preeclampsia, recurrent and spontaneous miscarriage, fetal cytokines, birth weight, gestational age, hyperemesis/nausea and vomiting of pregnancy, and endometriosis.

#### Supplementary Table 17: Mendelian Randomization analyses for effects of placental and birth weight on pregnancy outcomes.

Mendelian randomization analyses of the effects of placental weight and birth weight on pregnancy outcomes of gestational age and preeclampsia risk. Fetal effect refers to the partitioned fetal effect derived using the WLM. Effect sizes are units of outcome (column C) per SD of placental weight or birth weight. Egger regression[8] provides a sensitivity analysis to test for the effects of invalid instruments. The intercept term gives an estimate of directional pleiotropy in the instruments. Weighted median and penalized weighted median are sensitivity analyses which provide valid estimates when some instruments are invalid[9] .

#### Supplementary Table 18: Mendelian Randomization analyses for effects of maternal exposures on placental weight.

Mendelian randomization analyzes of the effects of height, fasting glucose, insulin secretion, insulin resistance, systolic blood pressure, and diastolic blood pressure on placental weight. Effect sizes are g of placental weight per unit of exposure. Egger regression[8] provides a sensitivity analysis to test for the effects of invalid instruments. The intercept term gives an estimate of directional pleiotropy in the instruments. Weighted median and penalized weighted median are sensitivity analyses which provide valid estimates when some instruments are invalid[9].

### Acknowledgements

The content of this manuscript and supplementary material is solely the responsibility of the authors and does not necessarily represent the official views of the funders.

#### Personal funding of coauthors

R.N.B. and R.M.F. were funded by a Wellcome Trust and Royal Society Sir Henry Dale Fellowship (WT104150). G.-H.M. is the recipient of an Australian Research Council Discovery Early Career Award (Project number: DE220101226) funded by the Australian Government and supported by the Research Council of Norway (Project grant: 325640). L.S. received support from a Carlsberg Foundation postdoctoral fellowship (CF15-0899). K.B. acknowledges the Novo Nordisk Foundation (grants NNF17OC0027594 and NNF14CC0001). C.Albiñana was supported by the Danish National Research Foundation, via a Niels Bohr Professorship to John McGrath. J.R., S.Sebert and M.-R.J. acknowledge the European Union's Horizon 2020 research and innovation program under grant agreements No 874739 (LongITools), 733206 (LifeCycle) and 633595 (DynaHEALTH), and the Academy of Finland under grant agreement No 285547 (EGEA). M.-R.J. acknowledges the European Union's Horizon 2020 research and innovation program under grant agreements No. 825762 (EDCMET) and EarlyCaur (848158). M.-R.J. also acknowledges the Joint Programming Initiative a Healthy Diet for a Healthy Life (JPI HDHL) (PREcisE proposal 655), the Medical Research Council (MRC) and the Biotechnology and Biological Sciences Research Council (BBSRC) [grant reference: MR/S03658X/1]. D.W. acknowledges the Novo Nordisk Foundation (grants NNF17OC0027594 and NNF14CC0001). C.S.S. is supported by a PhD studentship award from the Medical Research Council (MR/N013166/1). D.J.L. and R.M.F. are funded by a Wellcome Trust Senior Research Fellowship (WT220390). P.-É.J. is a J2 research scholar from the Fonds de la recherche du Québec en santé (FRQS) and members of the CR-CHUS, a FRQS-funded Research Center. L.Bouchard is a senior research scholar from the Fonds de la recherche du Québec en santé (FRQS) and members of the CR-CHUS, a FRQS-funded Research Center. H.C. and K.Hao were supported by NIH/NIEHS R01 ES029212. L.Bhatta and B.M.B. work in a research unit funded by Stiftelsen Kristian Gerhard Jebsen; Faculty of Medicine and Health Sciences, Norwegian University of Science and Technology (NTNU); the Liaison Committee for Education, Research, and Innovation in Central Norway; and the Joint Research Committee between St. Olavs Hospital and the Faculty of Medicine and Health Sciences, NTNU. S.B. acknowledges the Novo Nordisk Foundation (grants NNF17OC0027594 and NNF14CC0001). R.G. received funding from the Dutch Diabetes Foundation (grant number 2017.81.002), the Netherlands Organization for Health Research and Development (NWO, ZonMW, grant number 543003109, grant number 09150172110034) and from the European Union's Horizon 2020 research and innovation programme under the ERA-NET Cofund action (no 727565), EndObesity, ZonMW the Netherlands (no. 529051026). V.W.V.J. received a Consolidator Grant from the European Research Council (ERC-2014-CoG-648916). F.R. was supported by LEGENDARE ERC-ADG 2020 101021500. M.Vaarasmaki was supported by the Finnish Government Subsidiary for Health Care Research. B.J.V. was supported by a Lundbeck Foundation Fellowship (R335-2019-2339). A.T.H. was supported by a NIHR Senior Investigator award and also a Wellcome Trust Senior Investigator award (098395/Z/12/Z). D.A.L.'s contribution was supported by The European Research Council under the European Union's Horizon 2020 research and innovation program (grant agreement No 101021566), The British Heart Foundation (CH/F/20/90003 and AA/18/7/34219) and Medical Research Council (MC_UU_00011/6). D.M.E. is funded by an Australian National Health and Medical Research Council Senior Research Fellowship (APP1137714) and NHMRC project grants (GNT1157714, GNT1183074). B.J. received funding from The Swedish Research Council, Stockholm, Sweden (2019-01004), The Research Council of Norway, Oslo, Norway (FRIMEDBIO #547711), The Swedish Research Council, Stockholm, Sweden (2015-02559). G.Z. is supported by a grant from the Eunice Kennedy Shriver National Institute Of Child Health & Human Development of the National Institutes of Health under Award Number R01HD101669, a grant from the Burroughs Wellcome Fund (Grant 10172896), the March of Dimes Prematurity Research Center Ohio Collaborative and the Bill & Melinda Gates Foundation. S.J. was supported by Helse Vest's Open Research Grant (grants #912250 and F-12144), the Novo Nordisk Foundation (NNF20OC0063872) and the Research Council of Norway (grant #315599). B.F. received support from Independent Research Fund Denmark (0134-00244B) and from the Oak Foundation (OCAY-18-598). P.R.N. was supported by grants from the European Research Council (AdG SELECTionPREDISPOSED #293574), the Bergen Research Foundation ("Utilizing the Mother and Child Cohort and the Medical Birth Registry for Better Health"), Stiftelsen Kristian Gerhard Jebsen (Translational Medical Center), Trond Mohn Stiftelsen (Mohn Center for Diabetes Precision Medicine), the University of Bergen, the Research Council of Norway (FRIPRO grant #240413), the Western Norway Regional Health Authority (Strategic Fund "Personalized Medicine for Children and Adults"), the Novo Nordisk Foundation (grant #54741), and the Norwegian Diabetes Association.

This research was funded in part, by the Wellcome Trust [Grant numbers WT104150 and WT220390]. For the purpose of open access, the authors have applied a CC BY public copyright licence to any Author Accepted Manuscript version arising from this submission.

#### Acknowledgements and ethical approvals by contributing study

**Avon Longitudinal Study of Parents And Children (ALSPAC)**

Core funding for ALSPAC is provided by the UK Medical Research Council and Wellcome (217065/Z/19/Z) and the University of Bristol. Genotyping of the ALSPAC maternal samples was funded by the Wellcome Trust (WT088806), and the offspring samples were genotyped by Sample Logistics and Genotyping Facilities at the Wellcome Trust Sanger Institute and LabCorp (Laboratory Corporation of America) using support from 23andMe. Specific funds for recent detailed data collection on the mothers were obtained from the US National Institutes of Health (R01 DK077659) and Wellcome Trust (WT087997MA) for completion of selected items of obstetric data extraction, including placental weights. A comprehensive list of grants funding is available on the ALSPAC website (http://www.bristol.ac.uk/alspac/external/documents/grant-acknowledgements.pdf). This publication is the work of the authors and R.M. Freathy will serve as guarantor for the contents of this paper.

We are extremely grateful to all the families who took part in this study, the midwives for their help in recruiting them, and the whole ALSPAC team, which includes interviewers, computer and laboratory technicians, clerical workers, research scientists, volunteers, managers, receptionists and nurses.

Ethical approval for the study was obtained from the ALSPAC Ethics and Law Committee and the Local Research Ethics Committees. Informed consent for the use of data collected via questionnaires and clinics was obtained from participants following the recommendations of the ALSPAC Ethics and Law Committee at the time.

**Copenhagen Hospital Biobank Reproductive Health Study (CHB-RHS)**

The Copenhagen Hospital Biobank (CHB) is funded by the Danish Administrative Regions.

We are grateful to the staff at the general hospitals of the capital area as well as the staff in the Biobank Unit of the Copenhagen University Hospital for their assistance in the establishment and operation of the Copenhagen Hospital Biobank.

Approval of the CHB-RHS was obtained from the Danish National Committee on Health Research Ethics (NVK-1805807) and the Capital Region Data Protection Agency (P-2019-49).

**Danish Blood Donor Study (DBDS)**

The Danish Blood Donor Study (DBDS) is funded by The Danish Council for Independent Research - Medical Sciences, The Danish Administrative Regions (Bio- and Genome Bank Denmark).

We thank the Danish blood donors for their valuable participation in the Danish Blood Donor Study, and the staff at the blood centres for making this study possible.

The genetic studies under DBDS (DBDS healthy ageing genetic studies and DBDSII) have been approved by the Danish National Committee on Health Research Ethics (NVK-1700407) and Zealand Regional Committee on Health Research Ethics (Sj-740). Handling of data is approved by the Capital Region Data Protection Agency (P-2019-99)

**Danish National Birth Cohort Preterm Birth Study (DNBC-PTB)**

The DNBC Preterm Birth Study is a nested study within the Danish National Birth Cohort (DNBC). The DNBC was established with a significant grant from the Danish National Research Foundation. Additional support was obtained from the Danish Regional Committees, the Pharmacy Foundation, the Egmont Foundation, the March of Dimes Birth Defects Foundation, the Health Foundation and other minor grants. The DNBC Biobank is a part of the Danish National Biobank resource and has been supported by the Novo Nordisk Foundation and the Lundbeck Foundation. The generation of GWAS genotype data for the DNBC samples was carried out within the Gene Environment Association Studies (GENEVA) Consortium with funding provided through the National Institutes of Health's Genes, Environment, and Health Initiative (U01HG004423; U01HG004446; U01HG004438).

We are very grateful to all DNBC participants. We would also like to thank everyone involved in data collection and biological material handling.

The Danish Scientific Ethics Committee and the Danish Data Protection Agency approved the DNBC.

**Exeter Family Study of Childhood Health (EFSOCH)**

The Exeter Family Study of Childhood Health (EFSOCH) was supported by South West National Health Service (NHS) Research and Development, Exeter NHS Research and Development, the Darlington Trust, the National Institute for Health and Care (NIHR) Research Exeter Biomedical Research Centre and National Institute for Health and Care Research Exeter Clinical Research Facility. The views expressed are those of the authors and not necessarily those of the NIHR or the Department of Health and Social Care. Genotyping of the EFSOCH study samples was funded by the Wellcome Trust and Royal Society grant WT104150.

Ethical approval for the Exeter Family of Childhood Health was given by the North and East Devon (UK) Local Research Ethics Committee (approval number 1104), and informed consent was obtained from the parents of the newborns.

**Genetics of Glucose regulation in Gestation and Growth (Gen3G)**
The Gen3G prospective cohort was supported by the Fonds de Recherche du Québec – Santé (FRQS – subvention Fonctionnement – Recherche Clinique - grant #20697) and by a Canadian Institute of Health Research (CIHR) Operating grant (Institute of Nutrition, Metabolism and Diabetes; MOP-115071). Gen3G investigators would like to acknowledge the participants, the research staff from the endocrinology group at the Centre de Recherche of the Centre Hospitalier Universitaire de Sherbrooke (CRCHUS), the clinical and research staff of the Clinique de Prélèvement en Grossesse du CHUS, and the clinical staff from the CHUS Obstetric Department.

The ethics review committee at Centre Hospitalier Universitaire de Sherbrooke approved the study, and participants gave written informed consent.

**Generation R Study**
The general design of Generation R Study is made possible by financial support from Erasmus MC, University Medical Center Rotterdam, Erasmus University Rotterdam, the Netherlands Organization for Health Research and Development (ZonMw), the Netherlands Organisation for Scientific Research (NWO), the Ministry of Health, Welfare and Sport and the Ministry of Youth and Families. This project received funding from the European Union's Horizon 2020 Research and Innovation Programme under grant agreements 733206 (LifeCycle), 874739 (LongITools), 824989 (EUCAN-Connect), 874583 (ATHLETE).

The Generation R Study is conducted by Erasmus MC, University Medical Center Rotterdam in close collaboration with the School of Law and Faculty of Social Sciences of the Erasmus University Rotterdam, the Municipal Health Service Rotterdam area, Rotterdam, the Rotterdam Homecare Foundation, Rotterdam and the Stichting Trombosedienst & Artsenlaboratorium Rijnmond (STAR-MDC), Rotterdam. We gratefully acknowledge the contribution of children and parents, general practitioners, hospitals, midwives and pharmacies in Rotterdam. The generation and management of GWAS genotype data for the Generation R Study were done at the Genetic Laboratory of the Department of Internal Medicine, Erasmus MC, The Netherlands. We would like to thank Karol Estrada, Dr. Tobias A. Knoch, Anis Abuseiris, Luc V. de Zeeuw, and Rob de Graaf, for their help in creating GRIMP, BigGRID, MediGRID, and Services@MediGRID/D-Grid, (funded by the German Bundesministerium fuer Forschung und Technology; grants 01 AK 803 A-H, 01 IG 07015 G) for access to their grid computing resources. We thank Mila Jhamai, Manoushka Ganesh, Pascal Arp, Marijn Verkerk, Lizbeth Herrera and Marjolein Peters for their help in creating, managing and QC of the GWAS database. Also, we thank Karol Estrada for their support in creation and analysis of imputed data.

The Generation R Study has been approved by the Medical Ethics Committee of Erasmus MC, University Medical Center Rotterdam. Written informed consent was obtained for all participants.

**GoDARTS & goSHARE / Walker Cohort**
Genotype data was obtained from the Genetics of Diabetes Audit and Research Tayside and Scotland (GoDARTS) and goSHARE (genetics of Scottish Health Research Register). Phenotype data was obtained from The Walker Cohort.

The GoDARTS bioresource was funded largely by Wellcome Trust (072960/Z/03/Z, 084726/Z/08/Z, 084727/Z/08/Z, 085475/Z/08/Z, and 085475/B/08/Z) and as part of the European Union IMI-SUMMIT program. SHARE is a NHS Scotland Research (NRS) infrastructure initiative and is funded by the Chief Scientists Office of the Scottish Government. Additional funding and initiation of the spare blood retention in Tayside was supported by The Wellcome Trust Biomedical Resource Award Number 099177/Z/12/Z.

The GoDARTS bioresource, and its links to the long-term EMR, is approved by the NHS Tayside Caldicott Guardians, the local research ethics committee, and the Tayside Tissue Bank, and access to the resource is regulated by the GoDARTS Access Group. The EMR is fully anonymized and provided to researchers through robust information governance protocols administered by the Health Informatics Centre (HIC), which functions as a secure portal between the NHS Tayside and the University of Dundee research environment.  SHARE has NHS research ethics approval to approach patients attending NHS facilities to ask them to become part of SHARE and to use their stored data to identify them as potentially suitable participants in medical research.

**Genetics of Overweight Young Adults – reference group mothers and children (GOYA-REF)**
**Genetics of Overweight Young Adults – mothers with overweight and their children (GOYA-MO-OW)**

GOYA is a nested study within the Danish National Birth Cohort (DNBC), and conducted in collaboration with the MRC Integrative Epidemiology Unit at the University of Bristol (MC_UU_12013/1-9). The genotyping for DNBC-GOYA study was funded by the Wellcome Trust (WT 084762).

The study is approved by all the regional scientific ethics committees in Denmark, by the central scientific ethics committee for whole Denmark and by the Danish Data Protection Board.

**Genetics of Overweight Young Adults – children with overweight and their mothers (GOYA-CH-OW)**

GOYA-offspring study is a nested study within the DNBC-GOYA study. The genotyping of the GOYA-offspring was financed by the Novo Nordisk Foundation Center for Basic Metabolic Research.

The study was approved by the Danish Ethical Committee (1-10-72-195-13 and 1-10-72-261-14). The study was conducted in accordance with the principles of the Declaration of Helsinki.

**Nord-Trøndelag Health Study (The HUNT Study)**

The Trøndelag Health Study (HUNT) is a collaboration between HUNT Research Centre (Faculty of Medicine and Health Sciences, Norwegian University of Science and Technology NTNU), Trøndelag County Council, Central Norway Regional Health Authority, and the Norwegian Institute of Public Health.

Ethical approval was obtained from the Regional Committee for Medical Research Ethics (2018/2488/REK midt). All HUNT participants gave informed written consent.

**INfancia y Medio Ambiente (INMA)**

- **INMA-Omni1:** samples from INMA Sabadell and Valencia processed with the Illumina Omni1 array
- **INMA-GSA:** samples from INMA Gipuzkoa processed with the Illumina GSA array

This study was funded by grants from Instituto de Salud Carlos III (CB06/02/0041, G03/176, FIS PS09/00432, FIS-FEDER 03/1615, 04/1509, 04/1112, 04/1931, 05/1079, 05/1052, 06/1213, 06/0867, 07/0314, and 09/02647, 09/00090, 13/02187 and 18/01142), cofunded by ERDF, “A way to make

Europe", Fundació La Marató de TV3 (090430), Fundación Roger Torné, Generalitat de Catalunya-CIRIT (1999SGR 00241), Conselleria de Sanitat Generalitat Valenciana, Department of Health of the Basque Government (2005111093), Basque Department of Health (Projects 2005111093, 2009111069, 2013111089, 2015111065 and 2018111086), Provincial Government of Gipuzkoa (DFG06/002, DFG08/001 and DFG15/221), and annual agreements with the municipalities of the study area (Zumarraga, Urretxu , Legazpi, Azkoitia y Azpeitia y Beasain). ISGlobal acknowledges support from the Spanish Ministry of Science and Innovation through the "Centro de Excelencia Severo Ochoa 2019-2023" Program (CEX2018-000806-S), and from the Generalitat de Catalunya through the CERCA Program.

Informed consent was obtained from all participants and the study was approved by the Hospital Ethics Committees in each participating region.

**Initiative for Integrative Psychiatric Research (iPSYCH) Cohort**

The iPSYCH study was supported by grants from the Lundbeck Foundation (R102-A9118, R155-2014-1724 and R248-2017-2003), NIMH (1U01MH109514-01) and the Universities and University Hospitals of Aarhus and Copenhagen. The research was conducted using the Danish National Biobank resource, supported by the Novo Nordisk Foundation. Genotyping of iPSYCH samples was further supported by grants from the Stanley Foundation, the Simons Foundation (SFARI 311789), and the NIMH (5U01MH094432-02). High-performance computer capacity for handling and statistical analysis of iPSYCH data on the GenomeDK HPC facility was provided by the Center for Genomics and Personalized Medicine and the Centre for Integrative Sequencing, iSEQ, Aarhus University, Denmark.

The Danish Scientific Ethics Committee, the Danish Data Protection Agency and the Danish Neonatal Screening Biobank Steering Committee approved the iPSYCH study.

**The Norwegian Mother, Father and Child Cohort Study (MoBa)**
The Norwegian Mother, Father and Child Cohort Study (MoBa) is a population-based pregnancy cohort study conducted by the Norwegian Institute of Public Health. Participants were recruited from all over Norway from 1999-2008. The women consented to participation in 41% of the pregnancies. The cohort includes approximately 114.500 children, 95.200 mothers and 75.200 fathers. The current study is based on version 10 of the quality-assured data files released for research in *Novel Tools for Early Childhood Predisposition to Obesity*. The establishment of MoBa and initial data collection was based on a license from the Norwegian Data Protection Agency and approval from The Regional Committees for Medical and Health Research Ethics. The MoBa cohort is currently regulated by the Norwegian Health Registry Act. The current study was approved by The Regional Committees for Medical and Health Research Ethics (no. 2012/67). The Medical Birth Registry (MBRN) is a national health registry containing information about all births in Norway.

We thank the Norwegian Institute of Public Health (NIPH) for generating high-quality genomic data. This research is part of the HARVEST collaboration, supported by the Research Council of Norway (#229624). We also thank the NORMENT Centre for providing genotype data, funded by the Research Council of Norway (#223273), South East Norway Health Authorities and Stiftelsen Kristian Gerhard Jebsen. We further thank the Center for Diabetes Research, the University of Bergen for providing genotype data and performing quality control and imputation of the data funded by the

ERC AdG project SELECTionPREDISPOSED, Stiftelsen Kristian Gerhard Jebsen, Trond Mohn Foundation, the Research Council of Norway, the Novo Nordisk Foundation, the University of Bergen, and the Western Norway Health Authorities.

Supported by grants (to P.R.N.) from the European Research Council (AdG #293574), the Bergen Research Foundation ("Utilizing the Mother and Child Cohort and the Medical Birth Registry for Better Health"), Stiftelsen Kristian Gerhard Jebsen (Translational Medical Center), the University of Bergen, the Research Council of Norway (FRIPRO grant #240413), the Western Norway Regional Health Authority (Strategic Fund "Personalized Medicine for Children and Adults"), the Novo Nordisk Foundation (grant #54741), and the Norwegian Diabetes Association; and (to S.J.) Helse Vest's Open Research Grant (grant #912250). This work was partly supported by the Research Council of Norway through its Centres of Excellence funding scheme (#262700), Better Health by Harvesting Biobanks (#229624) and The Swedish Research Council, Stockholm, Sweden (2015-02559), The Research Council of Norway, Oslo, Norway (FRIMEDBIO #547711, March of Dimes (#21-FY16-121). The Norwegian Mother, Father, and Child Cohort Study is supported by the Norwegian Ministry of Health and Care Services and the Ministry of Education and Research, NIH/NIEHS (contract no N01-ES-75558), NIH/NINDS (grant no.1 UO1 NS 047537-01 and grant no.2 UO1 NS 047537-06A1).

Data from the Norwegian Mother, Father and Child Cohort Study and the Medical Birth Registry of Norway used in this study are managed by the national health register holders in Norway (Norwegian Institute of public health) and can be made available to researchers, provided approval from the Regional Committees for Medical and Health Research Ethics (REC), compliance with the EU General Data Protection Regulation (GDPR) and approval from the data owners. The consent given by the participants does not open for storage of data on an individual level in repositories or journals. Researchers who want access to data sets for replication should apply through helsedata.no. Access to data sets requires approval from The Regional Committee for Medical and Health Research Ethics in Norway and an agreement with MoBa.

Analyses were performed using digital laboratories in HUNT Cloud at the Norwegian University of Science and Technology, Trondheim, Norway. We are grateful for outstanding support from the HUNT Cloud community.

The participating families who contributed with data and biological material are gratefully acknowledged.

**Northern Finland Birth Cohort 1966 (NFBC1966)**

University of Oulu, Northern Finland Birth Cohort 1966. University of Oulu. http://urn.fi/urn:nbn:fi:att:bc1e5408-980e-4a62-b899-43bec3755243

NFBC1966 31yr follow-up received financial support from University of Oulu Grant no. 65354, Oulu University Hospital Grant no. 2/97, 8/97, Ministry of Health and Social Affairs Grant no. 23/251/97, 160/97, 190/97, National Institute for Health and Welfare, Helsinki Grant no. 54121, Regional Institute of Occupational Health, Oulu, Finland Grant no. 50621, 54231. NFBC1966 46yr follow-up received financial support from University of Oulu Grant no. 24000692, Oulu University Hospital Grant no. 24301140, ERDF European Regional Development Fund Grant no. 539/2010 A31592.

We thank all cohort members and researchers who participated in the study. We also wish acknowledge the work of the NFBC project center.

Approval for the studies was granted by the Northern Ostrobothnia Hospital District Ethical Committee 94/2011 (12.12.2011), Finland in accordance with the declaration of Helsinki.

**Northern Finland Birth Cohort 1986 (NFBC1986)**

University of Oulu, Northern Finland Birth Cohort 1986. University of Oulu.
http://urn.fi/urn:nbn:fi:att:f5c10eef-3d25-4bd0-beb8-f2d59df95b8e

Financial support was received from the following grants: EU QLG1-CT-2000-01643 (EUROBLCS) Grant no. E51560, NorFA Grant no. 731, 20056, 30167, USA / NIH 2000 G DF682 Grant no. 50945.

We thank all cohort members and researchers who have participated in the study. We also wish acknowledge the work of the NFBC project center.

Approval for the studies was granted by the Northern Ostrobothnia Hospital District Ethical Committee 108/2017 (15.1.2018), Finland in accordance with the declaration of Helsinki.

**Physical Activity and Nutrition in Children study (PANIC)**

Funding was received from Ministry of Education and Culture of Finland, Ministry of Social Affairs and Health of Finland, Social Insurance Institution of Finland, State Research Funding, Kuopio University Hospital Catchment Area, Finnish Cultural Foundation, Finnish Government, Finnish Innovation Fund Sitra, Juho Vainio Foundation, Finnish Paediatric Research Foundation, Finnish Diabetes Research Foundation, Yrjö Jahnsson Foundation, Finnish Foundation for Cardiovascular Research, Paavo Nurmi Foundation.

We thank all participants and researchers of the PANIC study for their contribution.

The Research Ethics Committee of the Hospital District of Northern Savo approved the study protocol in 2006 (Statement 69/2006). The parents or caregivers of the children gave their written informed consent, and the children provided their assent to participation. The PANIC study has been carried out in accordance with the principles of the Declaration of Helsinki.

**The Raine Study**

The authors are grateful to the Raine Study participants and their families, and to the Raine Study research staff for cohort coordination and data collection. The authors gratefully acknowledge the NHMRC for their long term contribution to funding the study over the last 30 years and also the following Institutions for providing funding for Core Management of the Raine Study: The University of Western Australia (UWA), Raine Medical Research Foundation, UWA Faculty of Medicine, Dentistry and Health Sciences, The Telethon Kids Institute, Women and Infants Research Foundation, Curtin University, Edith Cowan University, Murdoch University, and the University of Notre Dame. The authors gratefully acknowledge the assistance of the Western Australian DNA Bank (National Health and Medical Research Council of Australia National Enabling Facility). This work was supported by resources provided by the Pawsey Supercomputing Centre with funding from the Australian Government and the Government of Western Australia.

The Raine Study was supported by the Raine Medical Research Foundation, National Health and Medical Research Council of Australia (NHMRC grant numbers 572613, 403981 and 003209) and the Canadian Institutes of Health Research (CIHR grant number MOP-82893).

**Roskilde**
This work was supported by research grants from the Danish Diabetes Academy supported by the Novo Nordisk Foundation, Shipowner Per Henriksen and Wife's Foundation, The Research Foundation of Region Zealand, Dagmar Marshall's Foundation and Ivan Nielsen's Foundation.

The study was approved by the Ethics Committee for Region Zealand (SJ-55 and SJ-347) and the Danish Data Protection Agency and was registered in ClinicalTrials.gov (Identifier: NCT00836524). The study was conducted in accordance with the principles of the Helsinki Declaration. All study participants provided written informed consent.

**Statens Serum Institut Genetic Epidemiology (SSI-GE) cohorts.**
Note: SSI-GE is comprised of samples with GWAS data from studies conducted at Statens Serum Institut of febrile seizures (SSI-GE FS), infantile hypertrophic pyloric stenosis (SSI-GE IHPS casesCtrls and SSI-GE IHPS CIDR), pre-eclampsia (SSI-GE PE ctrls), opioid dependence (SSI-GE OPI), postpartum depression (SSI-GE PPD), and infectious mononucleosis (SSI-GE Mono ctrls).

Samples for these projects were drawn from the Danish National Biobank, which is supported by the Novo Nordisk Foundation with additional partial support from the Lundbeck Foundation and the Danish Medical Research Council.

The SSI-GE studies were approved by the Regional Scientific Ethics Committees of Copenhagen, the Danish Data Protection Agency and the Danish Neonatal Screening Biobank Steering Committee.

**23andMe, Inc. published GWAS summary statistics**

We thank the employees and research participants of 23andMe, Inc. for providing the full sets of summary statistics from the published GWAS of gestational duration (Zhang et al. 2017 NEJM) and of nausea and vomiting in pregnancy and hyperemesis gravidarum (Fejzo et al. 2018 *Nat Comms*). The full GWAS summary statistics for the 23andMe discovery data set is available through 23andMe to qualified researchers under an agreement with 23andMe that protects the privacy of the 23andMe participants. Please visit https://research.23andme.com/collaborate/#dataset-access/ for more information and to apply to access the data.

**Membership of the Early Growth Genetics (EGG) Consortium**

Full list of members (as of October 2022), listed in alphabetical order.

Linda S Adair[1], Emma Ahlqvist[2], Tarunveer S Ahluwalia[3,4,5], Sonia Anand[6,7,8], Izzuddin Aris[9], Mustafa Atalay[10], Jonas Bacelis[11], Robin N Beaumont[12], Jeffrey J Beck[13,14], Jose Ramon Bilbao[15,16,17], Thomas Bond[18,19,20,21], Klaus Bønnelykke[3], Dorret I Boomsma[22,23,24], Jonathan P Bradfield[25,26], Mariona Bustamante[27,28,29], Yee-Ming Chan[30,31,32], Lachlan Coin[33], Cyrus Cooper[34], William E Copeland[35], Diana L Cousminer[36,37,38], Alexessander Couto-Alves[39], John A Curtin[40], Adnan Custovic[41], Felix R Day[42], George Dedoussis[43], Maneka De Silva[44], Paul Elliott[18], Aino-Maija Eloranta[45], Johan Eriksson[46,47,48], David Evans[49,50,51], João Fadista[52], Bjarke Feenstra[53], Janine Felix[54,55], Joshua Fisher[56], Christopher Flatley[11], Tim Frayling[57], Rachel M Freathy[58], Romy Gaillard[59], Frank Geller[53], Joseph T Glessner[60,61], Struan F Grant[62,63,64,65,66], Niels a Grarup[67], Leif Groop[68,69], Monica Guxens[70,71,72,73], Dexter Hadley[74], Hakon Hakonarson[75], Torben Hansen[76], Andrew T Hattersley[77], M Geoffrey Hayes[78,79,80], Johannes Hebebrand[81,82], Sami Heikkinen[10], Joachim Heinrich[83], Anni Heiskala[84], Øyvind Helgeland[85,86,87], Tine Brink Henrisksen[88], Anke Hinney[81,82], Joel N Hirschhorn[89], Marie-France Hivert[9,90,91], Berthold Hocher[92,93,94], John W Holloway[95], Jens-Christian Holm[96], Momoko Horikoshi[97], Jouke-Jan Hottenga[98,99,100], Alice E Hughes[101], Elina Hyppönen[102,103], Bo Jacobsson[104,105], Vincent Jaddoe[54,55], Marjo-Riitta Järvelin[106,107,108,109,110], Stefan Johansson[86,111], Marika A Kaakinen[112,113,114], Heidi J Kalkwarf[115], Irfahan Kassam[116], Antje Körner[117,118,119], Sailesh Kotecha[120], Zoltán Kutalik[121,122,123], Timo A Lakka[124], Amel Lamri[125,7], Joan M Lappe[126], Deborah A Lawlor[51,127], Terho Lehtimäki[128,129,130], Alexandra M Lewin[131], Brandon Lim[12], Cecilia M Lindgren[132,133], Virpi Lindi[10], Allan Linneberg[134], Jun Liu[135], Bill Lowe[136], Leo-Pekka Lyytikäinen[128,129,130], Ronald CW Ma[137,138,139], Reedik Mägi[140], Per Magnus[141], Mark I McCarthy[142], Nina S McCarthy[143], Mads Melbye[144,145,146,147], Gunn-Helen Moen[148,149,150,151], Karen L Mohlke[152], Dennis Mook-Kanamori[153,154], Camilla S Morgen[155], Andrew P Morris[156], Ehsan Motazedi[157], Louis J Muglia[158,159,160,161], Jeff Murray[162], Ronny Myhre[163], Juha Mykkänen[164,165], Harri Niinikoski[164,165,166], Pal Rasmus Njølstad[167,168], Ellen Aagaard Nohr[169], Ioanna Ntalla[170], Sharon E Oberfield[171], Emily Oken[9], Ken Ong[172,173], Katja Pahkala[164,165,174], Kalliope Panoutsopoulou[175], Oluf Pedersen[176], Craig E Pennell[177,178], John Perry[179], Triinu Peters[81], Roseann E Peterson[180], Christine Power[181], Rashmi B Prasad[182,183], Inga Prokopenko[184,185,186], Olli T Raitakari[164,165,187], Rebecca M Reynolds[188], Rebecca C Richmond[189,24], Alina Rodriguez[190,191], Justiina Ronkainen[84], Rany Salem[192,193,194,195], Seang Mei Saw[196,197], William Schierding[198], Theresia M Schnurr[67], Sylvain Sebert[199], John A Shepherd[200], Angela Simpson[40], Line Skotte[52], Pol Sole-Navais[104], Thorkild IA Sørensen[201], Marie Standl[202], Eric AP Steegers[203], Sara Elizabeth Stinson[76], Beate St Pourcain[204,189], David Strachan[205], Jordi Sunyer[27,28,29,206], Yik Ying Teo[207,208], Elisabeth Thiering[202,209], Nic J Timpson[21], Jessica Tyrrell[12,210], Andre G Uitterlinden[211], Cornelia van Dujin[212,213], Marc Vaudel[214,105], Martine Vrijheid[27,28,29], Tanja Vrijkotte[157], Carol A Wang[177,178], Ni Wang[215], Nicole M Warrington[148,216,150], William J Watkins[120], H-Erich Wichmann[217,218,219], Elisabeth Widén[220], Gonneke Willemsen[221], James F Wilson[222], Xiaoping Wu[53], Hanieh

Yaghootkar[223,224,225], Eleftheria Zeggini[226,227], Babette S Zemel[228], Ge Zhang[229,159,160], Jia Zhu[230]

[1]Department of Nutrition, University of North Carolina, Chapel Hill, NC, United States, [2]Department of Clinical Sciences, Diabetes and Endocrinology, Lund University Diabetes Centre, Malmö, Sweden, [3]COPSAC, Copenhagen Prospective Studies on Asthma in Childhood, Herlev and Gentofte Hospital, University of Copenhagen, Copenhagen, Denmark, [4]Steno Diabetes Center Copenhagen, Herlev, Denmark, [5]The Bioinformatics Center, Department of Biology, University of Copenhagen, Copenhagen, Denmark, [6]Department of Medicine, McMaster University, Hamilton, Ontario, Canada., [7]Population Health Research Institute, Hamilton Health Sciences and McMaster University, Hamilton, Ontario, Canada, [8]Department of Health Research Methods, Evidence, and Impact, McMaster University, Hamilton, Ontario, Canada., [9]Department of Population Medicine, Harvard Pilgrim Health Care Institute, Harvard Medical School, Boston, USA, [10]Institute of Biomedicine, School of Medicine, University of Eastern Finland, Kuopio Campus, Finland, [11]Department of Obstetrics and Gynecology, Sahlgrenska University Hospital, Gothenburg, Sweden, [12]Institute of Biomedical and Clinical Science, University of Exeter Medical School, University of Exeter, Royal Devon and Exeter Hospital, Exeter, UK, [13]Avera Institute for Human Genetics, Avera McKennan Hospital and University Health Center, Sioux Falls, South Dakota, USA, [14]Department of Psychiatry, University of South Dakota Sanford School of Medicine, South Dakota, USA, [15]Faculty of Medicine and Nursing, Department of Genetics, Physical Anthropology and Animal Physiology, University of the Basque Country (UPV/EHU), Leioa, Bizkaia, Spain, [16]Biocruces Bizkaia Health Research Institute, Barakaldo, Bizkaia, Spain, [17]CIBER de Diabetes y Enfermedades Metabólicas Asociadas (CIBEDEM), Madrid, Spain, [18]Department of Epidemiology and Biostatistics, School of Public Health, Imperial College London, London, UK, [19]MRC-PHE Centre for Environment and Health, School of Public Health, Imperial College London, London, UK., [20]The University of Queensland Diamantina Institute, The University of Queensland, Brisbane, Australia, [21]MRC Integrative Epidemiology Unit, University of Bristol, Bristol, UK, [22]Netherlands Twin Register, Department of Biological Psychology, Vrije Universiteit, Amsterdam, the Netherlands, [23]Amsterdam Reproduction & Development (AR&D) research institute, the Netherlands, [24]Population Health Sciences, Bristol Medical School, University of Bristol, Bristol, UK, [25]Center for Applied Genomics, The Children's Hospital of Philadelphia, Philadelphia, USA, [26]Quantinuum Research LLC, San Diego, USA, [27]ISGlobal, Institute for Global Health, Barcelona, Spain, [28]CIBER de Epidemiología y Salud Pública (CIBERESP), Madrid, Spain, [29]Universitat Pompeu Fabra (UPF), Barcelona, Spain, [30]1 Division of Endocrinology, Boston Children's Hospital, Boston, MA 02115, USA, [31]Program in Medical and Population Genetics, The Broad Institute of MIT and Harvard, Cambridge, MA 02142, USA, [32]Department of Pediatrics, Harvard Medical School, Boston, MA 02115, USA, [33]Institute for Molecular Bioscience, University of Queensland, Brisbane, Australia, [34]MRC Lifecourse Epidemiology Unit, Faculty of Medicine, University of Southampton, Southampton, UK, [35]Department of Psychitry, University of Vermont,

Burlington, Vermont, [36]Division of Human Genetics, Children's Hospital of Philadelphia, Philadelphia, PA, USA., [37]Department of Genetics, University of Pennsylvania, Philadelphia, PA, USA, [38]Center for Spatial and Functional Genomics, Children's Hospital of Philadelphia, Philadelphia, PA, USA, [39]School of biosciences and medicine, University of Surrey, United Kingdom, [40]Division of Immunology, Immunity to Infection and Respiratory Medicine, School of Biological Sciences, The University of Manchester, Manchester Academic Health Science Centre, and Manchester University NHS Foundation Trust, Manchester, UK, [41]National Heart and Lund Institute, Imperial College London, UK, [42]MRC Epidemiology Unit, University of Cambridge School of Clinical Medicine, Cambridge, UK, [43]Department of Nutrition and Dietetics, Harokopio University of Athens, Athens, Greece., [44]Center for Life Course Health Research, Faculty of Medicine, University of Oulu, Oulu, Finland, [45]Institute of Public Health and Clinical Nutrition, School of Medicine, University of Eastern Finland, Kuopio Campus, Finland, [46]Department of General Practice and Primary Health Care, University of Helsinki, Folkhälsan Research Center, Helsinki, Finland, [47]National University Singapore, Yong Loo Lin School of Medicine, Human Potential Translational Research Programme and Department of Obstetrics and Gynecology, Singapore, Singapore, [48]Singapore Institute for Clinical Sciences (SICS), Agency for Science, Technology and Research (A*STAR), Singapore, [49]Institute for Molecular Bioscience, The University of Queensland, Brisbane, Australia, [50]University of Queensland Diamantina Institute, University of Queensland, Brisbane, Australia, [51]MRC integrative Epidemiology Unit at the University of Bristol, UK, [52]Department of Epidemiology Research, Statens Serum Institut, Copenhagen, Denmark, [53]Department of Epidemiology Research, Statens Serum Institute, Copenhagen, Denmark, [54]The Generation R Study Group, Erasmus MC, University Medical Center Rotterdam, Rotterdam, the Netherlands, [55]Department of Pediatrics, Erasmus MC, University Medical Center Rotterdam, Rotterdam, the Netherlands, [56]School of Medical Science, Menzies Health Institute Queensland, Griffith University Gold Coast Campus, Southport, QLD, Australia, [57]Genetics of Complex Traits, College of Medicine and Health, University of Exeter, Exeter, UK,, [58]Institute of Biomedical and Clinical Science, College of Medicine and Health, University of Exeter, Barrack Road, Exeter, Devon EX2 5DW, UK, [59]Department of Epidemiology, Erasmus MC, University Medical Center Rotterdam, Rotterdam, the Netherlands, [60]Department of Pediatrics, Children's Hospital of Philadelphia, 3401 Civic Center Blvd, Philadelphia, PA 19104, USA., [61]Department of Pediatrics, Division of Human Genetics, Perelman School of Medicine, 3400 Civic Center Blvd, Philadelphia, PA 19104, USA., [62]Division of Human Genetics, The Children's Hospital of Philadelphia, Philadelphia, USA, [63]Division of Endocrinology and Diabetes, The Children's Hospital of Philadelphia, Philadelphia, USA, [64]Department of Pediatrics, University of Pennsylvania Perelman School of Medicine, Philadelphia, United States, [65]Department of Genetics, University of Pennsylvania Perelman School of Medicine, Philadelphia, United States, [66]Center for Applied Genomics, Children's Hospital of Philadelphia, Philadelphia, United States, [67]Novo Nordisk Foundation Center for Basic Metabolic Research, Section of Metabolic Genetics, Faculty of Health and Medical Sciences, University of Copenhagen, Copenhagen, Denmark, [68]Institute for Molecular Medicine, Finland (FIMM), University of

Helsinki, Helsinki, Finland, [69]Department of Clinical Sciences, Diabetes and Endocrinology, Lund University Diabetes Centre, Malm√∂, Sweden, [70]ISGlobal, Barcelona, Spain, [71]CIBER de Epidemiología y Salud Pública (CIBERESP), Instituto de Salud Carlos III, Madrid, Spain, [72]Universitat Pompeu Fabra, Barcelona, Spain, [73]Department of Child and Adolescent Psychiatry/Psychology, Erasmus MC, University Medical Centre, Rotterdam, The Netherlands, [74]Department of Pediatrics, University of California San Francisco School of Medicine, San Francisco, USA, [75]Children's Hospital of Philadelphia, Leonard Madlyn Abramson Research Center, Philadelphia, PA, USA, [76]Novo Nordisk Foundation Center for Basic Metabolic Research, Faculty of Health and Medical Sciences, University of Copenhagen, Denmark, [77]University of Exeter Medical School, College of Medicine and Health, University of Exeter, Exeter, UK, [78]Division of Endocrinology, Metabolism, and Molecular Medicine, Department of Medicine, Northwestern University Feinberg School of Medicine, Chicago, IL USA, [79]Center for Genetic Medicine, Northwestern University Feinberg School of Medicine, Chicago, IL USA, [80]Department of Anthropology, Northwestern University, Evanston, IL USA, [81]Department of Child and Adolescent Psychiatry, Psychosomatics and Psychotherapy, University Hospital Essen, University of Duisburg-Essen, Essen, Germany, [82]Center for Translational Neuro- and Behavioural Sciences, University Hospital Essen, Essen, Germany, [83]Institute and Clinic for Occupational, Social and Environmental Medicine, University Hospital, LMU Munich, Munich, Germany, [84]Center for Life Course Health Research, University of Oulu, Oulu, Finland, [85]Department of Genetics and Bioinformatics, Domain of Health Data and Digitalisation, Norwegian Institute of Public Health, Oslo, Norway, [86]KG Jebsen Center for Diabetes Research, Department of Clinical Science, University of Bergen, Bergen, Norway, [87]Department of Pediatrics, Haukeland University Hospital, Bergen, Norway, [88]Department of Clinical Medicine - Department of Paediatrics, Aarhus University Hospital, Aarhus N, Denmark, [89]Broad Institute of MIT and Harvard, Cambridge, USA, [90]Diabetes Center, Massachusetts General Hospital, Boston, USA, [91]Department of Medicine, Universite de Sherbrooke, Sherbooke, Canada, [92]Institute of Nutritional Science, University of Potsdam, Nuthetal, Germany, [93]The First Affiliated Hospital of Jinan University, Guangzhou, China, [94]Department of Medicine Nephrology, Medical Faculty, Mannheim Heidelberg University, 68167 Mannheim, Germany, [95]Human Development & Health, Faculty of Medicine, University of Southampton, Southampton, UK, [96]The Children's Obesity Clinic, accredited European Centre for Obesity Management, Department of Pediatrics, Holbæk Hospital, Denmark, [97]RIKEN, Centre for Integrative Medical Sciences, Laboratory for Genomics of Diabetes and Metabolism, Yokohama, Japan, [98]Netherlands Twin Register, Department of Biological Psychology, Vrije Universiteit Amsterdam, Amsterdam, the Netherlands, [99]Netherlands Twin Register, Department of Biological Psychology, VU University, Amsterdam, the Netherlands, [100]Amsterdam Public Health, Amsterdam, the Netherlands, [101]Institute of Biomedical and Clinical Science, University of Exeter Medical School, Exeter, UK, [102]Australian Centre for Precision Health, Unit of Clinical and Health Sciences, University of South Australia, Adelaide, Australia, [103]South Australian Health and Medical Research Institute, Adelaide, Australia,

[104]Department of Obstetrics and Gynaecology, Sahlgrenska Academy, Institute of Clinical Science, University of Gothenburg, Gothenburg, Sweden, [105]Department of Genetics and Bioinformatics, Health Data and Digitalization, Norwegian Institute of Public Health, Oslo, Norway, [106]Institute of Health Sciences, University of Oulu, Oulu, Finland, [107]Biocenter Oulu, University of Oulu, Oulu, Finland, [108]Department of Epidemiology and Biostatistics, MRC Health Protection Agency (HPE) Centre for Environment and Health, School of Public Health, Imperial College London, UK, [109]Department of Children and Young People and Families, National Institute for Health and Welfare, Oulu, Finland, [110]1Unit of Primary Care, Oulu University Hospital, Oulu, Finland, [111]Center for Medical Genetics and molecular Medicine, Haukeland University Hospital, Bergen, Norway, [112]Department of Clinical and Experimental Medicine, University of Surrey, Guildford, UK, [113]People-Centred Artificial Intelligence Institute, University of Surrey, Guildford, UK, [114]Department of Medicine, Imperial College London, London, UK, [115]Division of Gastroenterology, Hepatology and Nutrition, Cincinnati Children's Hospital Medical Center, Cincinnati, USA, [116]Lee Kong Chian School of Medicine, Nanyang Technological University, Singapore, Republic of Singapore, [117]University of Leipzig, Medical Faculty, Dept. of Women and Child Health, Pediatric Research Center, Leipzig, Germany, [118]Helmholtz Institute for Metabolic, Obesity and Vascular Research (HI-MAG) of the Helmholtz Zentrum München at the University of Leipzig and University Hospital Leipzig, Leipzig, Germany, [119]LIFE Child, University of Leipzig, Medical Faculty, LIFE–Leipzig Research Center for Civilization Diseases, Leipzig, Germany,, [120]Department of Child Health, School of Medicine, Cardiff Univeristy, Cardiff, UK, [121]Institute of Primary Care and Public Health, University of Lausanne, Lausanne, Switzerland, [122]Department of Computational Biology, University of Lausanne, Lausanne, Switzerland, [123]Swiss Institute of Bioinformatics, Lausanne, Switzerland, [124]Institute of Biomedicine, School of Medicine, University of Eastern Finland, Kuopio Campus, Finland; Department of Clinical Physiology and Nuclear Medicine, Kuopio University Hospital, Kuopio, Finland; Foundation for Research in Health Exercise and Nutrition, Kuopio Research Institute of Exercise Medicine, Kuopio, Finland, [125]Department of Medicine, McMaster University, Hamilton, Ontario, Canada, [126]Division of Endocrinology, Department of Medicine, Creighton University, Omaha, USA, [127]Population Health Science, Bristol Medical School, Bristol University, UK, [128]Department of Clinical Chemistry, Fimlab Laboratories, Tampere 33520, Finland, [129]Department of Clinical Chemistry, Finnish Cardiovascular Research Center - Tampere, Faculty of Medicine and Health Technology, Tampere University, Tampere 33014, Finland, [130]Department of Cardiology, Heart Center, Tampere University Hospital, Tampere 33521, Finland, [131]Department of Medical Statistics, London School of Hygiene and Tropical Medicine, London, United Kingdom, [132]Wellcome Trust Centre for Human Genetics, University of Oxford, Oxford, UK, [133]Li Ka Shing Centre for Health Information and Discovery, The Big Data Institute, University of Oxford, Oxford, UK, [134]Center for Clinical Research and Prevention, Bispebjerg and Frederiksberg Hospital, Copenhagen, Denmark and Department of Clinical Medicine, Faculty of Health and Medical Sciences,University of Copenhagen, Copenhagen, Denmark, [135]Nuffield Department of Population Health, Univeristy of Oxford, Oxford, UK, [136]Department of Medicine,

Northwestern University Feinberg School of Medicine, Rubloff 12, 420 E. Superior St, Chicago, IL 60611, USA, [137]Department of Medicine and Therapeutics, The Chinese University of Hong Kong, [138]Li Ka Shing Institute of Health Sciences, The Chinese University of Hong Kong, Hong Kong, China, [139]Hong Kong Institute of Diabetes and Obesity, The Chinese University of Hong Kong, Hong Kong, China, [140]Estonian Genome Center, University of Tartu, Tartu, Estonia, [141]Norwegian Institute of Public Health, Oslo, Norway, [142]Wellcome Centre for Human Genetics, Univ of Oxford, Oxford, UK (for papers submitted before end 2022); after that, please contact me. Also pls include (Current address: Genentech, 1 DNA Way, South San Francisco, CA 94080) as a footnote, [143]Centre for Genetic Origins of Health and Disease (GOHaD), The University of Western Australia, Crawley, Australia, [144]Department of Clinical Medicine, Universityof Copenhagen, Copenhagen, Denmark, [145]Department of Medicine, Stanford UniversitySchool of Medicine, Stanford, California, USA, [146]Centre for Fertility and Health, NorwegianInstitute of Public Health, Oslo, Norway, [147]K.G. Jebsen Center for Genetic Epidemiology,Norwegian University of Science andTechnology, Trondheim, Norway, [148]Institute of Molecular Biosciences, The University of Queensland, Brisbane, Australia, [149]Institute of Clinical Medicine, Faculty of Medicine, University of Oslo, Norway., [150]K.G. Jebsen Center for Genetic Epidemiology, Department of Public Health and Nursing, NTNU, Norwegian University of Science and Technology, Norway, [151]Population Health Science, Bristol Medical School, University of Bristol, UK, [152]Department of Genetics, University of North Carolina, Chapel Hill, NC, United States, [153]Department of Clinical Epidemiology, Leiden University Medical Center, 2333 ZA Leiden, The Netherlands, [154]Department of Public Health and Primary Care, Leiden University Medical Center, 2333 ZA Leiden, The Netherlands, [155]Department of Public Health, Section of Epidemiology, Faculty of Health and Medical Sciences, University of Copenhagen, Copenhagen, Denmark, [156]Centre for Genetics and Genomics Verusus Arthritis, Centre for Musculoskeletal Research, The University of Manchester, Manchester, UK, [157]Department of Public and Occupational Health, Amsterdam University Medial Center, University of Amsterdam, Amsterdam, [158]Burroughs Wellcome Fund, Research Triangle Park and Department of Pediatrics, Cincinnati Children's Hospital Medical Center and University of Cincinnati College of Medicine, Cincinnati, OH, USA, [159]March of Dimes Prematurity Research Center Ohio Collaborative, Cincinnati, USA, [160]Human Genetics Division, Cincinnati Children's Hospital Medical Center, Cincinnati, USA, [161]President, Burroughs-Wellcome Fund, [162]Dept of Pediatrics, University of Iowa, Iowa City, IA 52240 USA, [163]Department of Genes and Environment, Division of Epidemiology, Norwegian Institute of Public Health, Oslo, Norway, [164]Centre for Population Health Research, University of Turku and Turku University Hospital, Turku, Finland, [165]Research Centre of Applied and Preventive Cardiovascular Medicine, University of Turku, Turku, Finland, [166]Department of Pediatrics and Adolescent Medicine, University of Turku and University Hospital of Turku, Turku, Finland, [167]Mohn Center for Diabetes Precision Medicine, Department of Clinical Science, University of Bergen, NO-5020 Bergen, Norway, [168]Children and Youth Clinic, Haukeland University Hospital, NO-5021 Bergen, Norway, [169]Department of Clinical Research, Research

Unit for Obstetrics and Gynecology, University of Southern Denmark, Odense, Denmark, [170]William Harvey Research Institute, Barts and the London School of Medicine and Dentistry, Queen Mary University of London, London, UK, [171]Division of Pediatric Endocrinology, Diabetes, and Metabolism, Department of Pediatrics, Columbia University Medical Center, New York, USA, [172]MRC Epidemiology Unit, Institute of Metabolic Science, University of Cambridge School of Clinical Medicine, Cambridge, UK, [173]Department of Paediatrics, University of Cambridge School of Clinical Medicine, Cambridge, UK, [174]Paavo Nurmi Centre and Unit for Health and Physical Activity, University of Turku, Turku, Finland, [175]Wellcome Trust Sanger Institute, Wellcome Genome Campus, Hinxton, Cambridgeshire, United Kingdom, [176]Novo Nordisk Foundation Center for Basic Metabolic Research, Faculty of Health and Medical Sciences, University of Copenhagen, 2200 Copenhagen, Denmark, [177]School of Medicine and Public Health, University of Newcastle, Newcastle, NSW, Australia, [178]Hunter Medical Research Institute, Newcastle, NSW, Australia, [179]MRC Epidemiology Unit, Institute of Metabolic Science, University of Cambridge School of Clinical Medicine, Cambridge Biomedical Campus, CB2 0QQ Cambridge, UK, [180]Department of Psychiatry and Behavioral Sciences, College of Medicine, State University of New York Downstate Health Sciences University, Brooklyn, New York, USA, [181]Population, Policy, Practice. Great Ormond Street Institute of Child Health, University College London, London, UK, [182]Department of Clinical Sciences, Diabetes and Endocrinology, Lund University Diabetes Centre, Malmo, Sweden, [183]Institute of Molecular Medicine, Helsinki, Finland, [184]Department of Clinical & Experimental Medicine, University of Surrey, Guildford, UK, [185]UMR 8199 - EGID, Institut Pasteur de Lille, CNRS, University of Lille, F-59000 Lille, France., [186]Institute of Biochemistry and Genetics, Ufa Federal Research Centre, Russian Academy of Sciences, Ufa, Russian Federation, [187]Department of Clinical Physiology and Nuclear Medicine, Turku University Hospital, Turku, Finland, [188]Centre for Cardiovascular Science, Queen's Medical Research Institute, University of Edinburgh, Edinburgh, UK, [189]Medical Research Council Integrative Epidemiology Unit at the University of Bristol, Bristol, UK, [190]Department of Epidemiology and Biostatistics, MRC‚ÄìPHE Centre for Environment & Health, School of Public Health, Imperial College London, London, UK, [191]Department of Psychology, Mid Sweden University, Östersund, Sweden, [192]Herbert Wertheim School of Public Health, University of California San Diego, La Jolla, CA USA, [193]Department of Genetics, Harvard Medical School, Boston, USA, [194]Center for Basic and Translational Obesity Research, Boston Children's Hospital, Boston, USA, [195]Program in Medical and Population Genetics, Broad Institute of Harvard and MIT, Cambridge, USA, [196]Singapore Eye Research Institute, Singapore, [197]Yong Loo Lin School of Medicine, National University of Singapore, Singapore, [198]Liggins Institute, University of Auckland, Auckland, NZ, [199]Research Unit of Population Health, Faculty of Medicine, University of Oulu, Oulu, Finland, [200]Department of Radiology, University of California San Francisco, San Francisco, USA, [201]Department of Public Health and Novo Nordisk Foundation Center for Basic Metabolic Research, Faculty of Health and Medical Sciences, University of Copenhagen, Denmark, [202]Institute of Epidemiology, Helmholtz Zentrum München - German Research Center for

Environmental Health, Neuherberg, Germany, [203]Department of Obstetrics and Gynecology, Erasmus MC, University Medical Center, Rotterdam, the Netherlands, [204]Max Planck Institute for Psycholinguistics, Nijmegen, the Netherlands, [205]Population Health Research Institute, St George's, University of London, London, UK, [206]IMIM (Hospital del Mar Medical Research Institute), Barcelona, Spain, [207]Saw Swee Hock School of Public Health, National University of Singapore and National University Health System, Singapore, Singapore, [208]National University of Singapore, Singapore, Singapore, [209]Division of Metabolic and Nutritional Medicine, Dr. von Hauner Children's Hospital, University of Munich Medical Center, Munich, Germany, [210]European Centre for Environment and Human Health, University of Exeter, Truro, UK, [211]Department of Internal Medicine, Erasmus Medical Centre, Rotterdam, The Netherlands, [212]Department of Epidemiology, Erasmus MC University Medical Center, Rotterdam, The Netherlands, [213]Department of Public Health, Oxford University, Oxford, UK, [214]Center for Diabetes Research, Department of Clinical Science, University of Bergen, Bergen, Norway, [215]COPSAC Copenhagen Prospective Studies on Asthma in Childhood Copenhagen University Hospital, Herlev-Gentofte, [216]University of Queensland Diamantina Institute, The University of Queensland, Brisbane, Australia, [217]Institute of Medical Informatics, Biometry and Epidemiology, Chair of Epidemiology, Ludwig Maximilians University, Munich, Germany, [218]Helmholtz Center Munich, Institute of Epidemiology, Neuherberg, Germany, [219]Institute of Medical Statistics and Epidemiology, Technical University Munich, Munich, Germany, [220]Institute for Molecular Medicine Finland (FIMM), HiLIFE, University of Helsinki, Helsinki, Finland, [221]Netherlands Twin Register, Department of Biological Psychology, Vrije Universiteit, Amsterdam, Amsterdam, Netherlands, [222]Centre for Global Health Research, Usher Institute, University of Edinburgh, Teviot Place, Edinburgh, EH8 9AG, Scotland, [223]Genetics of Complex Traits, University of Exeter Medical School, University of Exeter, Royal Devon & Exeter Hospital, Exeter, U.K, [224]Research Centre for Optimal Health, School of Life Sciences, University of Westminster, London, U.K, [225]Department of Life Sciences, Centre for Inflammation Research and Translational Medicine, Brunel University London, London, U.K., [226]Institute of Translational Genomics, Helmholtz Zentrum München – German Research Center for Environmental Health, 85764 Neuherberg, Germany, [227]Technical University of Munich (TUM) and Klinikum Rechts der Isar, TUM School of Medicine, Ismaninger Str. 22, 81675 Munich, Germany, [228]Division of Gastroenterology, Hepatology and Nutrition, The Children,Äôs Hospital of Philadelphia, Philadelphia, USA, [229]Division of Human Genetics, Cincinnati Children's Hospital Medical Center, Center for Prevention of Preterm Birth, Perinatal Institute, Cincinnati Children's Hospital Medical Center, Department of Pediatrics, University of Cincinnati College of Medicine, Cincinnati, USA, [230]Division of Endocrinology, Department of Pediatrics, Boston Children's Hospital
